# Functionally enriched human polymorphisms associate to species in the chronic wound microbiome

**DOI:** 10.1101/2025.01.15.25320612

**Authors:** Rebecca A. Gabrilska, Khalid Omeir, Jacob Ancira, Clint Miller, Craig D. Tipton, Kendra P. Rumbaugh, Joseph Wolcott, Ashley Noe, Kumudu Subasinghe, Megan Rowe, Nicole Phillips, Caleb D. Phillips

**Affiliations:** Department of Surgery, Texas Tech University Health Sciences Center, Lubbock, USA; Department of Biological Sciences, Texas Tech University, Lubbock, USA; Southwest Regional Wound Care Center, Lubbock, USA; RTL Genomics, MicroGenDX, Lubbock, USA; Microbiology, Immunology & Genetics, University of North Texas Health Science Center, Fort Worth, USA; Natural Science Research Laboratory, Texas Tech University, Lubbock, USA

## Abstract

Chronic wounds are a burden to millions of patients and healthcare providers worldwide. With rising incidence and prevalence, there is an urgent need to address non-healing wounds with novel approaches. Impaired wound healing has been shown to be associated with wound microbiota, and multiple bacterial species are known to contribute to delays in closure. Recent evidence suggests human genetics may shape differences in composition of wound microbiomes, and unraveling this relationship has important implications for understanding wound bioburden and tailoring treatment. Here, a two-stage microbiome genome wide association study (mbGWAS; n=509) was used to test effects of human genetics on the relative abundances of bacterial species detected in chronic wounds using bacterial 16S rRNA gene sequencing. Sixteen species were significantly associated to 193 genetic loci distributed across 25 non-overlapping genomic regions. No locus was associated with more than one species, with heritability estimates per species ranging up to 20%. Functional analyses on genomic regions and species resulted in overrepresentation pertaining to pathways relevant to microbial infection and wound healing, suggesting that genetic and species interactions jointly influence the wound microenvironment. Species associated to host genetics in turn exhibited significant co-occurrence relationships with common wound pathogens including *Staphylococcus aureus* and *Finegoldia magna*. Moreover, the overall genetic distance among patients was significantly related to differences in their overall wound microbiome composition. Identification of such genetic biomarkers reveals new mechanistic insight into patient-microbiome interactions and provides an avenue to identify predictive risk factors for diagnosis and management of chronic wounds.

## INTRODUCTION

Chronic wounds, defined by impaired healing and associated with high patient morbidity, represent a significant clinical challenge. Ever-rising global incidence and prevalence of chronic wounds costs the healthcare system billions, increases provider burnout, and decreases patient quality of life.^1-3^ The wound microenvironment is an ecosystem influenced by host, microbiological, and environmental factors that together may contribute to delay of closure. Many human genes involved in healing have been characterized^4^ and a misbalance, whether a deficit, overabundance, or aberrance in any of these gene products contribute to disruption of healing. On the other hand, genes involved in the development of chronic wounds are less understood. Some candidate genes have been reported to be important to the formation of different types of chronic wounds including diabetic foot ulcers^5^, decubitus ulcers^6^, and venous ulcers^7^. These genes include those important to inflammation and immunity, angiogenesis and fibrosis, oxidative stress and metabolism, supporting the notion that wound susceptibility and healing is influenced by individual host physiology. Although candidate gene approaches are valuable, it is appreciated that the breadth of genetic regulation that influences chronicity is much greater than currently understood.

Wound care guidelines and consensus statements instructing the management of biofilm and bioburden is an important aspect of wound care.^8-14^ The approach to, and understanding of, wound microbial bioburden is evolving. This is primarily because DNA identification of microbes with next-generation sequencing (NGS) technologies have enabled comprehensive characterization of microbial communities, facilitating the exploration of microbial-patient interactions. This is important as the collective community of bacteria within wounds, known as the wound microbiome, has been shown to contribute to significant healing delays.^15,16^ Similarly, there is evidence that microbiomes of chronic wounds are dysbiotic, creating a hostile environment that obstructs healing.^17-19^ Whereas wound microbiomes are typically polymicrobial,^20-22^ the biology is additionally complicated by substantial variation in species’ incidence and abundance, as well as poorly known species-specific pathophysiological effects and species interactions.

The basis for wound microbiome compositional variation among individuals is still not well understood but is considered multifaceted. Among plausible determinants of wound microbiome composition, and potentially ultimately consequential healing, are differences in individual patient genetics. In the only microbiome genome wide association study (mbGWAS) focusing on chronic wounds to date studied the relationship between microbial diversity to human polymorphisms.^23^ Although sample sizes were modest in this pilot study, loci were repeatedly associated across two independent cohorts, and genes adjacent to associated loci were functionally enriched, particularly for cell surface and integrin-related processes. Further, post-hoc experiments supported a genetic effect on differences in wound bed physiology hypothesized to influence wound microbiomes. While Tipton *et al.* (2020) documented for the first time how human genetics can directly influence composition of the wound microbiome, the extent and mechanistic diversity of this relationship is unknown. However, given the observed species and community diversity of wound microbiomes, it is anticipated that a variety of mechanisms shape interactions between microbes and the host during wound healing.

This study helps elucidate the human genetic basis of the wound microbiome by conducting an mbGWAS enrolling over 500 patients diagnosed with chronic wounds who presented to an outpatient wound care clinic. First, we identified human genetic variants (single nucleotide polymorphisms (SNPs) and length polymorphisms) significantly associated with the relative abundances of bacterial species in wounds. We investigated associated loci by studying the genes or pathways they are known or thought to influence and evaluated these functions. Further, to study how selection of specific species by genetics may more broadly shape community structure, we investigated abundance relationships among species and the effect of genomic distance on overall wound microbiome composition. By elucidating genetic predisposition to specific wound microbes and the genomic relationship to the wound microbiome more broadly, we contribute to an improved understanding of chronic wound pathogenesis.

## METHODS

### Ethical considerations

Human study protocol was executed after obtaining informed written consent from adult patients as approved under WCG IRB Protocol 56-RW-156DNA.

### Patient recruitment and sample collection

An overview of methods for mbGWAS is presented in Figure 1. Samples were collected from consenting patients receiving outpatient wound care at the Southwest Regional Wound Care Center in Lubbock, Texas (Figure 1a). Patient enrollment and collection of buccal swabs for genotyping spanned September 2017 to March 2023. Inclusion criteria included: bacterial profiling by 16S rRNA gene sequencing at initial assessment, adults older than 18 years of age presenting with a physician-determined full thickness, non-healing wound and candidates for sharp debridement therapy. Patients with incomplete or incongruent record information were excluded from the study, but were not excluded based on wound type (e.g., diabetic ulcer, venous ulcer, decubitus ulcer) or anatomical location. After consent, a single buccal swab for human genotyping was collected as previously described^23,24^ and immediately placed in liquid nitrogen for cryogenic archival in the Wolcott Wound Care Research Collection at the Natural Science Research Laboratory, Museum of Texas Tech University (Lubbock, Texas). As standard of care at the participating clinic, post-debridement wound tissue for bacterial community profiling were collected as previously described^23,24^ and sent to MicroGenDX (Lubbock, Texas), which is a College of American Pathologist (CAP) accredited and Clinical Laboratory Improvement amendments (CLIA) certified high complexity molecular diagnostic laboratory. Deidentified electronic medical records for consenting patients were acquired to collect demographic information, including sex, age, self-identified race or ethnicity, and past medical history (e.g., diagnosis of diabetes mellitus).

**Figure 1.**
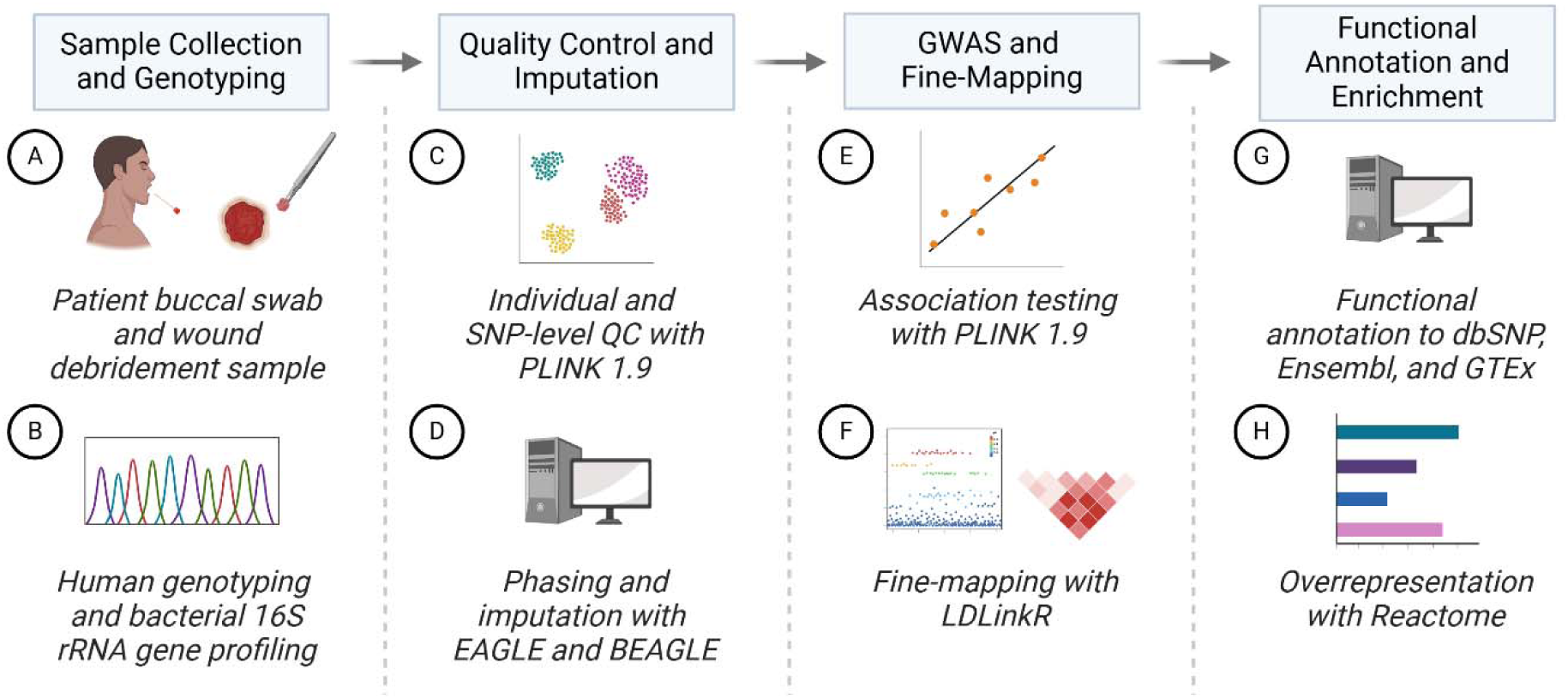
Overview of mbGWAS workflow used in this study. (a) Patient buccal swab and wound debridement samples were collected for (b) human genotyping and bacterial 16S rRNA gene sequencing, respectively. Microbiome results were collected from electronic records reported as relative abundance of species with a 1% study-wide incidence threshold. (c) Human genotype files were processed for individual- and SNP-level QC. To increase genomic density, (d) tag SNPs were imputed. (e) Association testing was performed between genomic variants and relative abundances of bacterial species, followed by (f) fine-mapping of associated SNPs to expand to candidate SNPs in strong LD. (g) Candidate loci were annotated using dbSNP, Ensembl, and GTEx databases for proximal loci and QTLs. (h) Candidate genes were summarized with ontological overrepresentation analysis. Created in BioRender.com.

### Wound microbiome profiling

Sequencing results (species relative abundances) of wound debridement samples from the earliest clinical visit for one wound per patient were taken directly from the electronic medical record and analyzed for bacterial microbiome composition (Figure 1b). Because sample collection spanned many years, the relative abundances of bacterial taxa in wounds were characterized as previously described using 454 Titanium Instrument (454 Life Sciences, Roche)^24^, Ion Torrent PGM (Thermo Fisher Scientific)^23^ or Illumina MiSeq^25^ at MicroGenDX (Lubbock, Texas). Clinical repeatability and congruence in community profile reporting between instruments was previously demonstrated during CAP/CLIA accreditation and certification, respectively.^25^ Additional thresholding included a study-wide incidence rate of 1%, and relative abundances were centered log-ratio (clr)-transformed using the clr function from the compositions package in R.^26^ Patients with inconclusive 16S rRNA gene sequencing results (i.e., insufficient sample) or only reported with species below the incidence threshold were excluded.

### Buccal DNA genotyping and imputation

Buccal swabs were processed for DNA extraction using the Gentra Puregene Buccal Kit (Qiagen). Briefly, swabs were lysed overnight at 55°C, followed by DNA purification with RNase A and a final elution in 50 µL molecular biology grade water (HyClone Hypure, GE Healthcare Life Sciences). Samples were quantified for DNA using a Qubit 3.9 fluorometer (ThermoFisher Scientific). All samples were titrated or dehydrated to a final concentration of 10-50 ng/µL and stored at -80°C prior to genotyping. Patients and samples included in Tipton, *et al.*^23^ derived from the same clinic were also included in this study. Samples were processed on the Illumina Infinium Global Screen Array v3.0. Data were collected using the iScan System (Illumina), and preliminary processing/analysis (i.e., normalization, SNP clustering, genotype calling, call rate calculations, array-based quality control (QC)) were conducted using GenomeStudio v.2.0 (Illumina). PLINK file sets were exported using the PLINK Input Report Plug-in v.2.1.4 within GenomeStudio.

Prior to QC filtering, genome sequencing arrays were merged based on intersecting Reference SNP cluster ID (RSID) using PLINK v1.9b7^27,28^. Multiallelic, duplicated, and variants with allele flip errors were excluded. Pre-imputation QC filtering was subsequently performed as previously reported^23^ with additional filtering to improve imputation accuracy using PLINK v1.9b7 (Figure 1c). Briefly, individual-level QC first removed individuals based on (1) elevated missing genotyping rates (>=0.09) or outlying heterozygosity rate (>3 standard deviations); (2) high degree of allele relatedness (identity-by-descent > 0.1875); or (3) high degree of ancestral divergence via principal component analysis. SNP-level QC removed variants with (1) low genotyping rates (<0.05); (2) Hardy-Weinberg equilibrium p-value deviations (<1×10^-5^)^29,30^; (3) low minor allele frequency (MAF, <0.05); and (4) variants on X, Y, and mitochondrial chromosomes were not included.

To expand variant density, phasing was performed with EAGLE 2.4.1^31^ followed by genome imputation with BEAGLE 5.4^32^ using the 1000 Genome Project phase 3 samples as the reference panel (Figure 1d). Haplotypes for phasing were estimated using the GRCh37/hg19 reference map. Imputed variants with a dosage R-squared value of <0.8 were omitted. The resulting SNPs were queried using USCS’s bigBedNamedItems command-line to convert to GRCh38/hg38 build.^33^ Lastly, a MAF threshold of 5% was employed post-liftover.

### Genome-wide association testing

Variables thought to influence wound healing and microbiome composition led to the inclusion of age, sex, and diagnosis of diabetes mellitus as covariates in our model. Additionally, we controlled for effects of genetic ancestry by inclusion of principal component analysis (PCA) eigenvectors. For this, variance normalized PCA was conducted with PLINK 1.9vb7 post-QC. The explanatory power of eigenvectors on self-described racial or ethnic category was assessed using PERMANOVA with Euclidean distances in which the explanatory power of vectors 1-10 on the dummy coded race/ethnicity matrix was assessed. Eigenvectors 1 and 2 were significant and explained ∼86% of variance, whereas eigenvector 3 was not significant and eigenvectors 3-10 cumulatively explained only 0.2% of remaining variation. Therefore, vectors 1 and 2 were also included as covariates in subsequent association testing, which is consistent with expectations of K-1 (K = number of populations) significant vectors as previously reported.^34^

A two-stage, exploratory-replication cohort design was employed by evenly stratifying individuals who passed QC filtering across both bacterial incidence and self-identified race or ethnicity. The exploratory-replication cohorts were repeated *inversus*, *i.e.*, the original exploratory reassigned to the replication cohort and the original replication reassigned to the exploratory cohort and the associations ran a second time. Genotype dosages plus five covariates (age, sex, presence of clinical history of diabetes mellitus, and eigenvectors 1 and 2) were included in linear regressions using PLINK v1.9b7 (Figure 1e). QQ-plots and Manhattan plots were calculated using the R package ‘qqman’^35^. From each GWAS, λ was calculated as the ratio of the median of the observed and expected chi-squared statistic with one degree of freedom. To determine repeatedly significant variants a suggestive threshold of two log_10_ lower than the strict Bonferroni correction (α = 0.05/6186583) was used for the exploratory cohort, followed by a strict Bonferroni adjusted significance threshold of <0.05 for the replication cohort.

### Fine-mapping of associated variants

Regions on the GRCh38.p14 assembly encompassing associated variants were fine-mapped to identify nearby variants in linkage disequilibrium (LD) using the LDproxy tool and R package ‘LDlinkR’^36^ (Figure 1f). The 1000 Genomes Project served as reference, and RSIDs that were in very strong LD (*R^2^*>0.8) to associated SNPs among all populations were identified using ‘LDproxy_batch’ function. Moving forward, the associated variants and those in very strong LD were altogether considered candidate variants.

All candidate variants were subsequently grouped into genomic regions. Here, a genomic region was defined as an approximately 250Kb genomic region inclusive of candidate variant sets. For visual representation, regions were plotted using the R package ‘karyoploteR’^37^.

### Measuring additive effects of candidate variants

Because genomic regions often contained multiple candidate variants, and because species were sometimes associated with candidate variants across multiple regions, the independent and additive effects of candidate variants explaining species’ relative abundance was tested. For this analysis, each species’ relative abundance was treated as the response variable and all candidate variants associated to that species were included as predictors in the initial model. Using the R package ‘olsrr’^38^, variants were successively removed from the model based on largest p-value until either any remaining variants were significant, or no variants remained. Following this procedure for each species, the relationship of effect sizes (R^2^) among models and their respective number of candidate loci or final model with predictor variants was tested with simple regression.

### Searching variants in public databases

The NCBI dbSNP database^39^ and NHGRI-EBI GWAS catalog^40^ were accessed for all 193 candidate variants to determine whether previous reports have associated these loci in other settings. Both databases were accessed 20 December 2024.

### Functional annotation of candidate variants

To perform functional annotation on candidate variants, R package ‘rsnps’^41^ was used to query the NCBI dbSNP database (batch v157) and the ANNOVAR tool^42^ to query the Ensembl database (release 113)^43^ (Figure 1g). Data collected included genomic location, variant class, potential consequence (i.e., intron, intergenic, exon), and proximal genes. Additionally, to identify genes potentially regulated (e.g., gene expression or splicing variation) by candidate SNPs, we assessed whether loci overlapped with expression quantitative trait loci (eQTL) or splicing quantitative trait loci (sQTL) in the Genotype-Tissue Expression Portal (GTEx v8)^44^ (Figure 1g).

### Overrepresentation analysis

To identify enriched biological processes and pathways, we performed pathway overrepresentation analysis using the Reactome database (v90)^45^ (Figure 1h). First, biomaRt (v2.62.0)^46^ was used to convert the Ensembl gene IDs to their corresponding Entrez gene IDs. Ensembl gene IDs without corresponding Entrez gene IDs were not included. Next, to facilitate overrepresentation analyses, we leveraged R packages ‘clusterProfiler’ (v4.14.0)^47^ and ‘ReactomePA’ (v1.50)^48^ with a threshold at FDR<0.05. To investigate different perspectives about how human functional variation may relate to bacterial abundances, enrichment analysis was conducted study-wide, based on predicted tissue-specificity of effect and for each species. Last, we employed the ‘ReactomeContentService4R’ package (v90)^49^ to retrieve the corresponding top Reactome pathways.

### Bacterial co-abundance analysis

To expand species-specific genetic associations into a broader multi-species community framework, a co-occurrence analysis that utilizes species relative abundance was used to assess significance of correlations among all species. The R function ‘REBACCA’^50^, which controls bias when comparing compositional data, was implemented with 100 bootstrap replicate subsampling individuals. Correlated species pairs are identified with FWER<0.05.

### Microbiome composition-patient genomic distance relationship

Because results from other analyses supported patient genetic effects on species abundances as well as significant co-abundance patterns among species, the broader relationship between human genetics and microbiome composition was explored. For this, microbiome composition was calculated as the Bray-Curtis dissimilarity metric using the R package ‘vegan’. Genomic distance among individuals were calculated as Euclidean distances using either all candidate variants, or only the candidate variants from each genomic region with the smallest p-value (the latter method was also considered because high correlation among nearby candidate variants can introduce noise in the distance matrix). The relationship among microbiome and genomic distance matrices was assessed using multiple regression of matrices with the R package ‘ecodist’^51^, controlling for covariates (age, sex, diagnosis of diabetes mellitus, effects of genetic ancestry) and implemented with 999 permutations for significance testing. A threshold of p<0.05 was considered significant.

## RESULTS

### Association of human genetic variation to species relative abundances

Following quality control, 509 patients were retained for mbGWAS. The average age was 66.8 (sd=13.2), with 61% males and 39% females. Approximately 64% of patients had a diagnosis of diabetes mellitus. Controlling for age, sex, diabetes diagnosis, and genetic ancestry, association testing of imputed allelic dosages of 6,186,583 variants to the relative abundances of 68 bacterial species was performed using a two-stage (exploratory-replication) *inversus* cohort design. At a suggestive threshold of p<8.08×10^-7^, there were 51,454 loci across 68 species which included 18,313 in cohort 1 and 33,165 in cohort 2. Manhattan and QQ plots of all conditions are in Supplementary Data 1a and 1b, in addition to the genomic inflation factor (λ) in Supplementary Data 1c. Each set of suggestive loci were subsequently tested in their respective confirmatory cohort using a strict Bonferroni correction at α<0.05. This testing resulted in 98 unique repeatedly significant loci associated across 16 bacterial species. The median number of associations across species was 2.5, with *Anaerococcus octavius* having the most associations with 16. Association results of significant loci for both cohorts are further detailed in Supplementary Data 2.

To increase odds of including causal variants and relating associations to genes of potential effect, patterns of LD to the associated loci were also evaluated. For this, LDlinkR was used for fine-mapping through which loci in strong LD (i.e., *R^2^*>0.8) to the associated loci were identified. This resulted in 95 additional loci for consideration, and 15 of these were not originally available in the imputed data set. Altogether, mbGWAS plus fine-mapping resulted in 193 variants across 16 bacterial species. This set of 98 associated loci and 95 LD-SNVs are collectively referred to as candidate variants and are listed with linkages in Supplementary Data 3. Although there were 8 RSIDs with prior reports in the dbSNP or GWAS catalog databases, none of these variants were previously linked with skin, wounds, microbes, or infection (Supplementary Data 4).

### Candidate variants for species are distributed across different genomic regions

Comparison of chromosome and base pair locations indicated that candidate variants originate from 25 genomic regions, defined here as within an approximately 250kbp window, across 14 autosomal chromosomes (Figure 2). Chromosomal base pair locations and identifiers, species associations, number of candidate and associated variants per region are summarized in Table 1. Four species were associated to multiple regions and included *Streptococcus anginosus* (5), *Anaerococcus obesiensis* (3), *Citrobacter koseri* (3), and *A. octavius* (2). For the remaining 12 species associated with one region, two species (*Burkholderia gladioli*, *Corynebacterium jeikeium*) only had a single variant in their respective region, and six species (*Corynebacterium amycolatum*, *Dialister propionicifaciens*, *Fusobacterium canifelinum*, *Pasturella canis*, *Prevotella bivia*, *Staphylococcus simulans*) had less than 5 variants per region. However, four species had at least 5 variants, including *Streptococcus dysgalactiae* (44), *Veillonella parvula* (16), *Staphylococcus pseudintermedius* (11), and *Bacteroides fragilis* (5). The genomic region with the most variants (44) spans approximately 260kbp on chromosome 2 and was associated with *S. dysgalactiae*. This was followed by a 73kbp region on chromosome 1 that had 43 variants associated to *A. octavius*.

**Figure 2.**
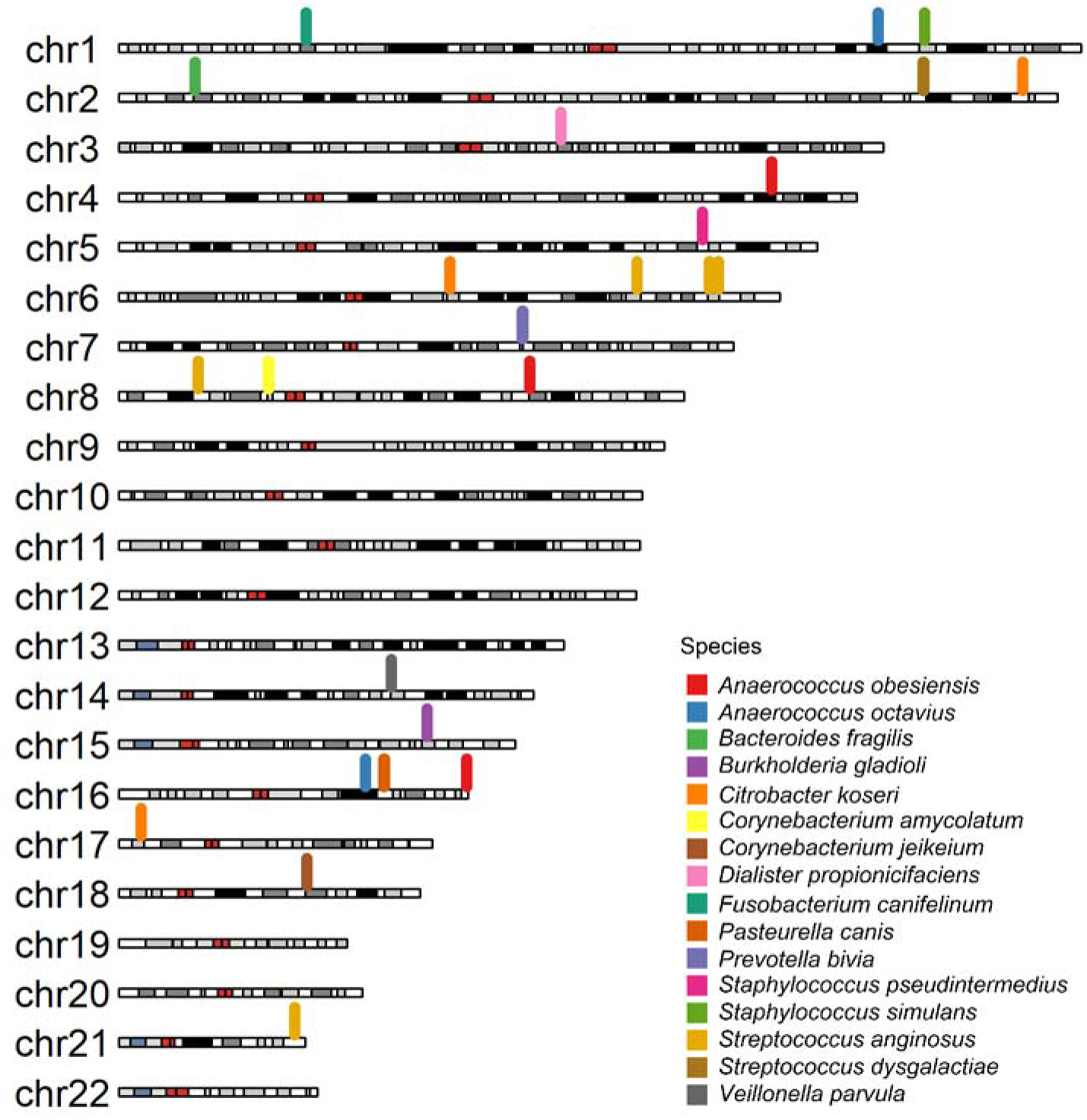
Chromosome ideogram illustrating locations of genomic regions for all 193 candidate loci. The 193 candidate variants clustered across 25 genomic regions of approximately 250kbp. Individual colors represent each of 16 associated bacterial species, while colors on chromosomes indicate centromeres (red) and cytobands (white, gray, black).

**Table 1.** Genomic regions with associated bacterial taxa and respective variants. Each of the 25 genomic regions ranging from 0 to approximately 266Kb is associated with one of the 16 associated bacterial taxa and contains between 1-44 candidate variants.

| Genomic Region | Bacterial Taxa | Number of Candidate Variants | Number of Associated Variants | Chromosome | Starting Position | Ending Position | Genomic Size (bases) | RSID of Candidate Variants |
| --- | --- | --- | --- | --- | --- | --- | --- | --- |
| 1 | *Anaerococcus obesiensis* | 1 | 1 | 4 | 169072680 | 169072680 | 0 | rs747279 |
| 2 | *Anaerococcus obesiensis* | 18 | 10 | 8 | 106320218 | 106357764 | 37546 | rs116952730;<br>rs1170264083;<br>rs12680041;<br>rs12682257;<br>rs144684346;<br>rs149573143;<br>rs150326499;<br>rs17251771;<br>rs17336699;<br>rs75473788;<br>rs75730553;<br>rs76232722;<br>rs77452376;<br>rs77600282;<br>rs78277517;<br>rs79097397;<br>rs79823074;<br>rs996097462 |
| 3 | *Anaerococcus obesiensis* | 2 | 2 | 16 | 89943309 | 89947728 | 4419 | rs117424452;<br>rs145268882 |
| 4 | *Anaerococcus octavius* | 43 | 13 | 1 | 196533814 | 196607234 | 73420 | rs10399651;<br>rs10737676;<br>rs10737678; |
|  |  |  |  |  |  |  |  | rs10754192;<br>rs10754193;<br>rs10754194;<br>rs10754197;<br>rs10922087;<br>rs12072649;<br>rs12128544;<br>rs12133987;<br>rs13375443;<br>rs1360143;<br>rs1416960;<br>rs1538683;<br>rs1571489;<br>rs200035495;<br>rs2335906;<br>rs35988088;<br>rs4072395;<br>rs4294368;<br>rs4314834;<br>rs4405100;<br>rs4515738;<br>rs4518841;<br>rs4562566;<br>rs4601531;<br>rs4614205;<br>rs4625226;<br>rs4633231;<br>rs4658040;<br>rs4658042;<br>rs4658043;<br>rs5779839;<br>rs6663649;<br>rs6680890; |
|  |  |  |  |  |  |  |  | rs6688468;<br>rs73069765;<br>rs7533533;<br>rs7540922;<br>rs7552170;<br>rs869047855;<br>rs869128137 |
| 5 | *Anaerococcus octavius* | 6 | 3 | 16 | 63717446 | 63733603 | 16157 | rs10500476;<br>rs16966464;<br>rs57243990;<br>rs7204388;<br>rs73583531;<br>rs9924260 |
| 6 | *Bacteroides fragilis* | 5 | 1 | 2 | 19718575 | 19726699 | 8124 | rs11356656;<br>rs12619053;<br>rs1666756894;<br>rs6708954;<br>rs72775105 |
| 7 | *Burkholderia gladioli* | 1 | 1 | 15 | 79811673 | 79811673 | 0 | rs141101943 |
| 8 | *Citrobacter koseri* | 1 | 1 | 2 | 233997402 | 233997402 | 0 | rs6752001 |
| 9 | *Citrobacter koseri* | 1 | 1 | 6 | 85747760 | 85747760 | 0 | rs78568090 |
| 10 | *Citrobacter koseri* | 4 | 3 | 17 | 5646305 | 5659654 | 13349 | rs10459902;<br>rs12949515;<br>rs35083030;<br>rs6502871 |
| 11 | *Corynebacterium amycolatum* | 2 | 2 | 8 | 38632020 | 38668740 | 36720 | rs116980906;<br>rs117956478 |
| 12 | *Corynebacterium jeikeium* | 1 | 1 | 18 | 48674597 | 48674597 | 0 | rs8085434 |
| 13 | *Dialister* | 2 | 2 | 3 | 114392560 | 114396415 | 3855 | rs1105821; |
|  | *propionicifaciens* |  |  |  |  |  |  | rs1402444 |
| 14 | *Fusobacterium canifelinum* | 2 | 2 | 1 | 48493905 | 48508278 | 14373 | rs17470284;<br>rs79415770 |
| 15 | *Pasteurella canis* | 3 | 3 | 16 | 68485089 | 68759496 | 274407 | rs117559006;<br>rs12444852;<br>rs79045992 |
| 16 | *Prevotella bivia* | 4 | 1 | 7 | 104469967 | 104486719 | 16752 | rs10216054;<br>rs75782629;<br>rs76224663;<br>rs76324407 |
| 17 | *Staphylococcus pseudintermedius* | 11 | 8 | 5 | 151030428 | 151051135 | 20707 | rs2233302;<br>rs2233311;<br>rs72790107;<br>rs72790109;<br>rs72790110;<br>rs72790113;<br>rs72790114;<br>rs72790116;<br>rs72790117;<br>rs7713223;<br>rs8177834 |
| 18 | *Staphylococcus simulans* | 2 | 2 | 1 | 208679576 | 208682580 | 3004 | rs11579666;<br>rs61813642 |
| 19 | *Streptococcus anginosus* | 2 | 2 | 6 | 134100550 | 134106344 | 5794 | rs12663809;<br>rs9399087 |
| 20 | *Streptococcus anginosus* | 10 | 9 | 8 | 20481821 | 20485358 | 3537 | rs10103232;<br>rs10107126;<br>rs12679719;<br>rs17092580;<br>rs28436903;<br>rs28506103;<br>rs28562717;<br>rs7846516; |
|  |  |  |  |  |  |  |  | rs9325869;<br>rs9325870 |
| 21 | *Streptococcus*<br>*anginosus* | 7 | 7 | 21 | 45422585 | 45428241 | 5656 | rs11911327;<br>rs200093729;<br>rs200571781;<br>rs74673807;<br>rs76241995;<br>rs78833614;<br>rs79785189 |
| 22 | *Streptococcus*<br>*dysgalactiae* | 44 | 5 | 2 | 208264090 | 208530434 | 266344 | rs10165453;<br>rs10173938;<br>rs10176626;<br>rs10182087;<br>rs10189031;<br>rs10932260;<br>rs11693993;<br>rs11899172;<br>rs12617780;<br>rs13392540;<br>rs13393025;<br>rs1347665;<br>rs1561239;<br>rs1584200;<br>rs1898435;<br>rs1898437;<br>rs1900239;<br>rs2118294;<br>rs2304545;<br>rs2363466;<br>rs3856350;<br>rs4234089;<br>rs4293511;<br>rs4462735; |
|  |  |  |  |  |  |  |  | rs4673391;<br>rs4673393;<br>rs4673394;<br>rs4675748;<br>rs4675761;<br>rs6435448;<br>rs6704628;<br>rs6709293;<br>rs6724208;<br>rs6725548;<br>rs6736599;<br>rs6752329;<br>rs6757684;<br>rs7583022;<br>rs7587752;<br>rs869262013;<br>rs892359390;<br>rs9288392;<br>rs959660;<br>rs970377 |
| 23 | *Veillonella*<br>*parvula* | 16 | 13 | 14 | 70427998 | 70456344 | 28346 | rs11621649;<br>rs11625103;<br>rs11625292;<br>rs11629001;<br>rs11846101;<br>rs1284590816;<br>rs199616474;<br>rs34817626;<br>rs61979083;<br>rs61979102;<br>rs61979103;<br>rs61979104;<br>rs61979105; |
|  |  |  |  |  |  |  |  | rs8004222;<br>rs8013270;<br>rs868264570 |
| 24 | *Streptococcus*<br>*anginosus* | 2 | 2 | 6 | 152835266 | 152970748 | 135482 | rs112442552;<br>rs76686834 |
| 25 | *Streptococcus*<br>*anginosus* | 3 | 3 | 6 | 155143411 | 155180671 | 37260 | rs138845881;<br>rs141057867;<br>rs183941014 |

### Subsets of variants have additive effects on species relative abundances

To evaluate additive effects of variants explaining bacterial relative abundances, we performed backward stepwise regression. For these analyses, individuals from both cohorts were combined, species relative abundances were response variables, and all candidate variants for a given species were initial predictor variables. At least one variant was retained in the final model for 12 of the 16 associated bacterial taxa. Between 1 and 6 variants significantly influenced relative abundances with effect sizes (R^2^) ranging from approximately 1.5% to 20% (Figure 3a). The highest effect size (20.1%) was for *S. dysgalactiae*, which was contributed by 6 of its initial 42 candidate variants. Seven species were associated to 1 variant with effect sizes ranging between 2.3-6.8%. Whereas no relationship was observed between effect sizes of the reduced model and total number of candidate variants per taxa (Figure 3b; p=0.082), a positive relationship was observed between effect size and number of predictor variants (Figure 3a; p<0.0001, β=0.026, error=0.0042, adjusted R^2^=0.72).

**Figure 3.**
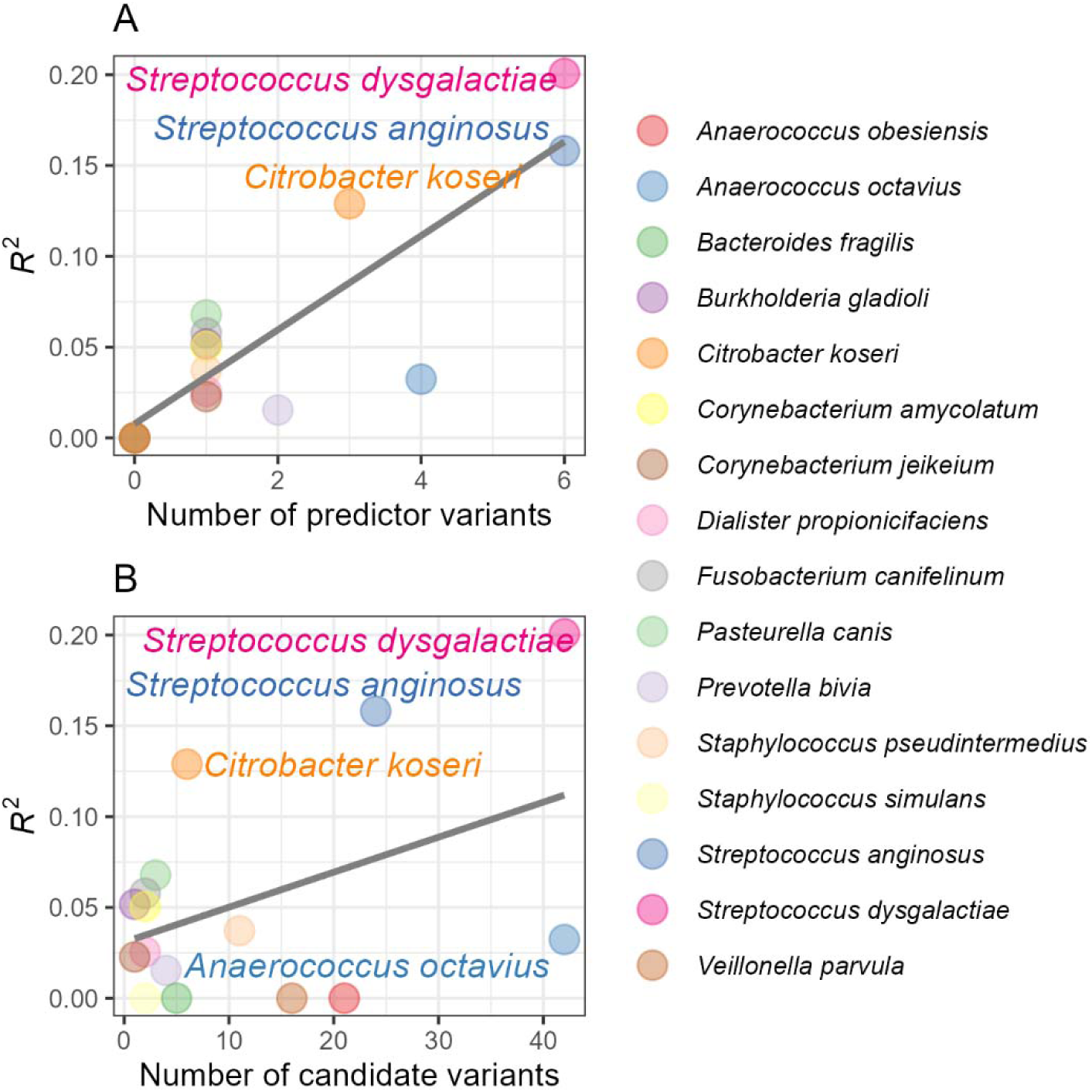
Relationships of backward selected regression model effect sizes for each of the associated bacterial species to the (a) number of predictor variants retained in the final models and (b) number of candidate variants that were initially considered for each model.

### Functional annotation of candidate variants

Candidate variants were queried to the NCBI SNP and Ensembl genome databases to detail variant information including class (SNV or INDEL), position (e.g., intron, intergenic, exon), and proximal genes (Supplementary Data 5). Among 193 candidate variants, a majority (93%) were SNPs and the remaining were INDELs. Also, the majority (71%) of variants were intronic sequences, 19% were intergenic, and 8 were exonic.

Among the exonic variants, three are in the collagen type XVIII alpha 1 chain antisense RNA 1 locus (*COL18A1-AS1*; rs76241995, rs78833614, rs79785189) and located in region 21. rs7204388 is a non-coding exon variant located in *LINC02165* (Long Intergenic Non-Protein Coding RNA 2165). A fifth exonic variant, rs2233311, which is in strong LD with eight other non-exonic candidates, is a nonsynonymous SNP (C>A,G) within the TNF alpha induced protein 3-interacting protein 1 (*TNIP1*) gene. The sixth exonic variant, rs2304545 which is in strong LD with one other non-exonic candidate is a synonymous SNV (G>A,C,T) located within the FYVE finger-containing phosphoinositide kinase (*PIKFYVE*) gene. The final two exonic variants, rs76686834 and rs8177834, are located within 3’UTR of genes F-Box protein 5 (*FBXO5*) and *TNIP1*, respectively.

To further annotate candidate variants to potential function, the Genotype-Tissue Expression (GTEx) Portal was used to identify if any candidate variants were previously identified as expression and splicing quantitative trait loci (eQTL and sQTL) (Supplementary Data 5). Among 193 candidate variants, 67 (34.7%) were also GTEx eQTLs, with 22 originating as associated loci and the remaining 45 as LD-SNPs. The eQTLs were distributed across 12 of the 25 genomic regions described above. These eQTLs have been predicted to influence expression of 35 genes across 33 tissues, with a majority (89.6%, 60/67) influencing gene expression in eight tissues of a lower extremity wound environment (sun-exposed skin, non-sun exposed skin, subcutaneous adipose tissue, skeletal muscle, tibial nerve, tibial artery, primary fibroblast culture, and whole blood). This subset of 60 wound-associated eQTLs predicted expression of 18 genes. For variants associated with splicing variation, 72 candidate variants are GTEx sQTLs associated with 135 splice variants of 15 genes in 41 tissues distributed across 7 genomic risk regions. Nearly all (98.6%, 71/72) sQTLs are associated with splicing variation in wound-relevant tissues with 33 splice variants of 13 genes. Genomic regions as defined herein, on average contained candidate variants related to the expression of, or otherwise proximal to, 3.4 genes (median=2, range=1-12).

Overrepresentation analysis revealed distinct functions for species and genomic regions Multiple functional overrepresentation analyses were conducted using subsets of the overall set of 96 genes which included 50 GTEx genes and 46 genes proximal to candidate variants. Analysis based on the overall set (i.e., all genes from all phenotypes) resulted in no enriched terms. Focusing enrichment analysis to only genes tied to known QTLs for all tissues or wound-relevant tissues also resulted in no enriched terms. However, when conducting overrepresentation for each species containing at least three genes (nine species), a total of 25 enriched terms were observed across five species (*A. obesiensis*, *B. gladioli*, *C. koseri*, *F. canifelinum*, *V. parvula*; Figure 4a). Overrepresentation analysis was also conducted for each genomic region (nine regions) through which 66 enriched terms were distributed across seven regions (3, 7, 10, 14, 21, 23, 24; Figure 4b). Supplementary Data 6 details the genomic regions, associated species, and corresponding enriched terms. There was a total of 19 top level Reactome pathways represented among significant terms. For these, ‘extracellular matrix organization’ was the most abundant (10), followed by ‘cell cycle’ and ‘vesicle-mediated transport’ (7), and ‘metabolism of proteins’ (5).

**Figure 4.**
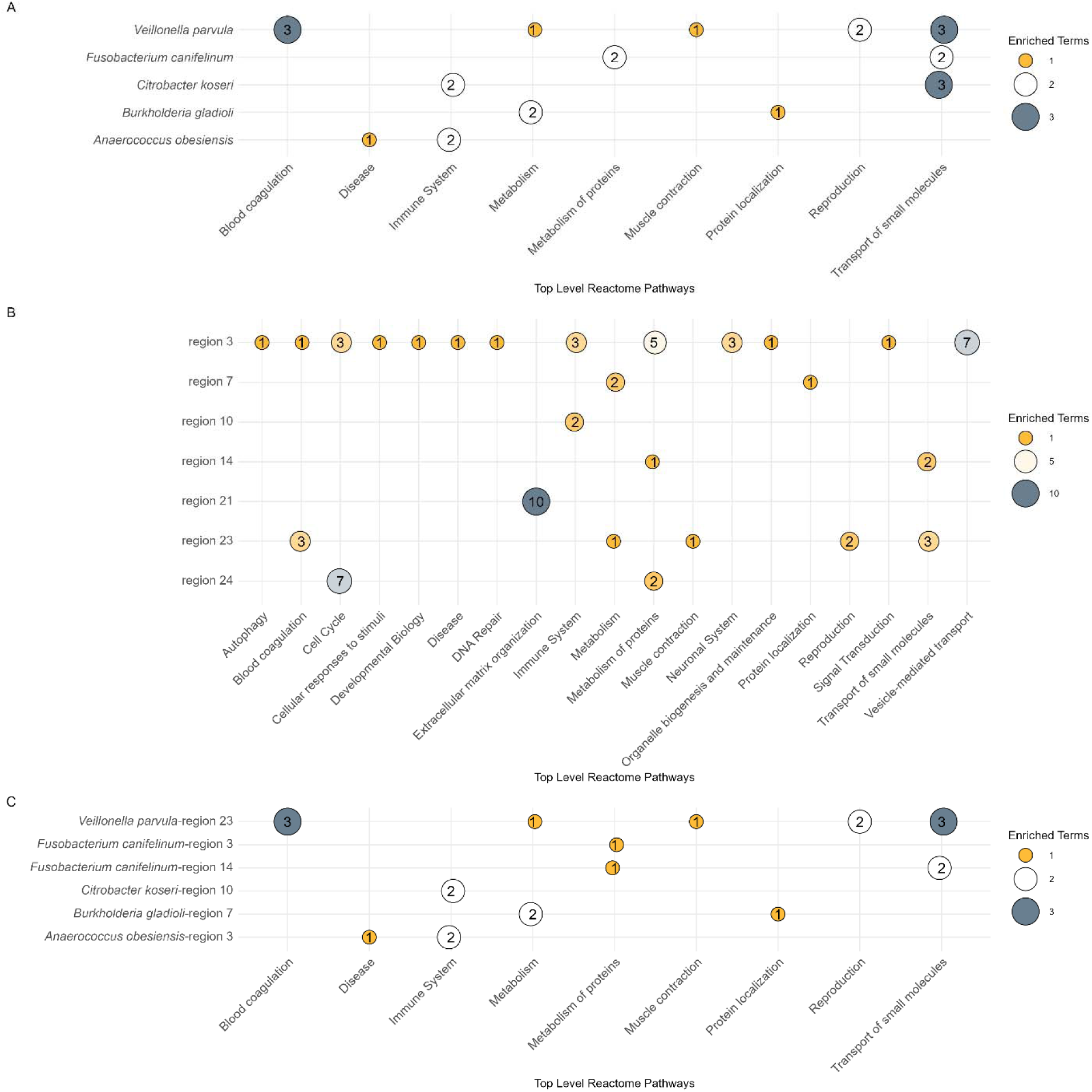
Overrepresentation of pathways for candidate loci. Input for Reactome overrepresentation analysis included eQTL, sQTL, and proximal genes to candidate variants and clustered by (a) bacterial species or (b) genomic region. (c) Of taxa-region enrichments, 9 top-level Reactome pathways characterized 6 taxa-region combinations.

Because some species were associated with multiple genomic regions, it was important to understand if species- (Figure 4a) and region-level (Figure 4b) groupings of genes provided additional or redundant perspectives about overrepresentation. When intersecting Reactome enriched pathways between the genomic regions and associated species subsets (e.g., “taxa-regions”), 22 terms overlapped for 6 region-species combinations (Figure 4c). Of these overlapping taxa-region loci, although both *A. obesiensis* and *C. koseri* associated to three regions (Table 1), only a single region is overrepresented for each species, meaning that terms or pathways commonly shared among species originated from different genomic regions. *B. gladioli* and *V. parvula* are each associated with one region (Table 1), and each taxa-region pairing has overrepresented Reactome pathways (Figure 4c). Finally, *F. canifelinum* is associated with only a single region (14) that is enriched for “Carboxyterminal post-translational modifications of tubulin”, yet this term was enriched in two regions; these enrichments arise from AGBL4 (region 14) and TUBB3 (region 3) genes.

### Associated species co-occur with common wound pathogens

In addition to the potential that host genetics selects for wound colonizing microbes, we hypothesize that genetically-selected species further shape the wound microbiome. Most wounds are polymicrobial, with approximately 82% of our samples harboring more than 2 species and a median of 3 species per wound. Here, we used co-abundance analysis to indicate potential interactions between species. This approach utilized relative abundance and controlled the number of co-occurrences to reduce spurious correlations and therefore is not affected by study-wide relative abundance. All 16 associated species significantly covaried (FWER<0.05) with at least one other species totaling 119 significant relationships (Figure 5). Co-abundance interactions were mostly positive (70.6%). Among GWAS-associated species, *D. propionicifaciens* had the fewest co-abundances (2) and *B. fragilis* had the most (18). Assessing co-abundance with non-associated species, *Staphylococcus aureus* (13), *Staphylococcus epidermidis*, *Finegoldia magna*, and *Anaerococcus vaginalis* (12) had the most significant co-abundances to associated species. The top five species*-*species combinations with the strongest positive correlations were *A. octavius*-*Anaerococcus prevotii* (0.417), *F. canifelinum-Fusobacterium nucleatum* (0.334), *D. propionicifaciens-Prevotella buccalis* (0.285), *B. gladioli-Escherichia coli* (0.271), *F. magna-A. octavius* (0.229). Of the 16 bacterial species identified herein associated with human variants, 2 are obligate aerobes (*B. gladioli, C. jeikeium*), 7 are obligate anaerobes (*A. obesiensis, A. octavius, B. fragilis, D. propionicifaciens, F. canifelinum, P. bivia, V. parvula*), and the remaining 7 species are considered facultative anaerobes (*C. koseri, C. amycolatum, P. canis, S. pseudintermedius, S. simulans, S. anginosus, S. dysgalactiae*). However, aerotolerance did not explain the co-abundance patterns (p>0.05).

**Figure 5.**
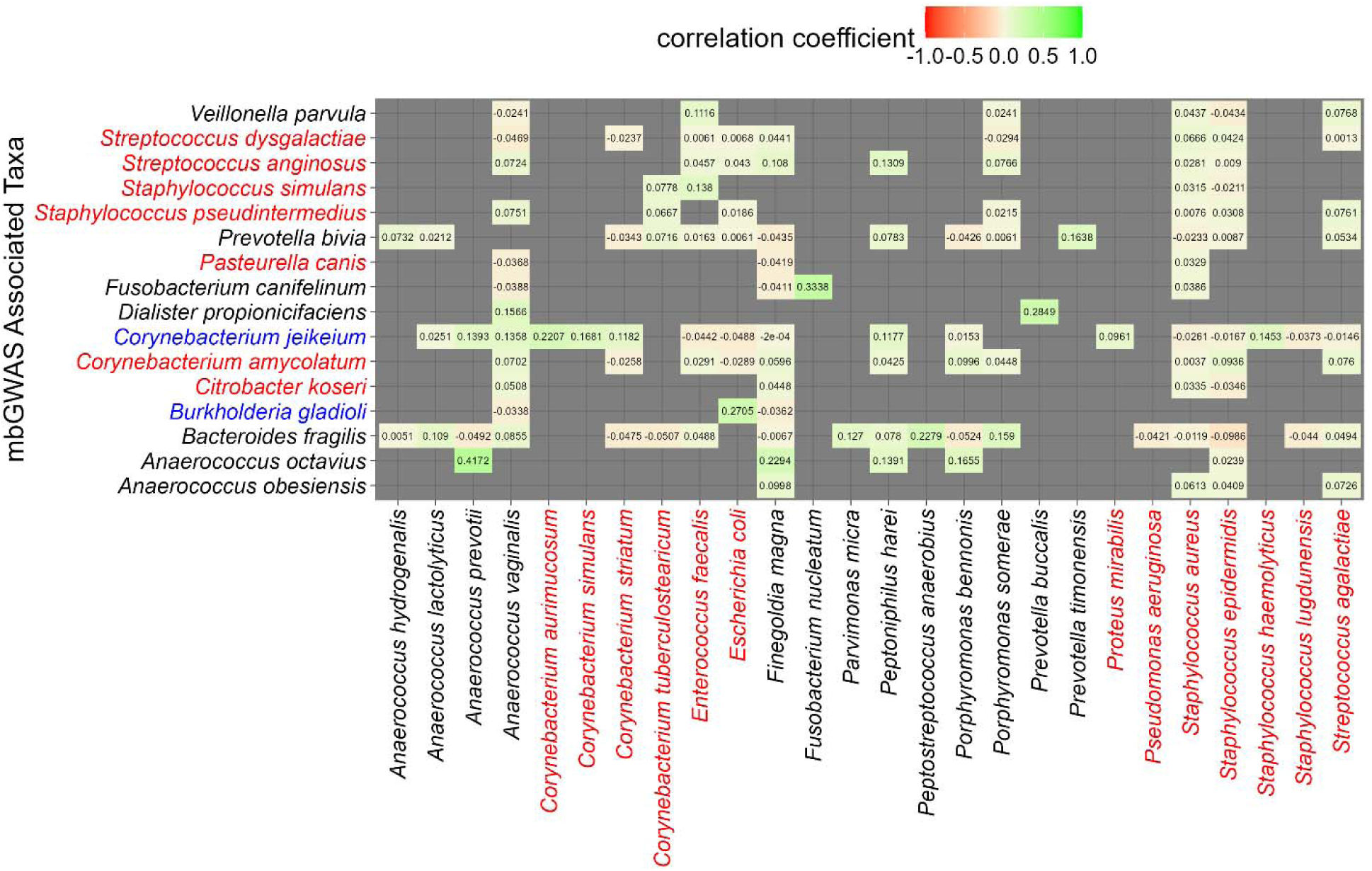
Co-abundance of associated species to all wound species. Blue font represents aerobic species, red font represents facultative anaerobes, and black font represents obligate anaerobes. All correlations shown were significant (FWER<0.05). Positive and negative co-abundances are indicated by positive and negative correlation coefficients, respectively, and are also shaded to indicate the direction of relationships.

Genomic similarity explained overall wound microbiome compositional differences Given the significant association of patient genetics to species, as well as the significant relationship among species, the relationship between candidate genetic variants and overall wound microbiome composition was tested. We employed a multiple regression to assess if the Bray-Curtis beta diversity metric was a function of genomic Euclidean distance accounting for covariates (age, sex, presence of clinical history of diabetes mellitus, and eigenvectors 1 and 2).

Considering all candidate variants in the Euclidean distance calculation resulted in no relationship (p>0.05). However, a follow up model including only variants for each region with the smallest p-values (i.e., peak variants, 27 variants across 25 regions) was conducted and found to be significant (p=0.037, intercept=0.892, β=0.009, R^2^=0.0014). Figure 6 illustrates the positive relationship between beta diversity and genomic distance of peak variants.

**Figure 6.**
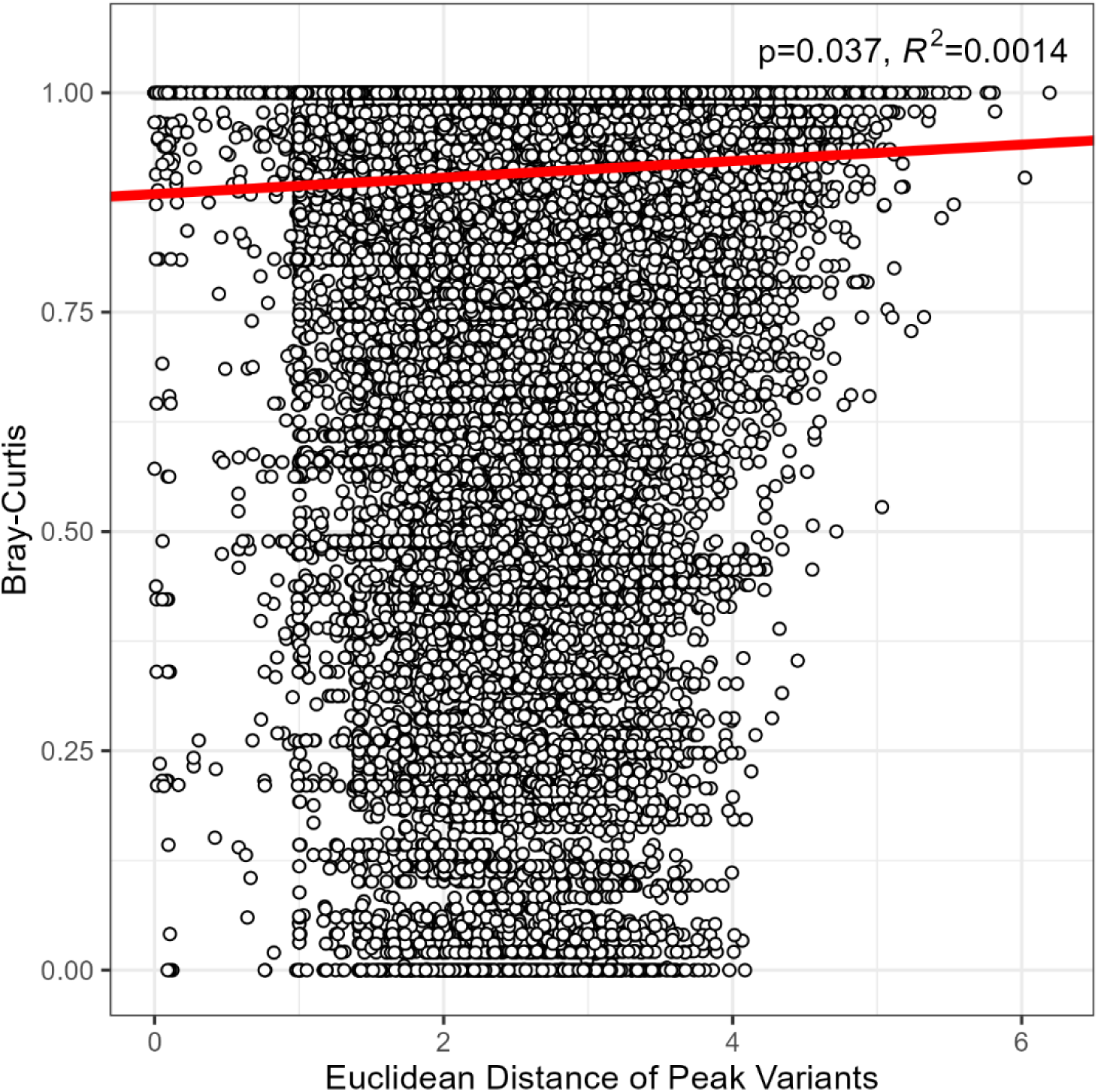
Association of microbiome composition to genomic distance. Wound microbiome compositional differences (Bray-Curtis dissimilarity) significantly increased as the genetic distance (Euclidean distance of peak variants) among patients increased (p=0.037, intercept=0.892, β=0.009, R^2^=0.0014). Analysis controlled for covariates included age, sex, history of diabetic diagnosis, and eigenvectors summarizing genetic ancestry.

## DISCUSSION

Elucidating predictors of stalled wounds is important for diagnosis and clinical management. Here, with the goal of expanding the perspective of how patient genetics relates to wound microbiomes, we conducted an mbGWAS among 509 patients from NGS wound microbiome profiles at initial clinical visit. Results support that patient genomic differences influencing the abundance of bacterial species in wounds may be common. Specifically, 193 candidate loci distributed across 25 distinct genomic regions were repeatably associated to 16 bacterial species. Notably, none of the variants associated herein have been previously implicated in skin, wounds, or infection, potentially reflecting the broad knowledge gap in our current understanding of mechanisms of human-microbe interactions.

Compared to culture-dependent identification, DNA sequencing provides improved sensitivity and specificity to accurately identify multiple bacterial species, especially anaerobes.^52^ Whereas the contribution to bioburden by uncommon or low abundance species uniquely reported by NGS is still uncertain, it is clear from the current work that species resolved by NGS are related to patient genomics, and thus are not to be considered irrelevant. Recent work suggests that finer taxonomic or genetic delineation yields more associations to the human genome as compared to higher levels.^53^ A species-level analysis was used here for translation from bedside-to-bench as species relative abundances were directly supplied to providers by the clinical diagnostic lab (MicroGenDX, Lubbock, Texas), however it is possible that analysis based on a different taxonomic level (e.g., genus or strain-level) or based on functional gene content would lead to still novel genetic associations.

Overall, results suggest that specific regions within the human genome influence the wound microenvironment in distinctive ways that are relevant to colonizing microbes. The association between distinct genomic regions and bacterial taxa was illustrated in Figure 2. Given that species were commonly found associated to multiple genomic regions, or at least multiple variants, their presence in the wound appears to have polygenic inheritance. Here, initial estimates of heritability are provided by measuring the additive effects of variants associated to each species. By comparison to all candidate variants for each species, only a subset of variants (e.g., predictor variants, Figure 3a) explained unique components of the variation of species relative abundances. For species associated with multiple candidate loci, the refined models retained predictor variants from multiple regions that additively contribute to inheritance. For example, *C. koseri* was associated with 6 variants in 3 genomic regions. In its final model, one variant from each region was retained with an R^2^ of 12.9%. The retention of these variants supports they may be causal, and all are linked to genes that may indicate the mechanistic basis: rs6752001 is an intronic variant within transient receptor potential melastatin family member 8 (*TRPM8*), a non-selective cation channel activated by factors like temperature and pressure, and has been investigated in wound closure^54,55^; rs10459902 is downstream of NLR family pyrin domain containing 1 (*NLRP1*), a mediator of inflammation largely expressed in epithelial cells, and has been implicated in skin disorders including autoimmunity and cancer^56^; lastly, rs78568090 is downstream of small nucleolar RNA host gene 5 (*SNHG5*) which has been implicated in cell migration of multiple cancers.^57-59^

In contrast to independent variants from multiple regions having additive influence, additive effects of multiple variants from a single region were also observed. This was highlighted with *S. dysgalactiae* which only associated to one genomic region. Of its candidate variants, 14% (6/42) explain 20% of variance in *S. dysgalactiae* relative abundance (Figure 3). Multiple variants from the same region each significantly contributing to this heritability estimate supports that more than one variant, and potentially more than one gene, in this region causally influences *S. dysgalactiae*. Despite this high effect size, no functional overrepresentation of this taxa or its region was observed; however, the kinase PIKfyve was the primary implicated gene for QTLs of this region, which was linked to expression differences in wound relevant tissues including tibial artery and nerve, gastrocnemius muscle, subcutaneous adipose (sampled from lower limb) and cultured primary fibroblasts from lower leg (Supplementary Data 5). PIKfyve has been shown to be important for wound healing via cell migration and proliferation of multiple cell types.^60,61^ Models for other species include their own unique combinations of predictor variants, distributions across genomic regions, and related functional enrichment. It also seems unlikely that heritability estimates for species have plateaued, but that future studies from diverse cohorts will reveal additional uniquely important variants.

In addition to false positive protection afforded by a two-cohort design, the relevance of associations was also supported by functional overrepresentation analysis (Figure 4). Notably, no enriched biological pathways were recovered when considering loci associated across all species collectively. Rather, overrepresented terms emerged when focused on specific species and genomic regions, a pattern that suggests species interact with the wound microenvironment in their own distinctive ways. Patterns indicate that different species can be selected by similar biological themes, but via different genes. For instance, Figure 4a indicates that ‘metabolism’, ‘transport of small molecules’, and ‘immune system’ were all enriched for multiple species although none of these species were ever associated to the same genomic region. Whereby analysis supported instances of species-level enrichment across genomic regions, region-specific analysis also suggested unique reasons why species may be selected by individual regions or genes. For example, region 3 contains many enriched terms (Figure 4b) that did not remain when pooled at the species level for *A. obesiensis*. Similarly, *S. anginosus* was associated with five genomic regions, but pooling across regions resulted in no significant overrepresentation. Rather, when considering these regions separately, a sole region (21) was overrepresented by 10 terms pertaining to ‘extracellular matrix organization.’ In general, the types of species and regional overrepresentations emphasize the diversity of interactions that appear to vary depending on patient genetics.

Although most associated variants lie within intronic and intergenic regions most likely of regulatory importance,^44^ eight candidate loci were exonic. Of these, three were associated with *S. anginosus* and within the aforementioned *COL18A1-AS1* (collagen type XVIII alpha 1 chain antisense RNA 1). Collagen is a major extracellular matrix component of wound healing, serving as structural strengthening of the skin.^62^ Particularly, type XVIII collagen is found in the basement membrane of vascular and epithelial structures.^63^ The association of exonic SNPs in a collagen antisense RNA suggests that genetically variable differential regulation of *COL18A1* may alter integrity of the ECM and its necessary functions, including host cell adhesion, recruitment and signaling. Where there is disrupted ECM, microbes can evade the immune response or hijack host molecules.^64^

Enrichment analysis was based on genes related to associated loci either through proximity or previously defined as eQTLs or sQTLs based on GTEx Consortium efforts. Consistent with the results reported herein being true positive associations, QTLs were disproportionately among those estimated to contribute to tissue-specific regulation of wound-relevant tissues, which are defined as tissues that can be localized to a lower extremity wound and will likely directly contribute to healing outcomes. Still, other QTLs influence regulation at other body sites that may indirectly contribute to poor healing. For instance, among identified QTLs were those known to influence thyroid gene expression, and dysregulation of the thyroid has systemic metabolic consequences. A common clinical disorder is hypothyroidism, which is associated with systemic thyroid hormone deficiency, which can result in slowed metabolism, reduced cell proliferation, and reduced collagen production.^65-67^ Thus, it is plausible to summarize both wound site-specific and systemic consequences of candidate variant effects.

Of the 68 bacterial species tested, the 16 repeatedly associated species to the human genome tended to occur at a low average relative abundance (range = 1.9-10.0%) and occurrence (range = 6-19 patients), suggesting a genetic predisposition to less common wound species. While these species tend to be less common, overall, 27.1% of patients in this study had at least one of these species detected in their wound. In addition, staphylococci and *Bacteroides* were reported to be genetically linked to skin microbiome,^53^ as is in concordance with observations in this study. It is unclear from current results why conventional wound species were not associated, but similar studies of the nasal microbiome also demonstrate limited heritability of *S. aureus*, despite being known for nasal colonization and carriage.^68,69^ That said, many common species were significantly correlated through co-abundance analysis to genetically associated species. Whereas co-abundance does not directly confirm species-species interactions, it is consistent with community assembly processes. Furthermore, it is not possible to comment from current results if there is an order to species colonization. One possibility is that genetically associated species colonize first facilitating habitat for the other species. In any case*, S. aureus*, *S. epidermidis*, *A. vaginalis*, and *F. magna* stood out by having significant co-abundance relationships to all but one (*B. gladioli*) genetically associated species.

The findings of this study should be interpreted with the following limitations. This was a cross-sectional evaluation of the wound microbiome, whereby only the earliest assessment to a specialty wound clinic was considered. Methodological approaches to include longitudinal sampling (i.e., multiple microbiomes per wound) into GWAS analysis will be undertaken in future work. Also, whereas this study included one wound per patient, patients may suffer multiple wounds, and modelling multiple phenotypes per patient could be important to further understand patient genetic effect. Samples were also collected from one single-physician outpatient clinic, so potential center-specific effects such as treatment strategy were not considered. Lastly, all wounds were sequenced for microbial profiling in response to clinical non-healing; that is, communities with a positive healing trajectory were not available. Therefore, future work will not only include expansion of patient and sample collection into a longitudinal context but will also include multicenter recruitment.

Our long-term goal is to empower precision medicine by developing a clinical genetic tool that contributes to decision making of wound care management and as a risk assessment of wound prognosis. In the future, patients presenting with non-healing, recalcitrant wounds may benefit from assessment of genetic predisposition to poor healing, which may be due to either dysregulated host healing, predisposition to wound pathogens, or both. Likewise, clinical assessment utilizing a genetic test at preventative care visits establishes the urgency of referral to wound care specialists prior to the need for intervention. Current results establish that patients with chronic wounds differ in their wound microbiome composition in part due to individual genetic uniqueness.

## ACKNOWLEDGEMENTS

We thank MicroGenDX and CEO Rick Martin for their support of this work. We thank the staff at the Southwest Regional Wound Care Center for their commitment to excellence in wound care, scientific collaboration, and logistical support. This work was supported by NIH award R15GM141973-01.

## DATA AVAILABILITY

The datasets generated and analyzed during the current study are available in the Gene Expression Omnibus (GEO) repository with the accession number: GSE276944.

## SUPPLEMENTARY INFORMATION

Supplementary Data 1a. Manhattan plots

Supplementary Data 1b. QQ-plots

Supplementary Data 1c. Genomic inflation factor

Supplementary Data 2. PLINK results

Supplementary Data 3. LDlink results

Supplementary Data 4. Variant database search results

Supplementary Data 5. Functional annotation of candidate variants

Supplementary Data 6. Overrepresentation analysis results

