## Supplementary Data 1 for "Functionally enriched human polymorphisms associate to species in the chronic wound microbiome"

Supplementary Data 1. (a) Manhattan plots, (b) QQ-plots, and (c) genomic inflation factor (λ) of all mbGWAS across 68 bacterial species. Cohort 1 corresponded to the original exploratory cohort and cohort 2 was the *inversus* exploratory cohort (i.e., the original replication cohort). In Manhattan plots, the red line represents the significant threshold ($\alpha=8.08\text{x}10^{-9}$), and blue line represents the suggestive threshold ($\alpha=8.08\text{x}10^{-7}$).

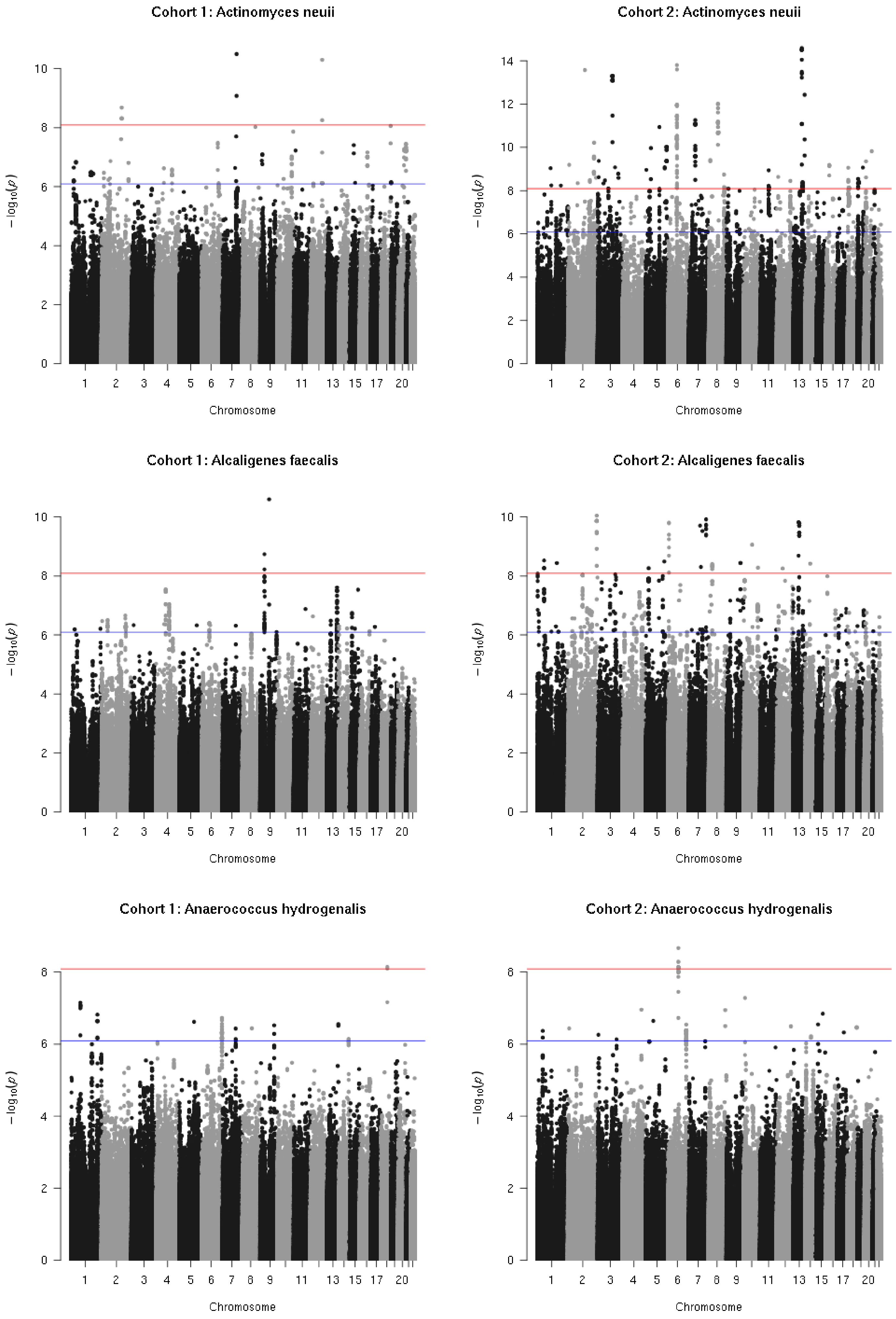
Cohort 1: Actinomyces neuii
Cohort 2: Actinomyces neuii
Cohort 1: Alcaligenes faecalis
Cohort 2: Alcaligenes faecalis
Cohort 1: Anaerococcus hydrogenalis
Cohort 2: Anaerococcus hydrogenalis
$-\log_{10}(p)$
Chromosome
1 2 3 4 5 6 7 8 9 11 13 15 17 20

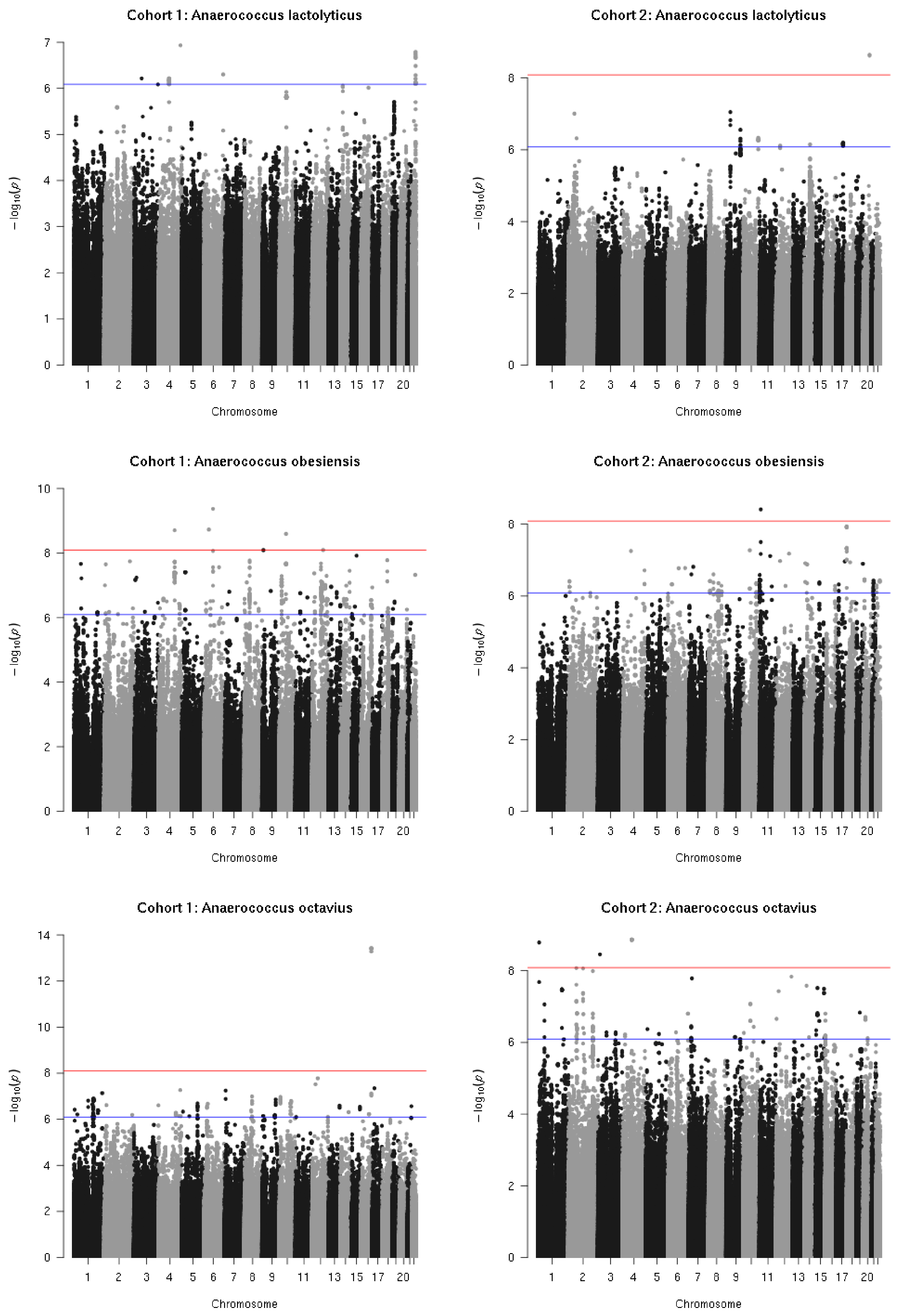
Cohort 1: Anaerococcus lactolyticus
Cohort 2: Anaerococcus lactolyticus
Cohort 1: Anaerococcus obesiensis
Cohort 2: Anaerococcus obesiensis
Cohort 1: Anaerococcus octavius
Cohort 2: Anaerococcus octavius
$-\log_{10}(p)$
Chromosome
1 2 3 4 5 6 7 8 9 11 13 15 17 20

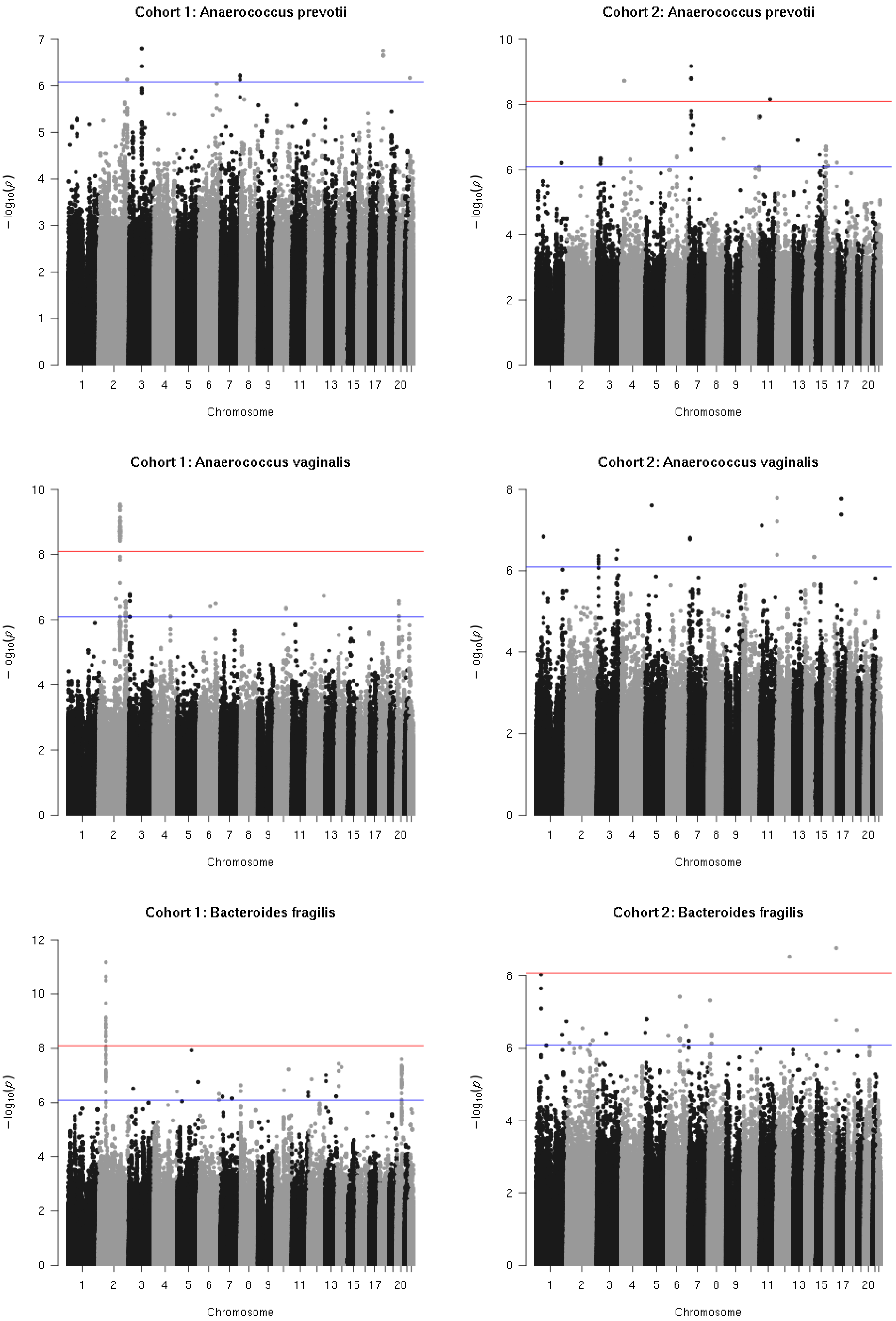
Cohort 1: Anaerococcus prevotii
Cohort 2: Anaerococcus prevotii
Cohort 1: Anaerococcus vaginalis
Cohort 2: Anaerococcus vaginalis
Cohort 1: Bacteroides fragilis
Cohort 2: Bacteroides fragilis
$-\log_{10}(p)$
Chromosome
1 2 3 4 5 6 7 8 9 11 13 15 17 20

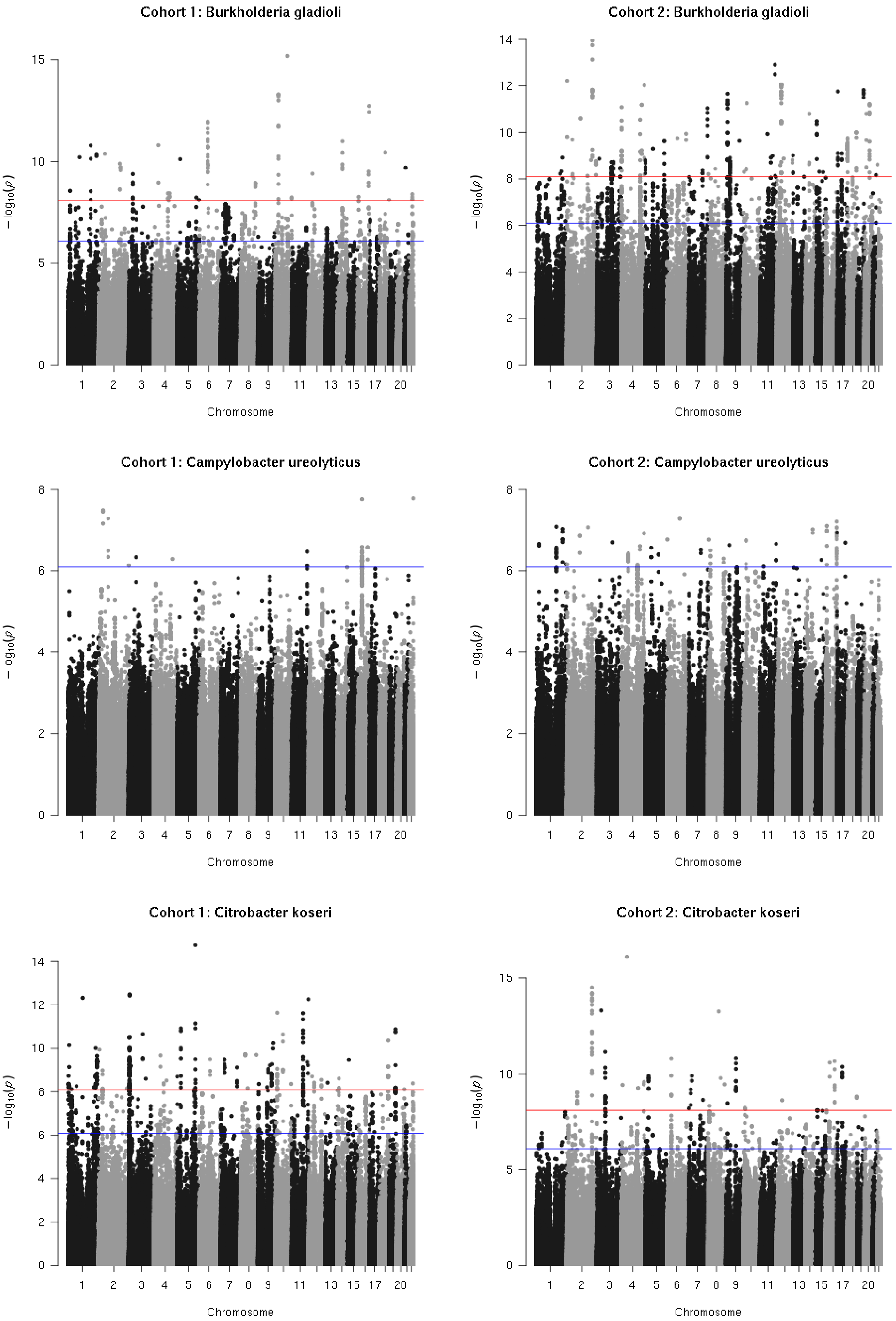
Cohort 1: Burkholderia gladioli
Cohort 2: Burkholderia gladioli
Cohort 1: Campylobacter ureolyticus
Cohort 2: Campylobacter ureolyticus
Cohort 1: Citrobacter koseri
Cohort 2: Citrobacter koseri
$-\log_{10}(p)$
Chromosome
1 2 3 4 5 6 7 8 9 11 13 15 17 20

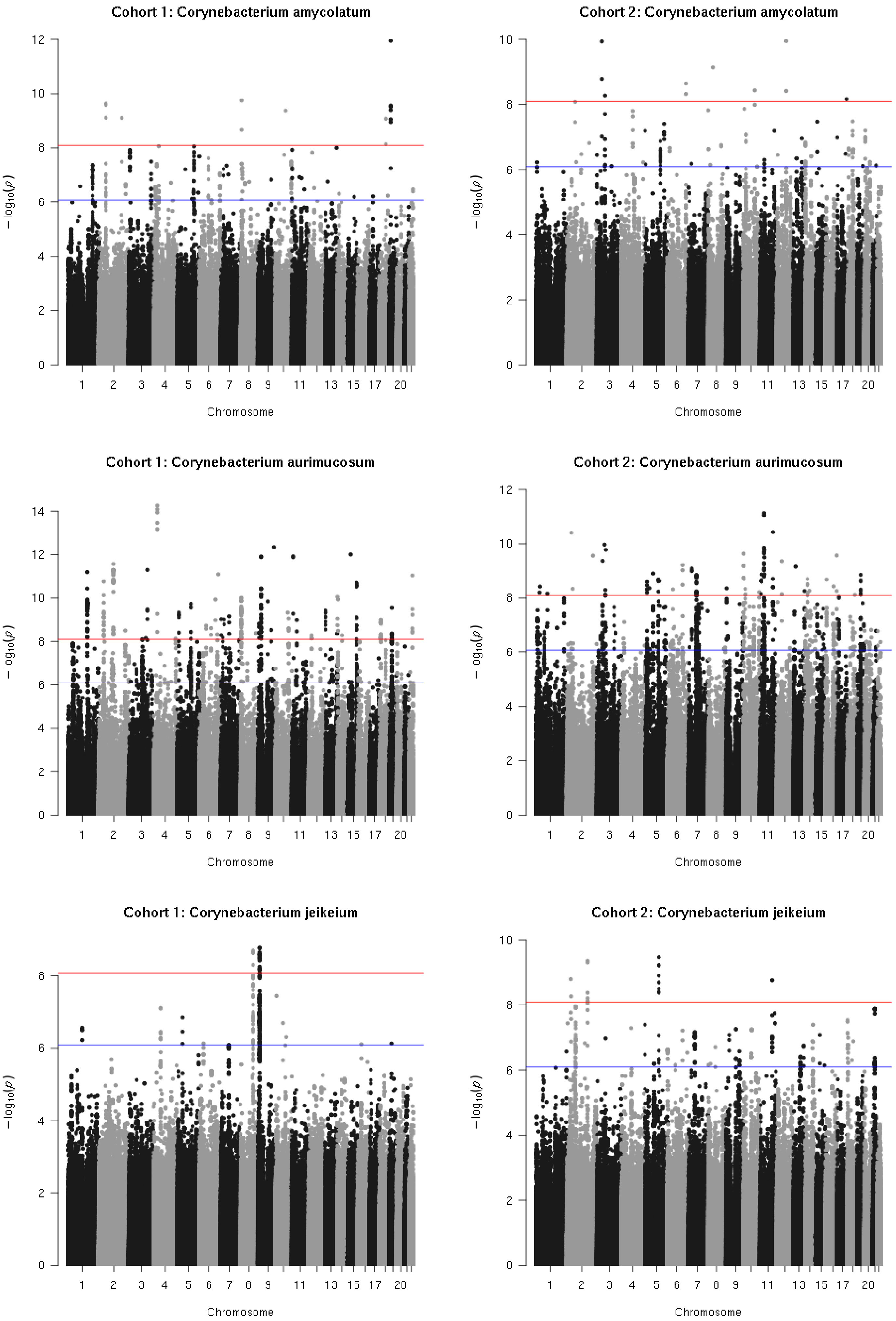
Cohort 1: Corynebacterium amycolatum
Cohort 2: Corynebacterium amycolatum
Cohort 1: Corynebacterium aurimucosum
Cohort 2: Corynebacterium aurimucosum
Cohort 1: Corynebacterium jeikeium
Cohort 2: Corynebacterium jeikeium
$-\log_{10}(p)$
Chromosome

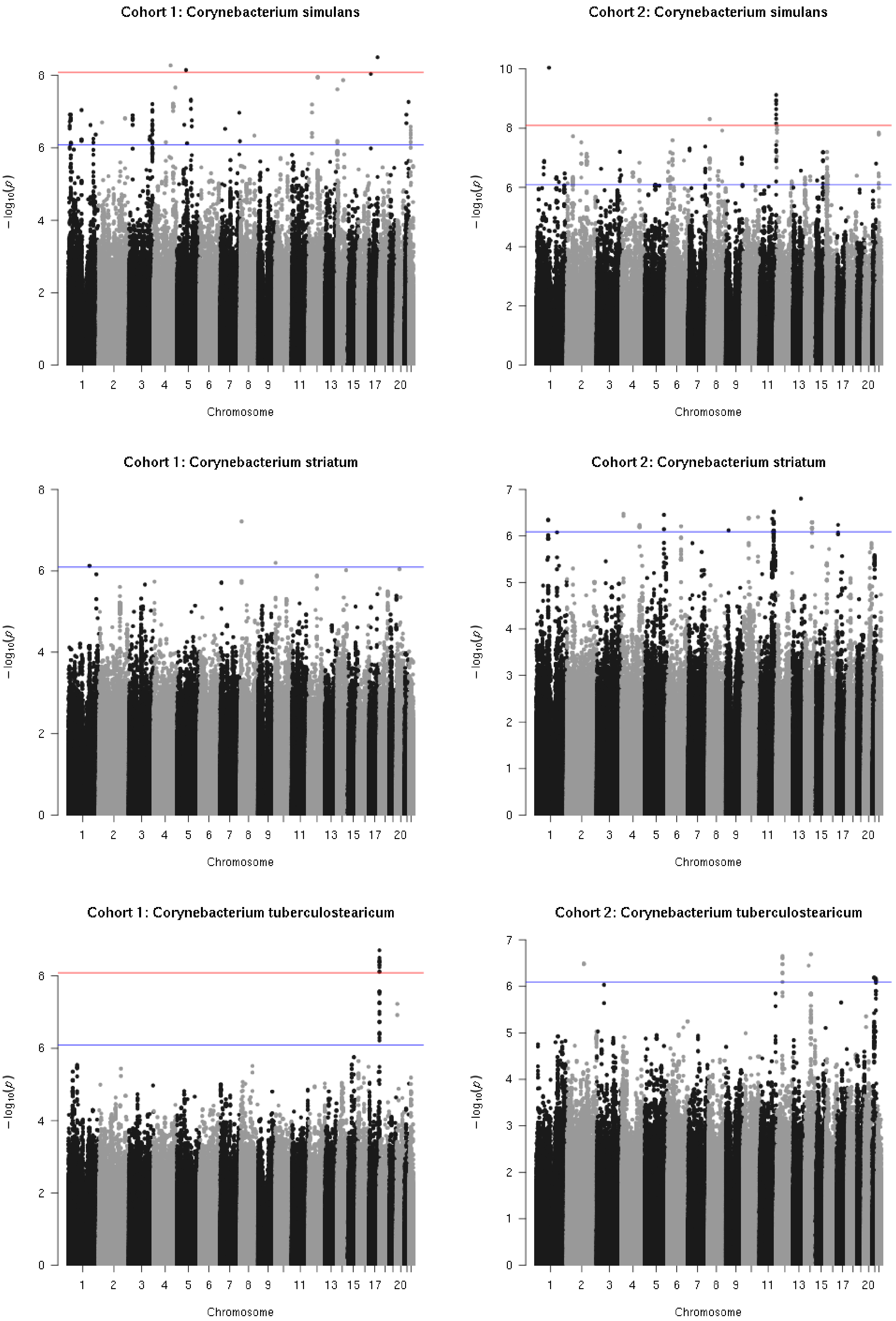
Cohort 1: Corynebacterium simulans
Cohort 2: Corynebacterium simulans
Cohort 1: Corynebacterium striatum
Cohort 2: Corynebacterium striatum
Cohort 1: Corynebacterium tuberculostearicum
Cohort 2: Corynebacterium tuberculostearicum
$-\log_{10}(p)$
Chromosome
1 2 3 4 5 6 7 8 9 11 13 15 17 20

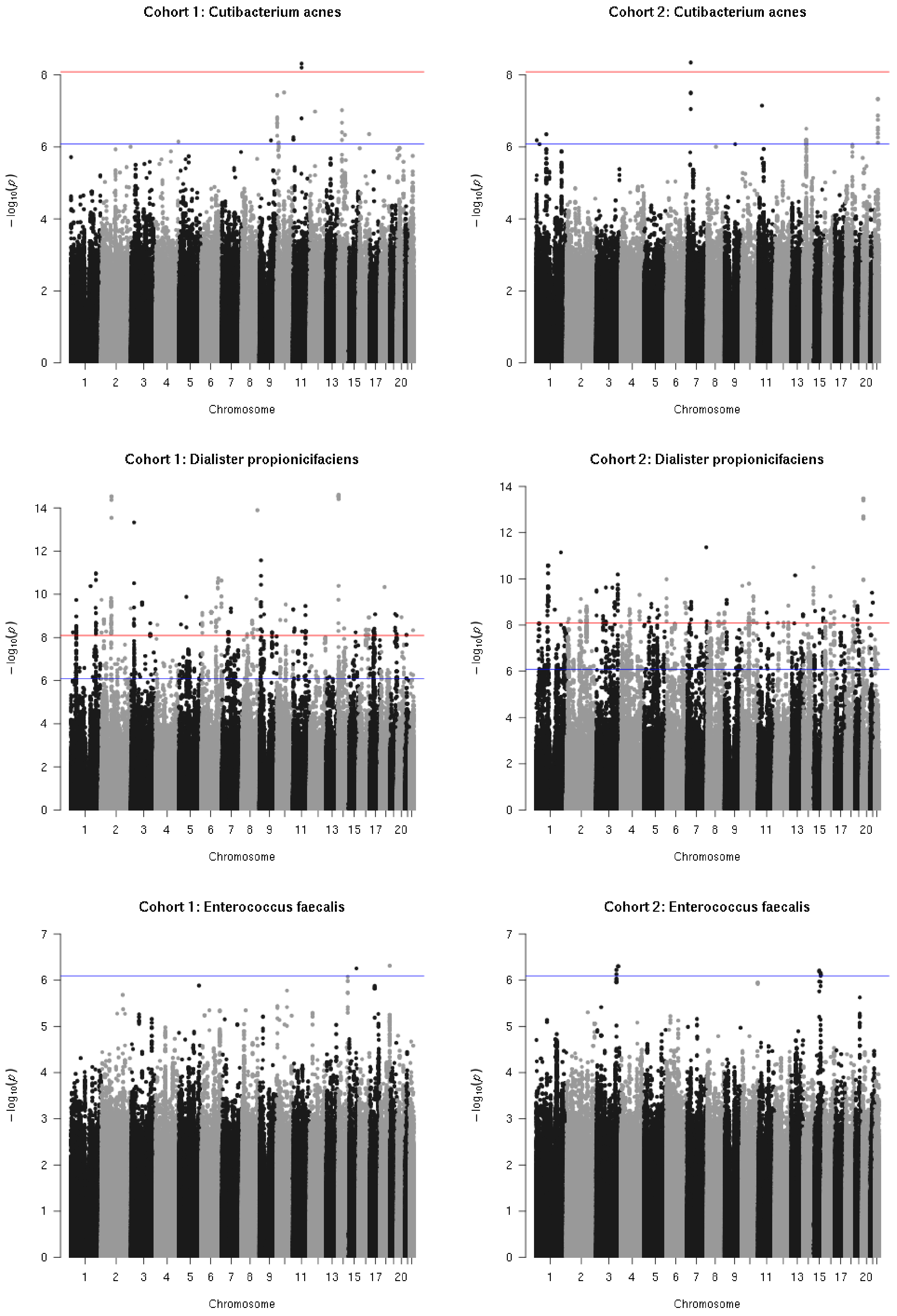
Cohort 1: Cutibacterium acnes
Cohort 2: Cutibacterium acnes
Cohort 1: Dialister propionicifaciens
Cohort 2: Dialister propionicifaciens
Cohort 1: Enterococcus faecalis
Cohort 2: Enterococcus faecalis
$-\log_{10}(p)$
Chromosome
1 2 3 4 5 6 7 8 9 11 13 15 17 20

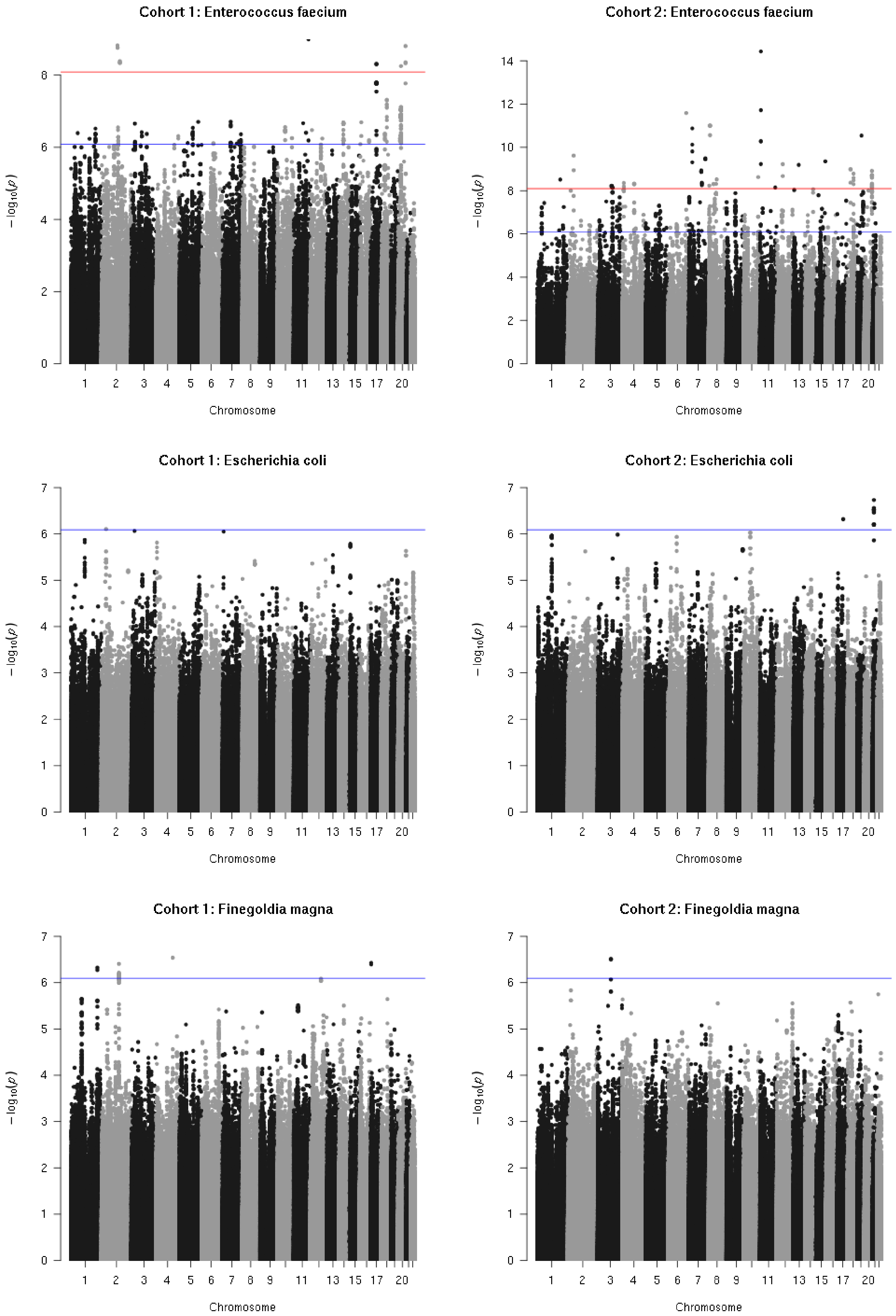
Cohort 1: Enterococcus faecium
Cohort 2: Enterococcus faecium
Cohort 1: Escherichia coli
Cohort 2: Escherichia coli
Cohort 1: Finegoldia magna
Cohort 2: Finegoldia magna
$-\log_{10}(p)$
Chromosome

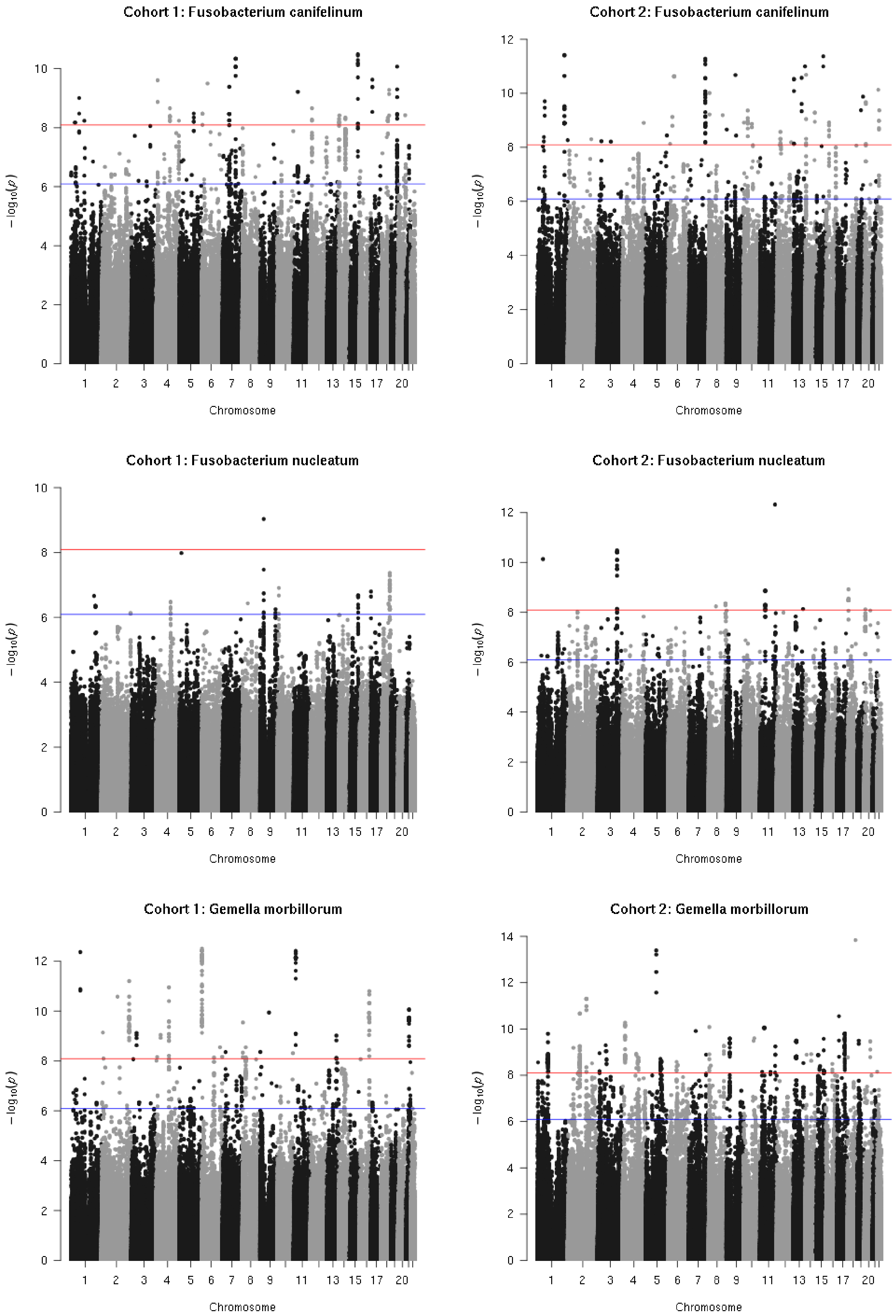

Cohort 1: Fusobacterium canifelinum
Cohort 2: Fusobacterium canifelinum
Cohort 1: Fusobacterium nucleatum
Cohort 2: Fusobacterium nucleatum
Cohort 1: Gemella morbillorum
Cohort 2: Gemella morbillorum
$-\log_{10}(p)$
Chromosome
1 2 3 4 5 6 7 8 9 11 13 15 17 20

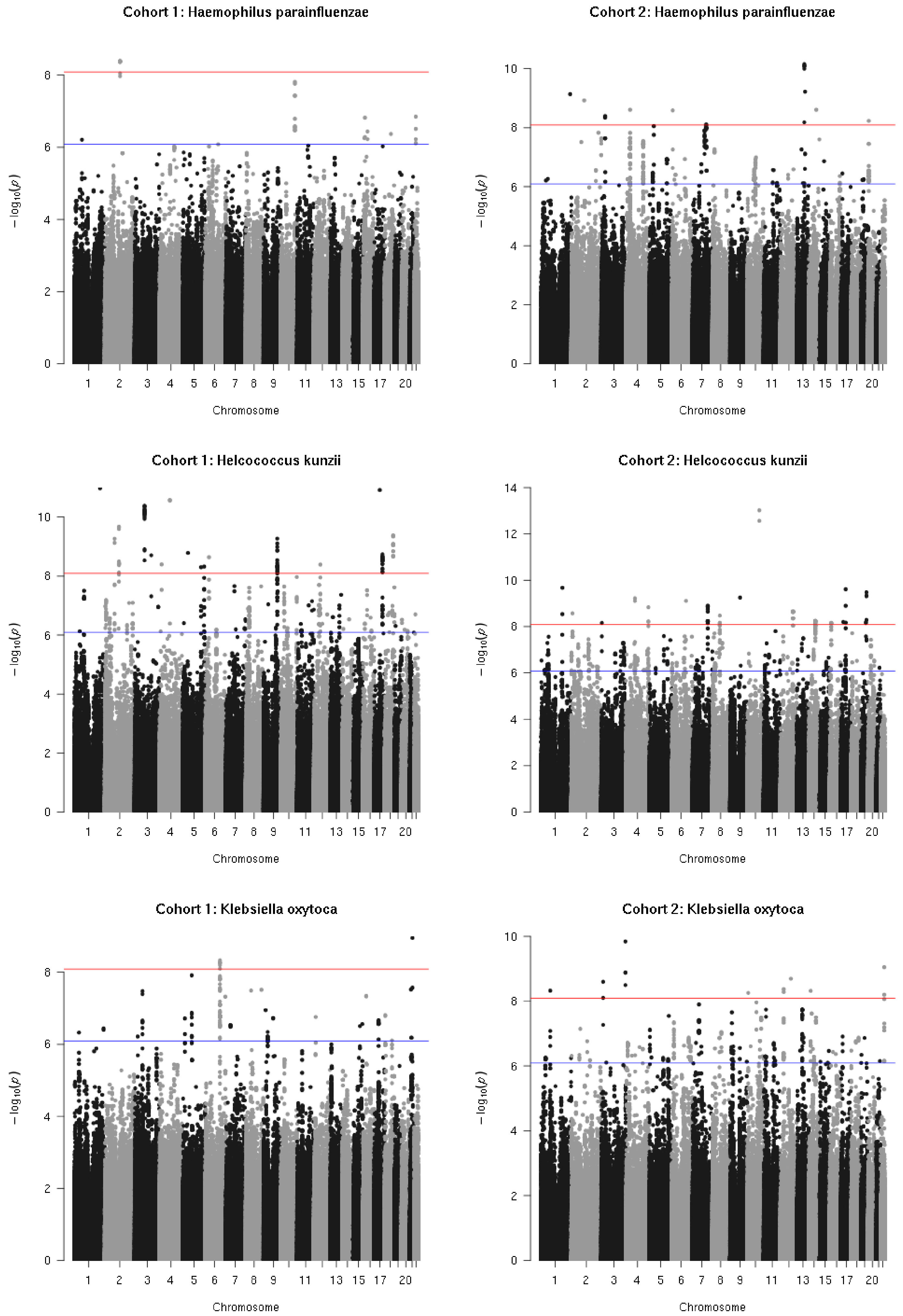
Cohort 1: Haemophilus parainfluenzae
Cohort 2: Haemophilus parainfluenzae
Cohort 1: Helcococcus kunzii
Cohort 2: Helcococcus kunzii
Cohort 1: Klebsiella oxytoca
Cohort 2: Klebsiella oxytoca
$-\log_{10}(p)$
Chromosome
1 2 3 4 5 6 7 8 9 11 13 15 17 20

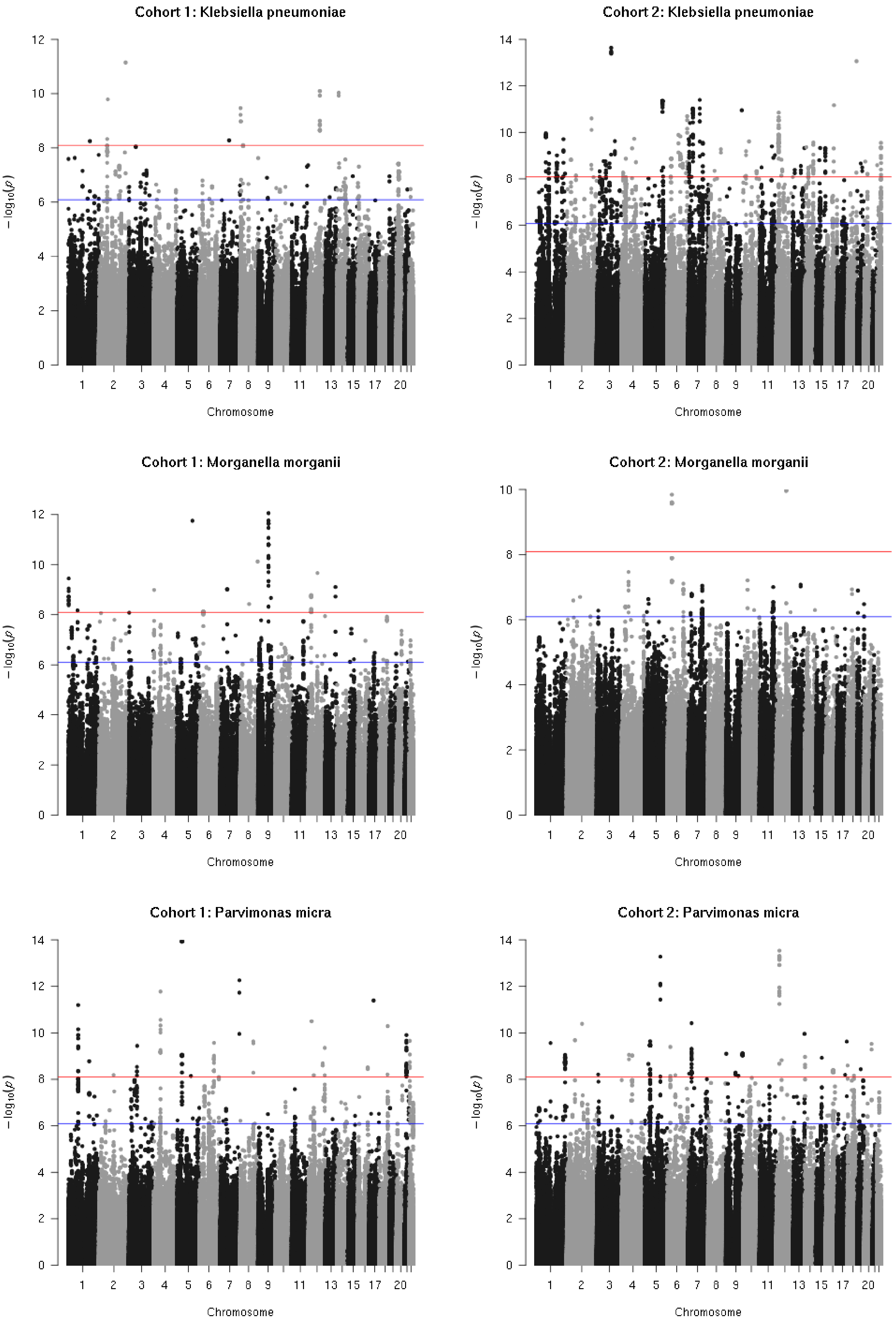

Cohort 1: Klebsiella pneumoniae
Cohort 2: Klebsiella pneumoniae
Cohort 1: Morganella morganii
Cohort 2: Morganella morganii
Cohort 1: Parvimonas micra
Cohort 2: Parvimonas micra
$-\log_{10}(p)$
Chromosome
1 2 3 4 5 6 7 8 9 11 13 15 17 20

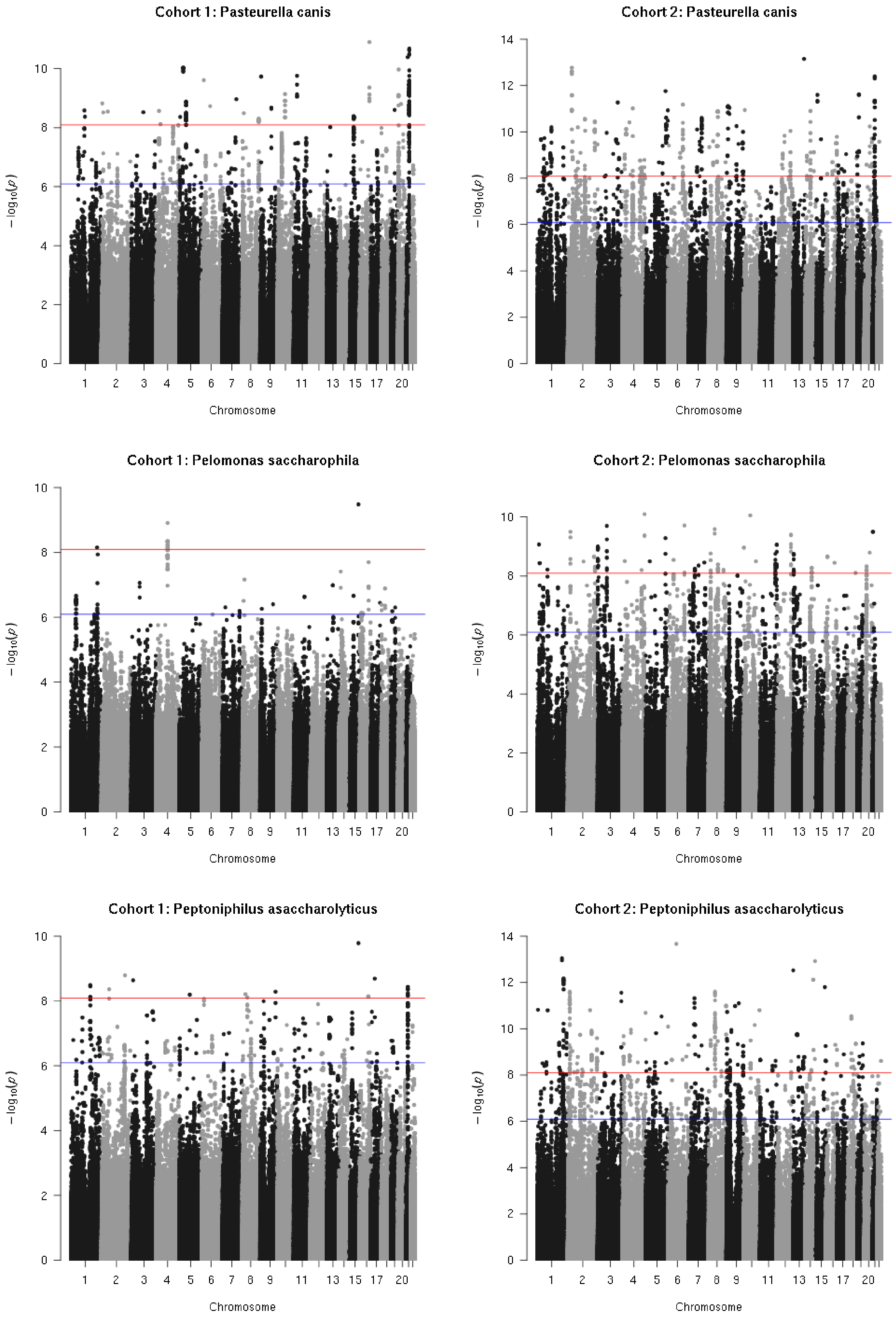
Cohort 1: Pasteurella canis
Cohort 2: Pasteurella canis
Cohort 1: Pelomonas saccharophila
Cohort 2: Pelomonas saccharophila
Cohort 1: Peptoniphilus asaccharolyticus
Cohort 2: Peptoniphilus asaccharolyticus
$-\log_{10}(p)$
Chromosome
1 2 3 4 5 6 7 8 9 11 13 15 17 20

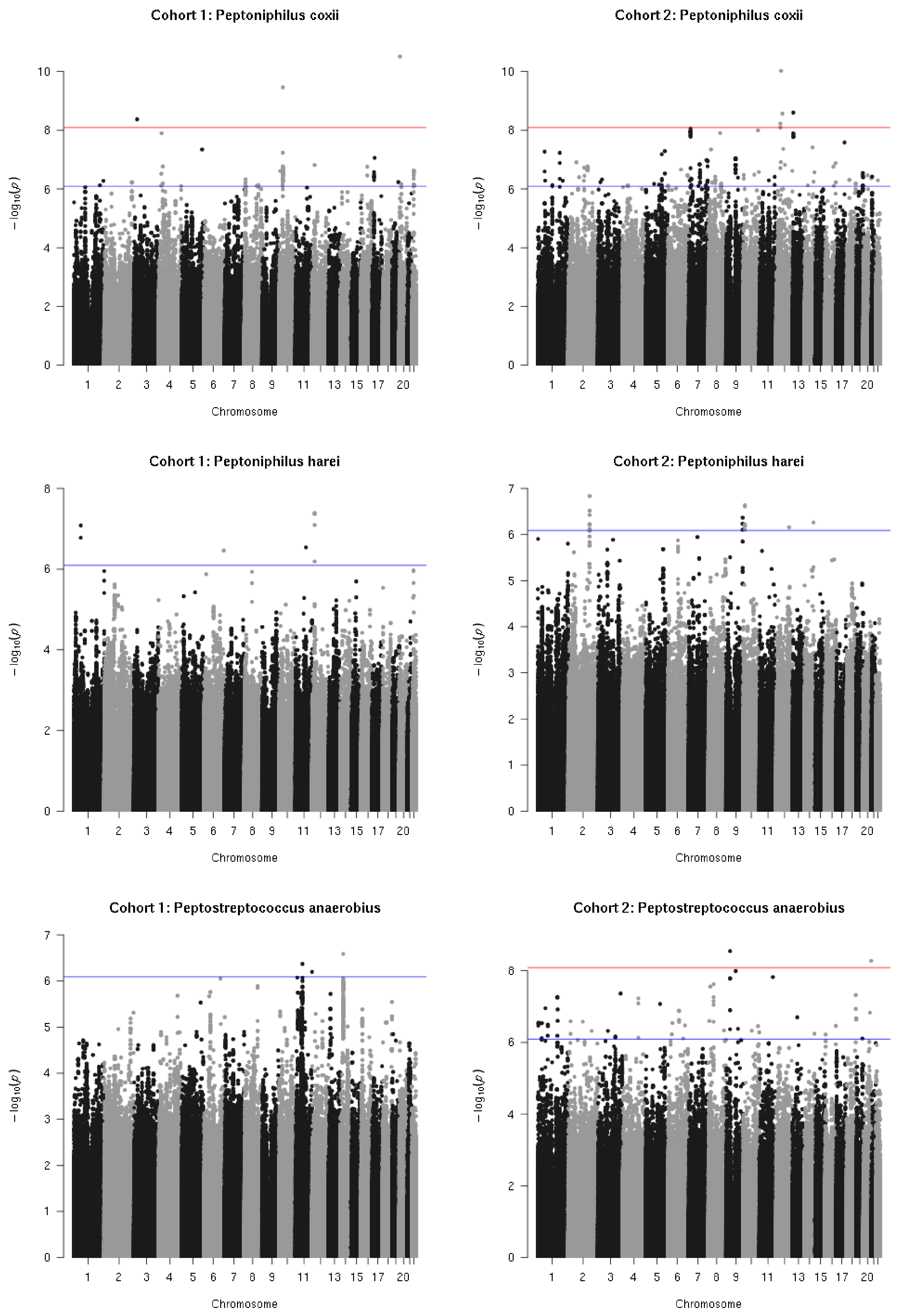
Cohort 1: Peptoniphilus coxii
Cohort 2: Peptoniphilus coxii
Cohort 1: Peptoniphilus harei
Cohort 2: Peptoniphilus harei
Cohort 1: Peptostreptococcus anaerobius
Cohort 2: Peptostreptococcus anaerobius
$-\log_{10}(p)$
Chromosome

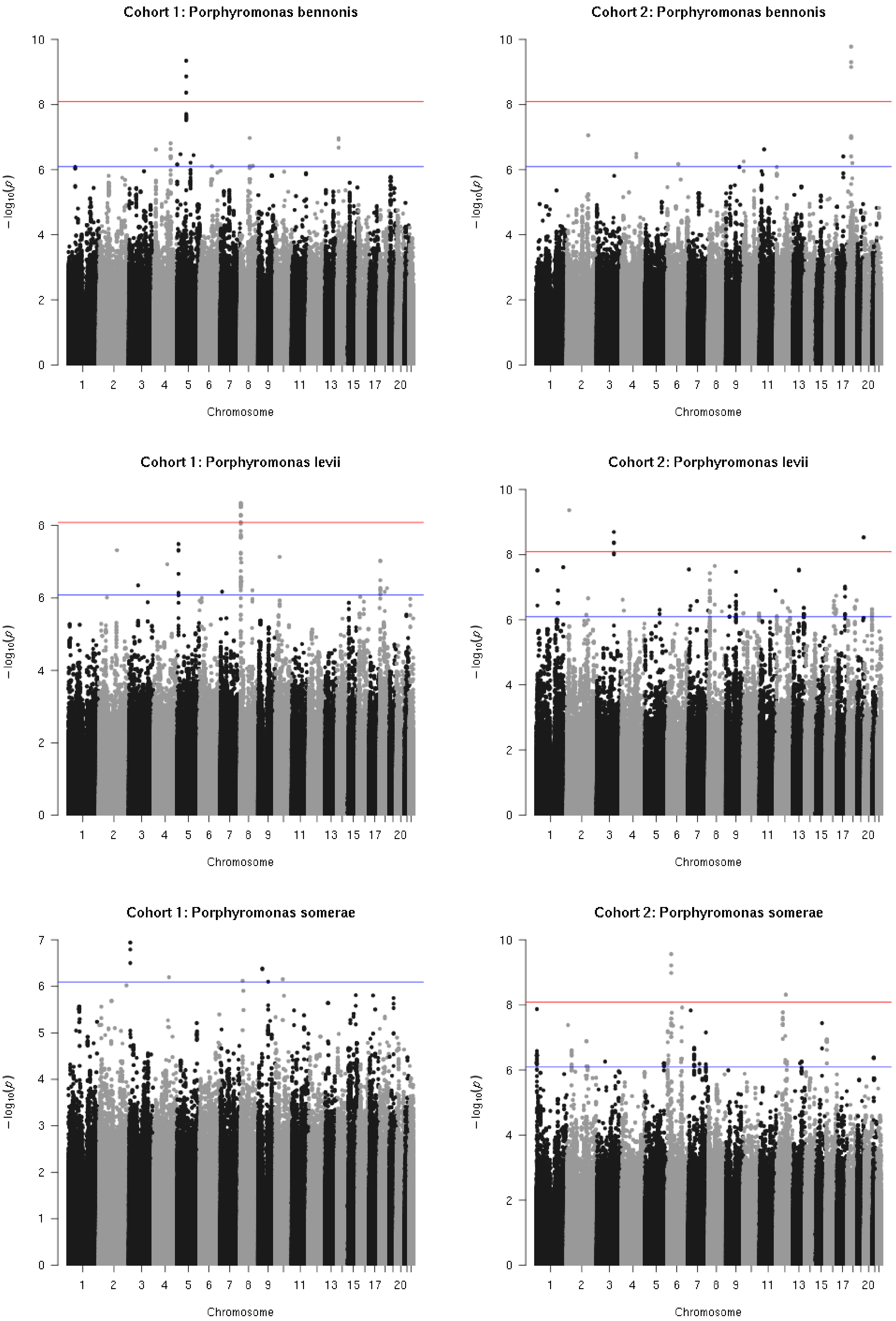
Cohort 1: Porphyromonas bennonis
Cohort 2: Porphyromonas bennonis
Cohort 1: Porphyromonas levii
Cohort 2: Porphyromonas levii
Cohort 1: Porphyromonas somerae
Cohort 2: Porphyromonas somerae
$-\log_{10}(p)$
Chromosome
1 2 3 4 5 6 7 8 9 11 13 15 17 20

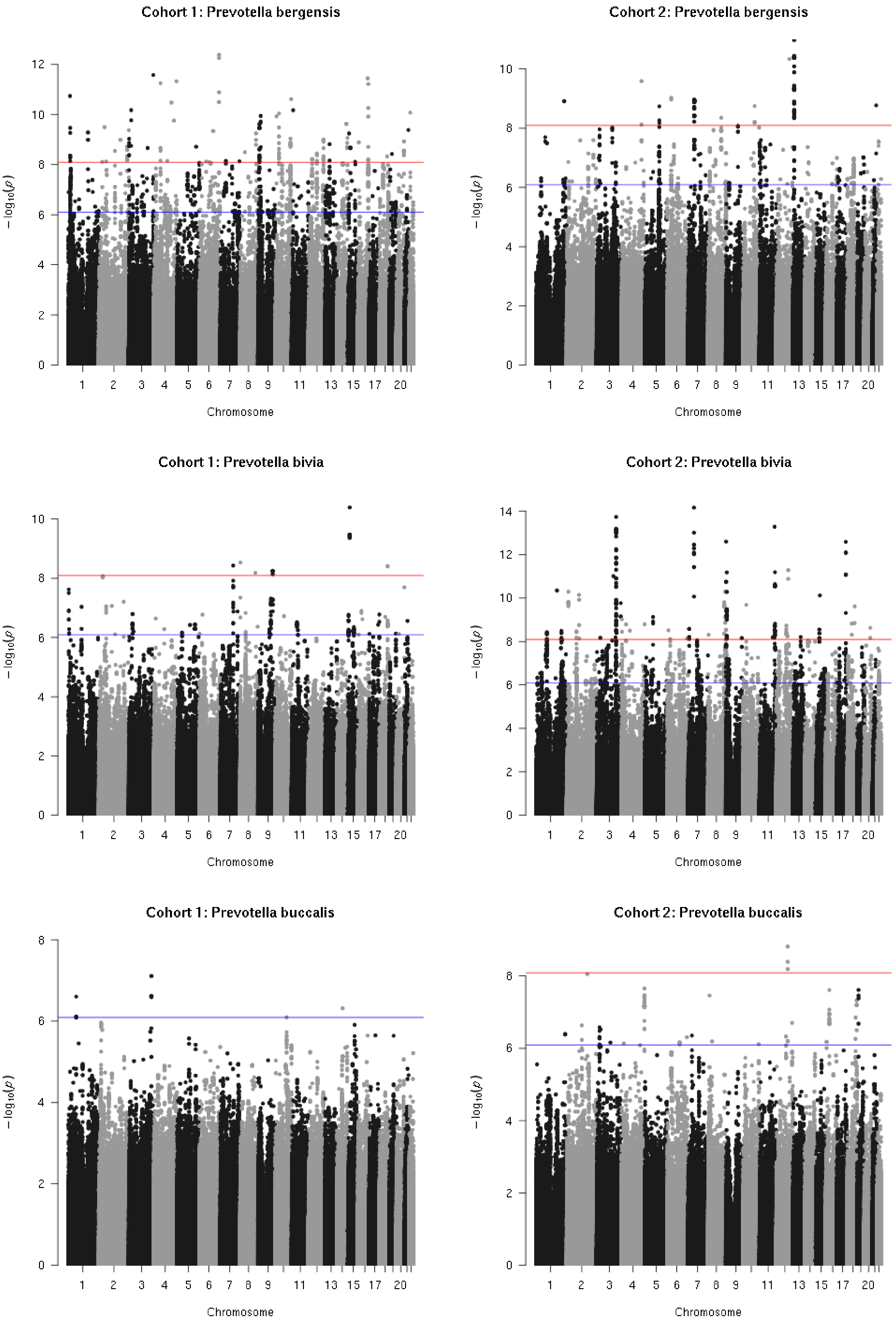
Cohort 1: Prevotella bergensis
Cohort 2: Prevotella bergensis
Cohort 1: Prevotella bivia
Cohort 2: Prevotella bivia
Cohort 1: Prevotella buccalis
Cohort 2: Prevotella buccalis
$-\log_{10}(p)$
Chromosome
1 2 3 4 5 6 7 8 9 11 13 15 17 20

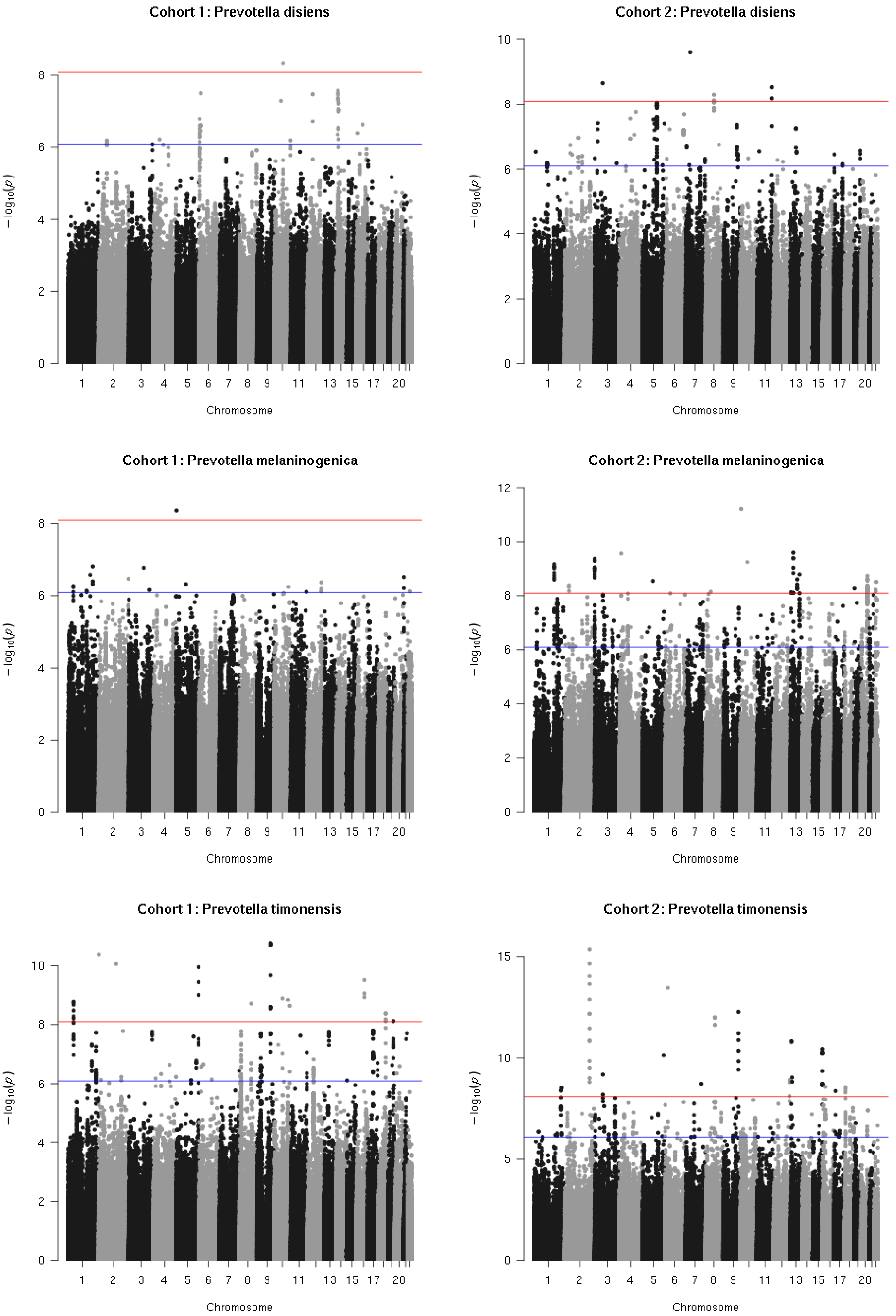
Cohort 1: Prevotella disiens
Cohort 2: Prevotella disiens
Cohort 1: Prevotella melaninogenica
Cohort 2: Prevotella melaninogenica
Cohort 1: Prevotella timonensis
Cohort 2: Prevotella timonensis
−log10(p)
Chromosome

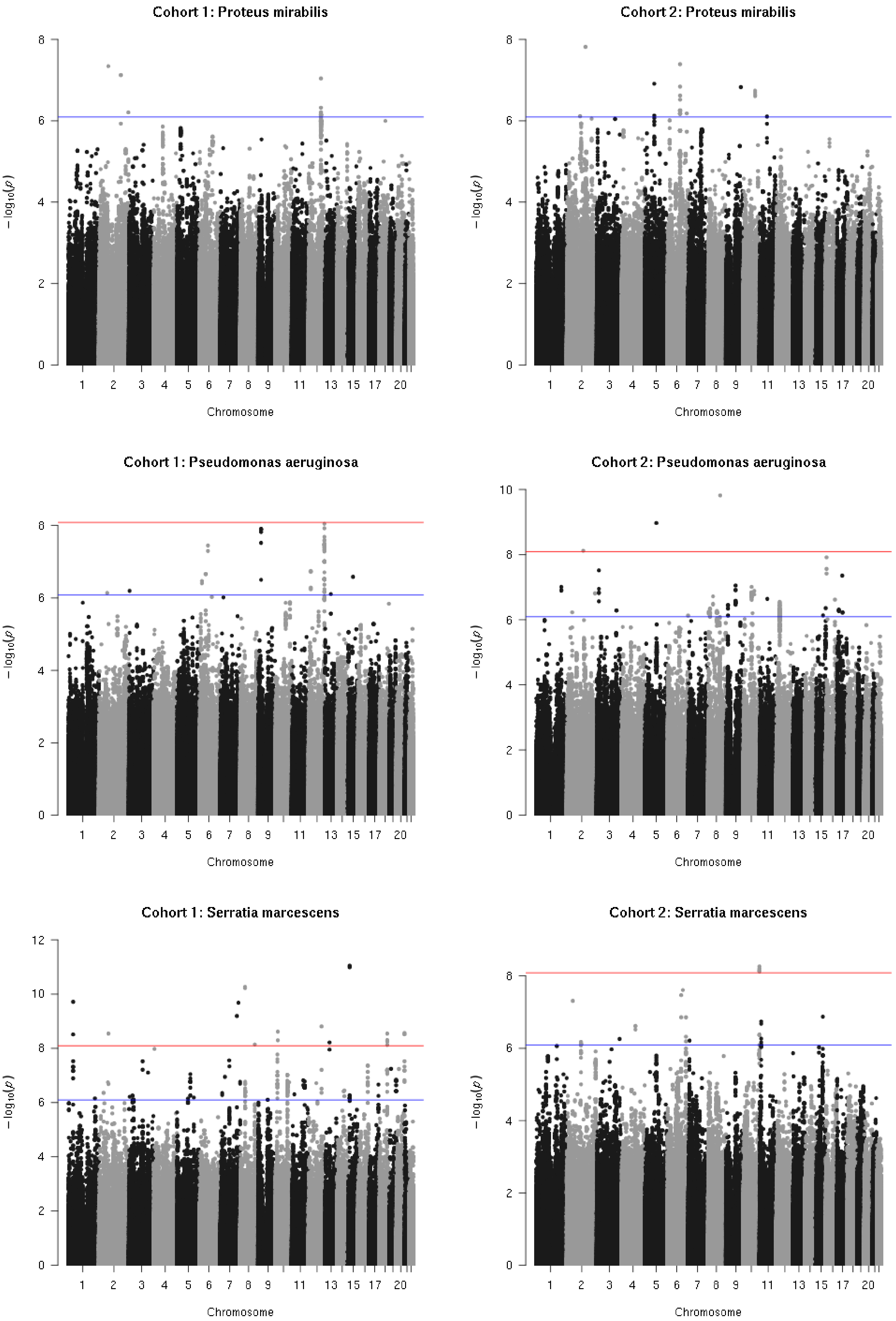
Cohort 1: Proteus mirabilis
Cohort 2: Proteus mirabilis
Cohort 1: Pseudomonas aeruginosa
Cohort 2: Pseudomonas aeruginosa
Cohort 1: Serratia marcescens
Cohort 2: Serratia marcescens
$-\log_{10}(p)$
Chromosome

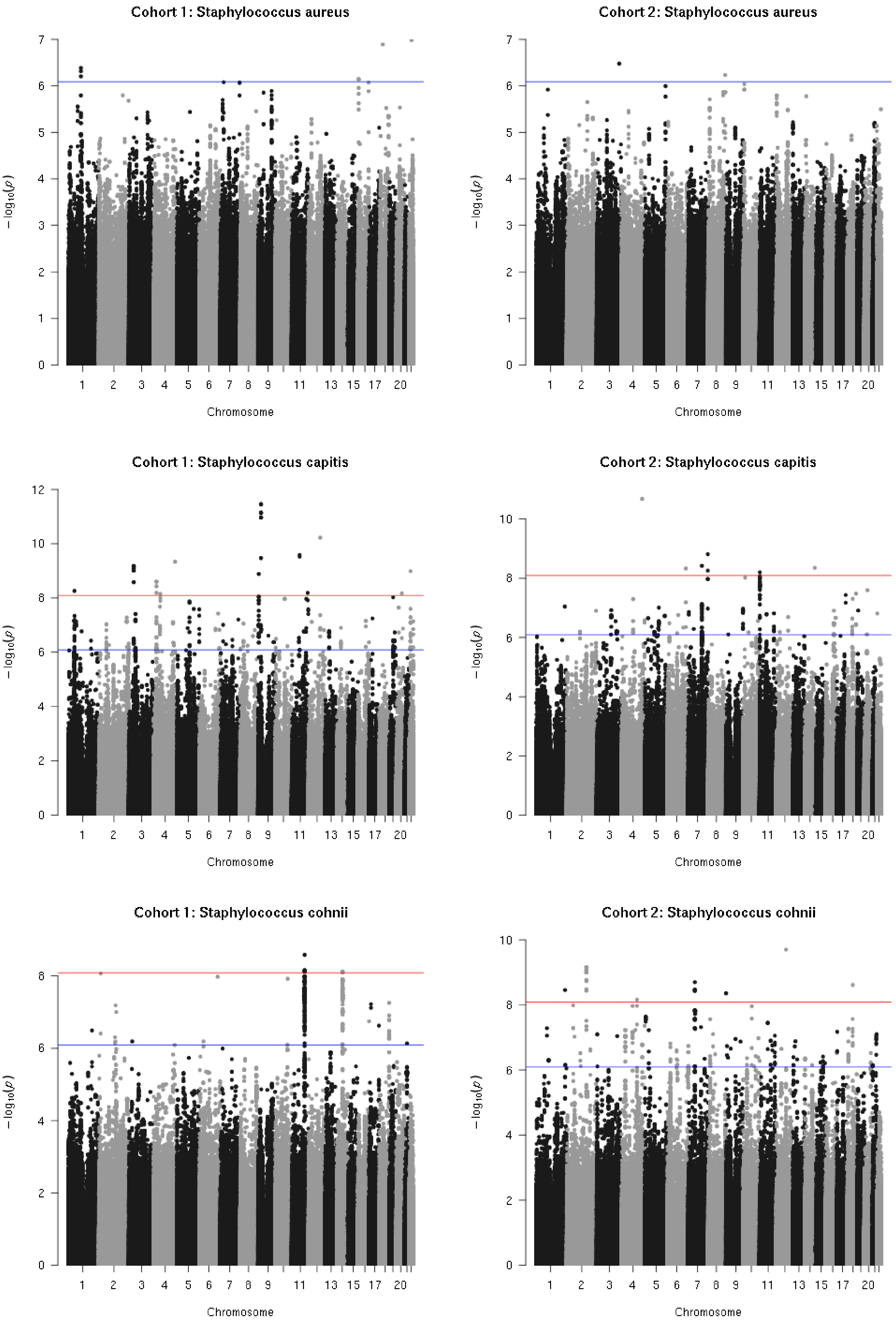
Cohort 1: Staphylococcus aureus
Cohort 2: Staphylococcus aureus
Cohort 1: Staphylococcus capitis
Cohort 2: Staphylococcus capitis
Cohort 1: Staphylococcus cohnii
Cohort 2: Staphylococcus cohnii
$-\log_{10}(p)$
Chromosome
1 2 3 4 5 6 7 8 9 11 13 15 17 20

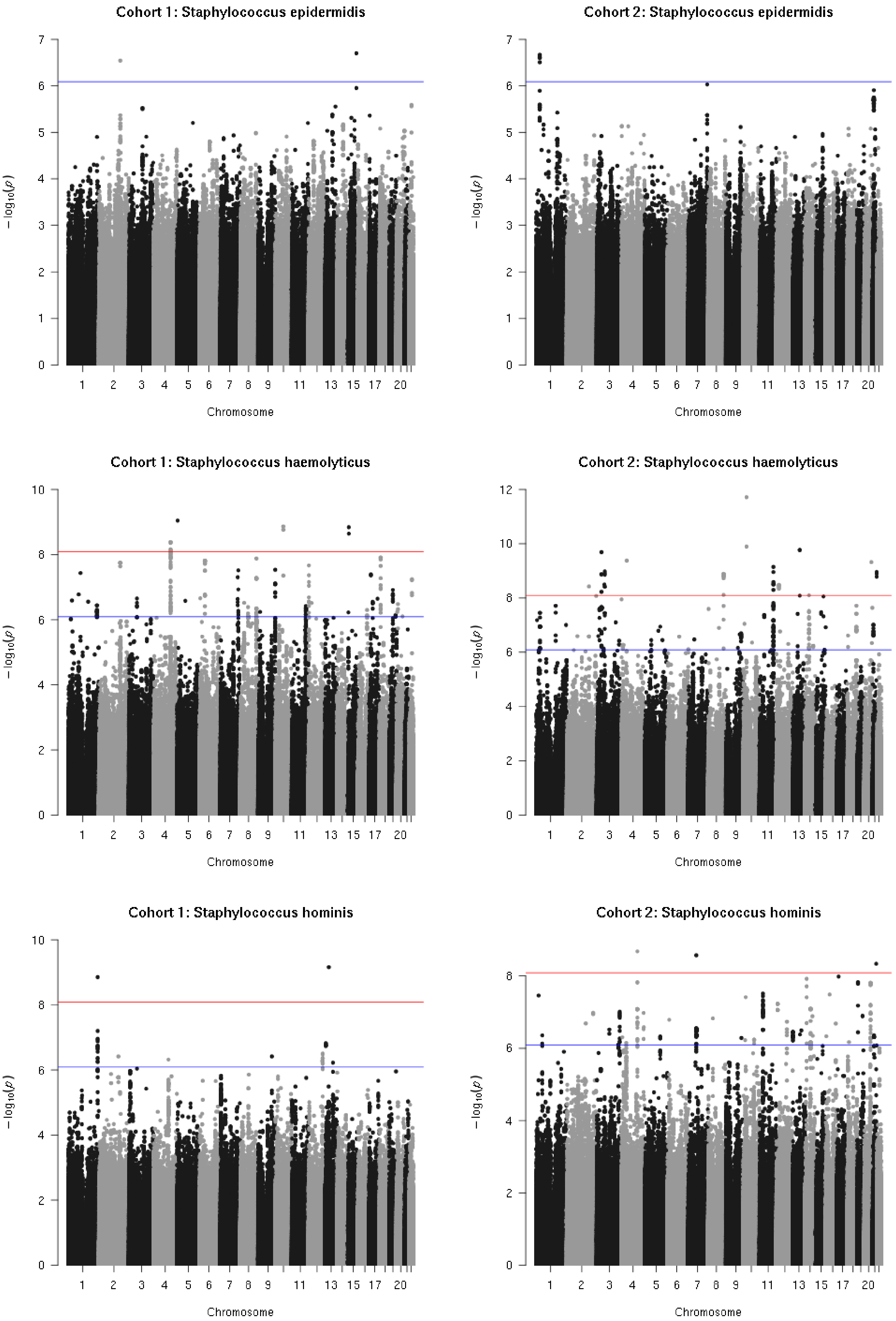
Cohort 1: Staphylococcus epidermidis
Cohort 2: Staphylococcus epidermidis
Cohort 1: Staphylococcus haemolyticus
Cohort 2: Staphylococcus haemolyticus
Cohort 1: Staphylococcus hominis
Cohort 2: Staphylococcus hominis
$-\log_{10}(p)$
Chromosome
1
2
3
4
5
6
7
8
9
11
13
15
17
20

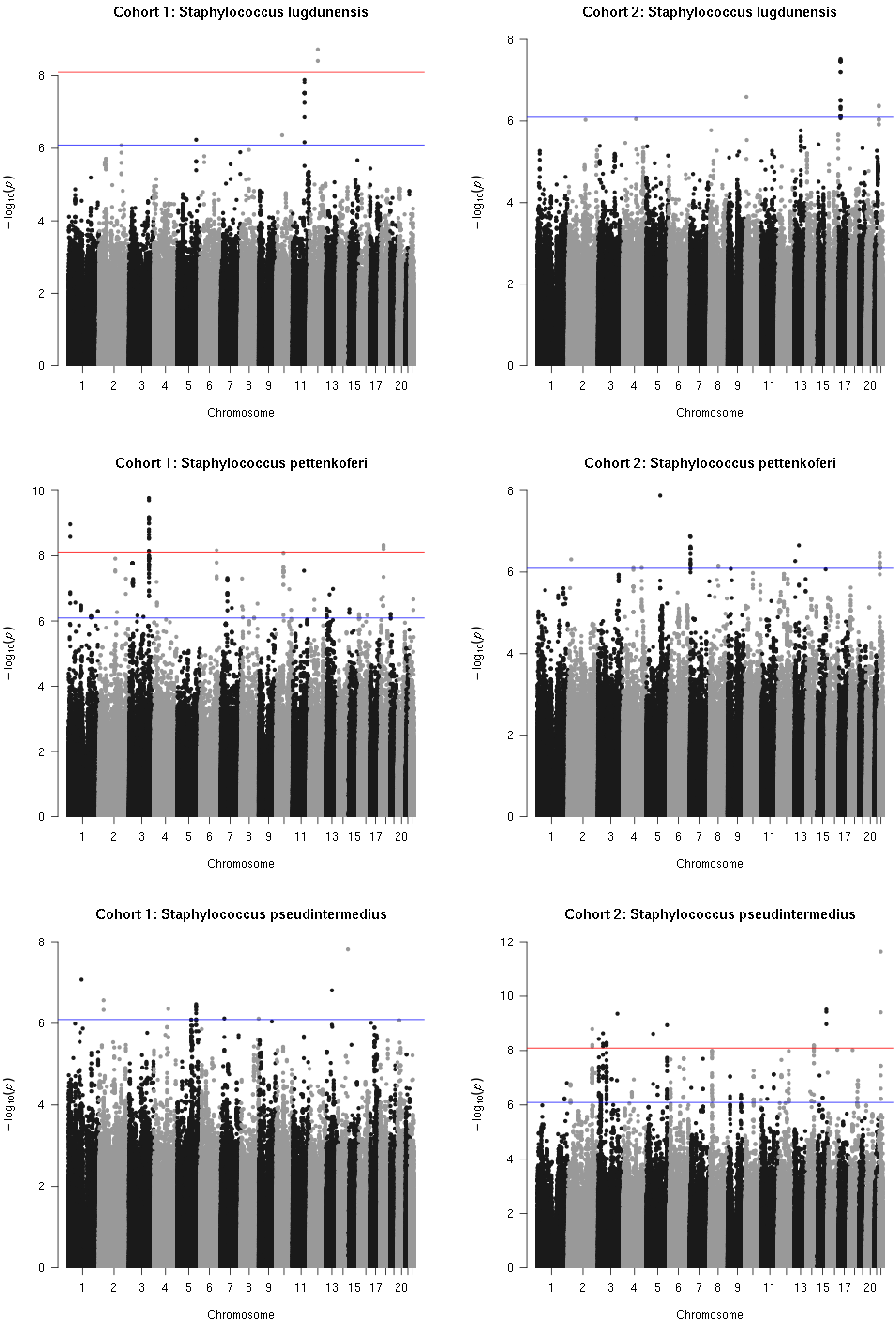
Cohort 1: Staphylococcus lugdunensis
Cohort 2: Staphylococcus lugdunensis
Cohort 1: Staphylococcus pettenkoferi
Cohort 2: Staphylococcus pettenkoferi
Cohort 1: Staphylococcus pseudintermedius
Cohort 2: Staphylococcus pseudintermedius
$-\log_{10}(p)$
Chromosome
1
2
3
4
5
6
7
8
9
11
13
15
17
20

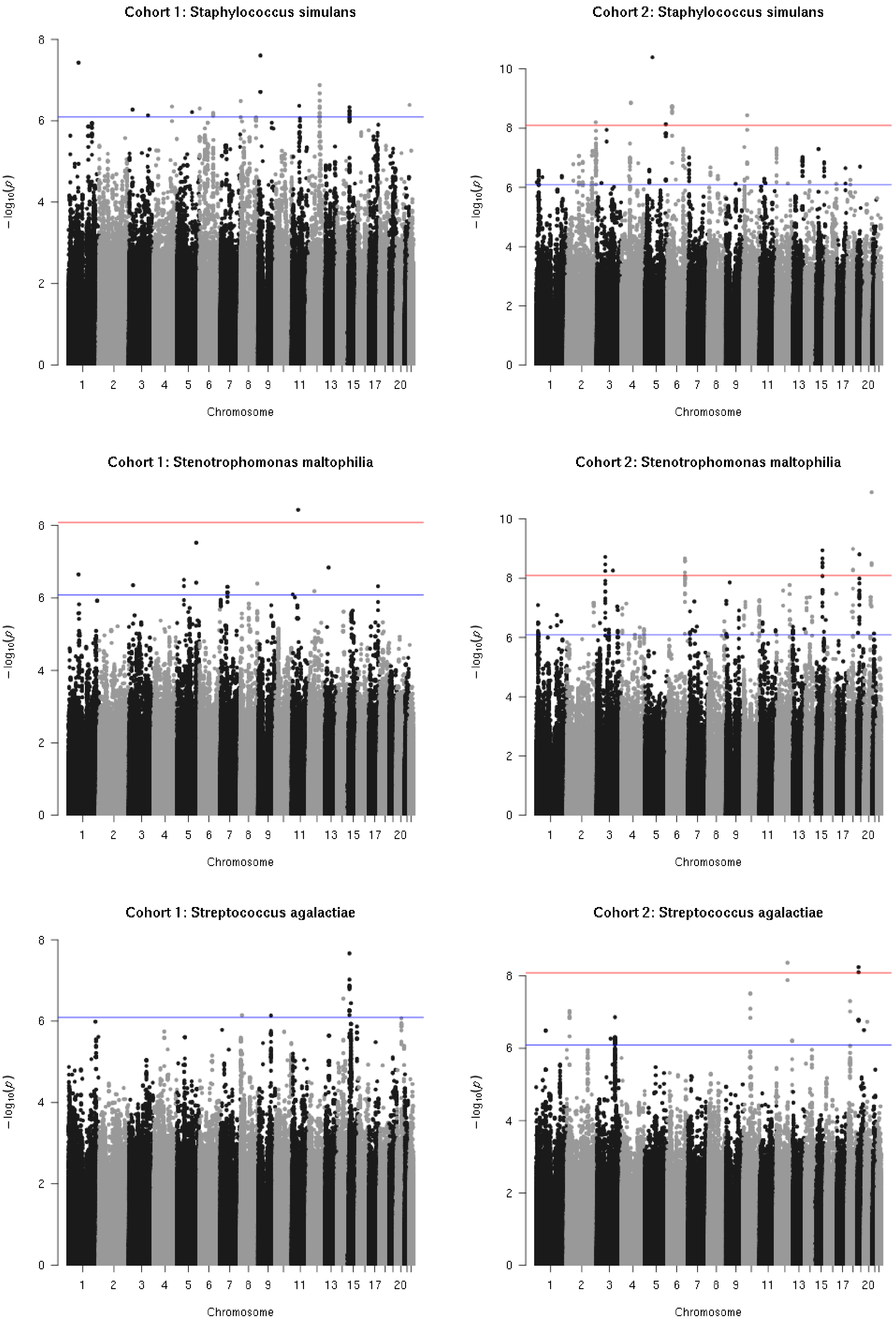
Cohort 1: Staphylococcus simulans
Cohort 2: Staphylococcus simulans
Cohort 1: Stenotrophomonas maltophilia
Cohort 2: Stenotrophomonas maltophilia
Cohort 1: Streptococcus agalactiae
Cohort 2: Streptococcus agalactiae
$-\log_{10}(p)$
Chromosome
1 2 3 4 5 6 7 8 9 11 13 15 17 20
0 2 4 6 8 10

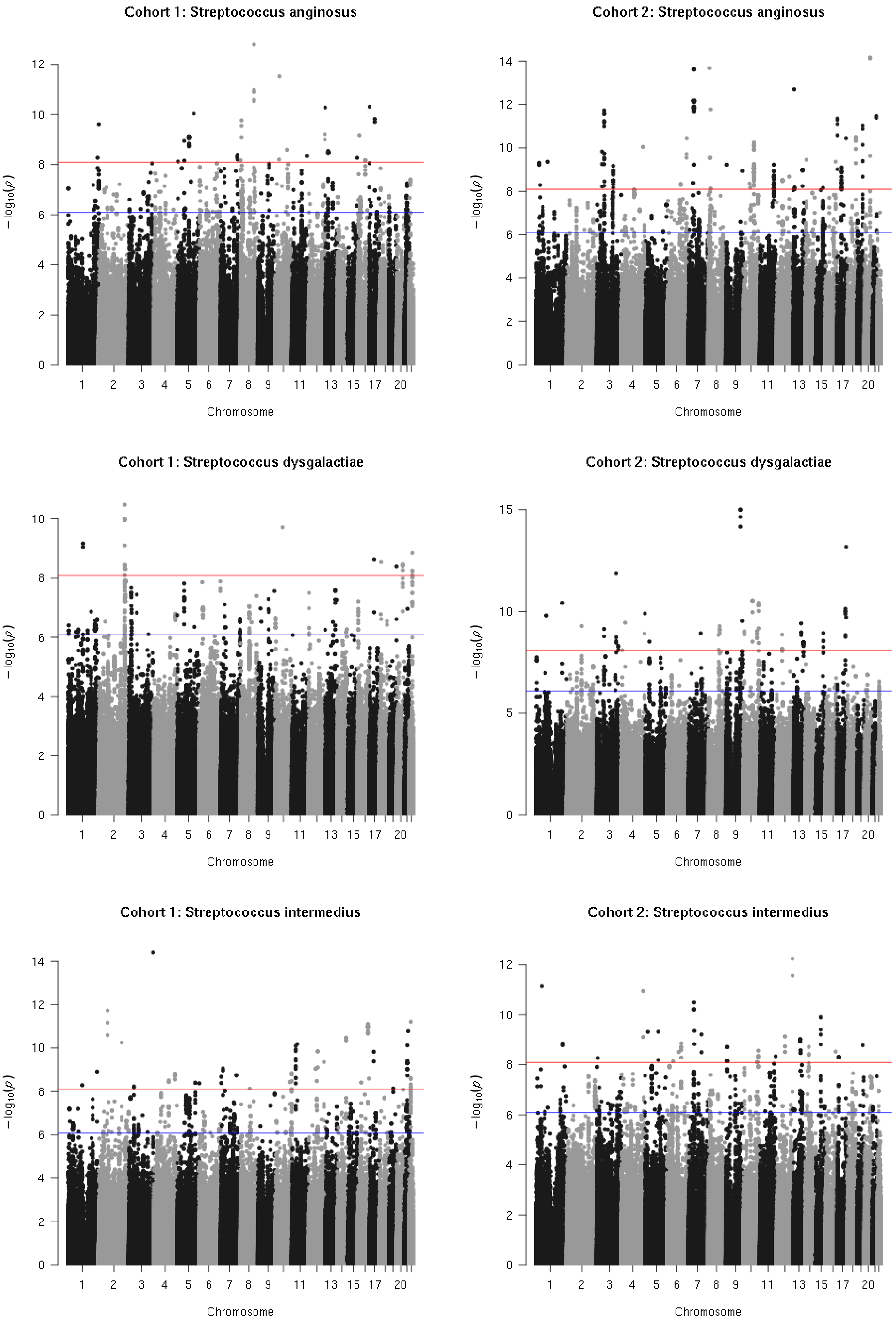

Cohort 1: Streptococcus anginosus
Cohort 2: Streptococcus anginosus
Cohort 1: Streptococcus dysgalactiae
Cohort 2: Streptococcus dysgalactiae
Cohort 1: Streptococcus intermedius
Cohort 2: Streptococcus intermedius
$-\log_{10}(p)$
Chromosome
1 2 3 4 5 6 7 8 9 11 13 15 17 20

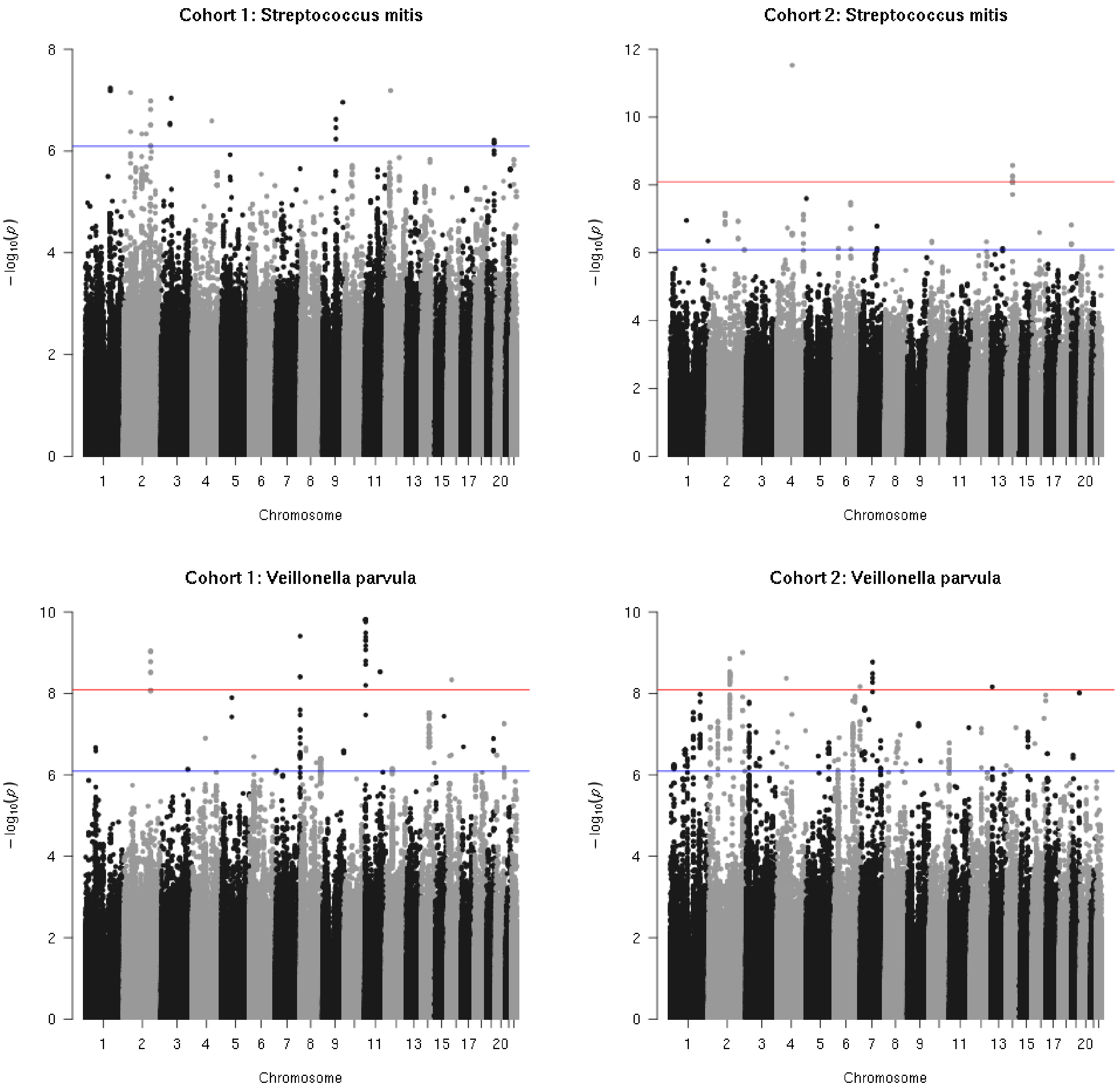
Cohort 1: Streptococcus mitis
Cohort 2: Streptococcus mitis
Cohort 1: Veillonella parvula
Cohort 2: Veillonella parvula
$-\log_{10}(p)$
Chromosome
1
2
3
4
5
6
7
8
9
11
13
15
17
20

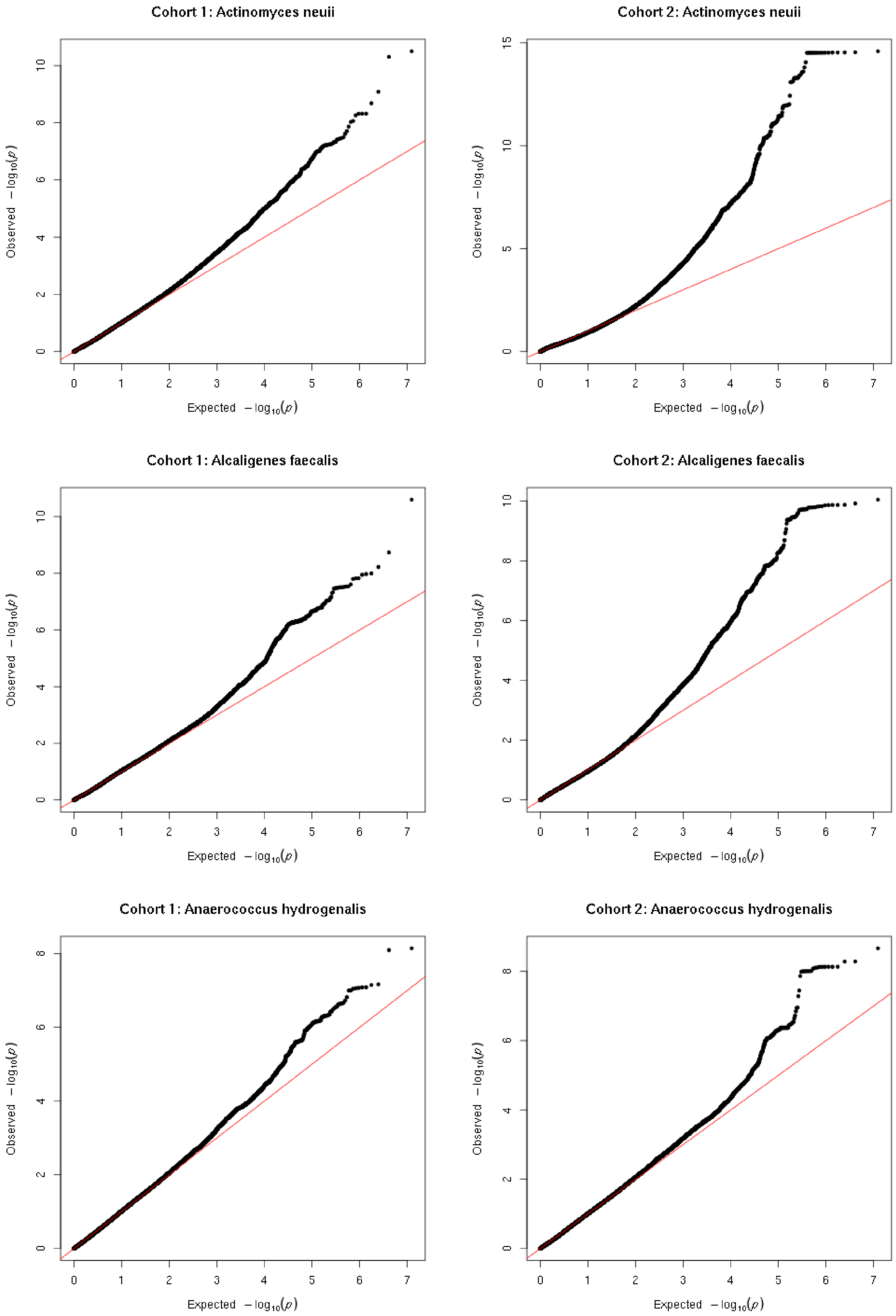
Cohort 1: Actinomyces neuii
Cohort 2: Actinomyces neuii
Cohort 1: Alcaligenes faecalis
Cohort 2: Alcaligenes faecalis
Cohort 1: Anaerococcus hydrogenalis
Cohort 2: Anaerococcus hydrogenalis
Observed $-\log_{10}(p)$
Expected $-\log_{10}(p)$

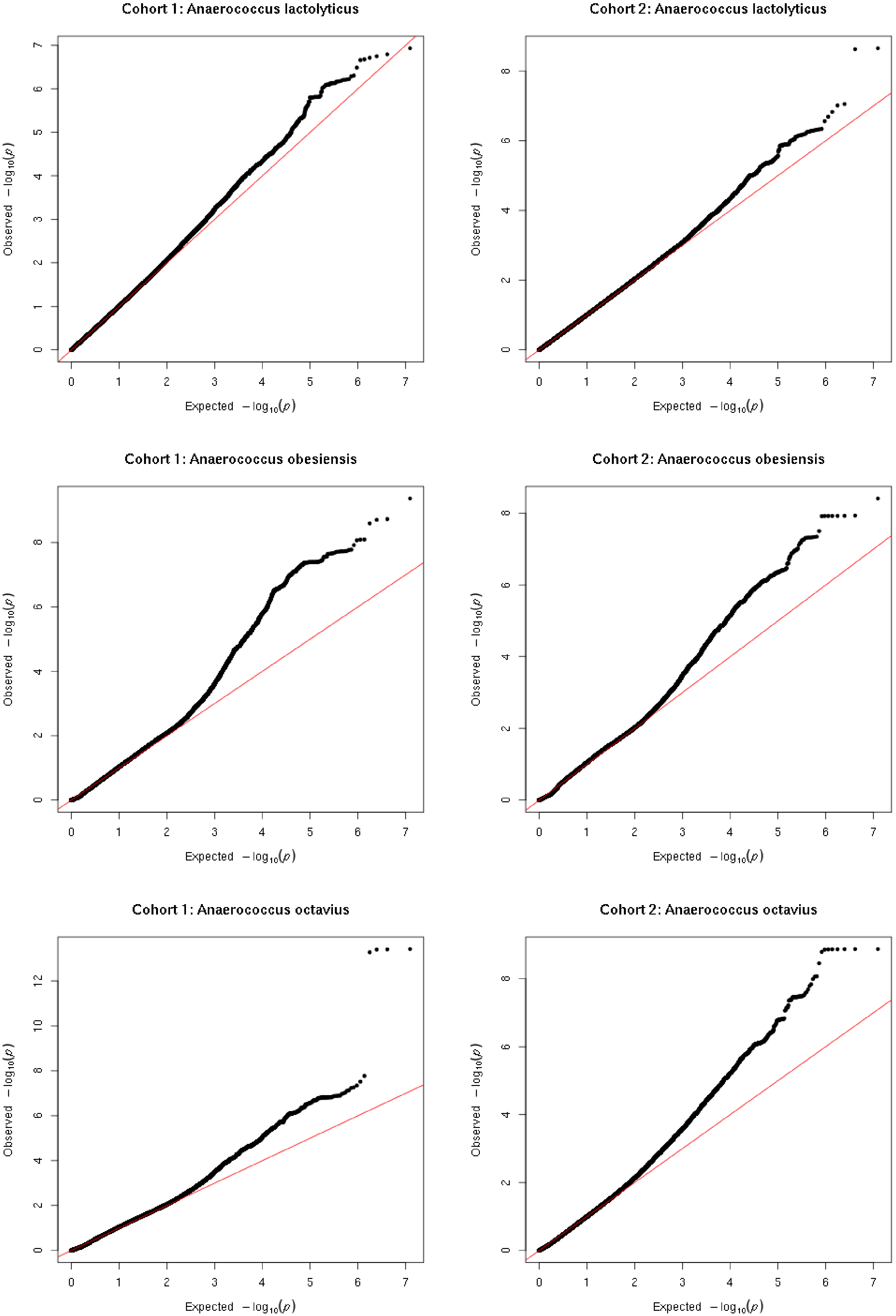
Cohort 1: Anaerococcus lactolyticus
Cohort 2: Anaerococcus lactolyticus
Cohort 1: Anaerococcus obesiensis
Cohort 2: Anaerococcus obesiensis
Cohort 1: Anaerococcus octavius
Cohort 2: Anaerococcus octavius
Observed $-\log_{10}(p)$
Expected $-\log_{10}(p)$

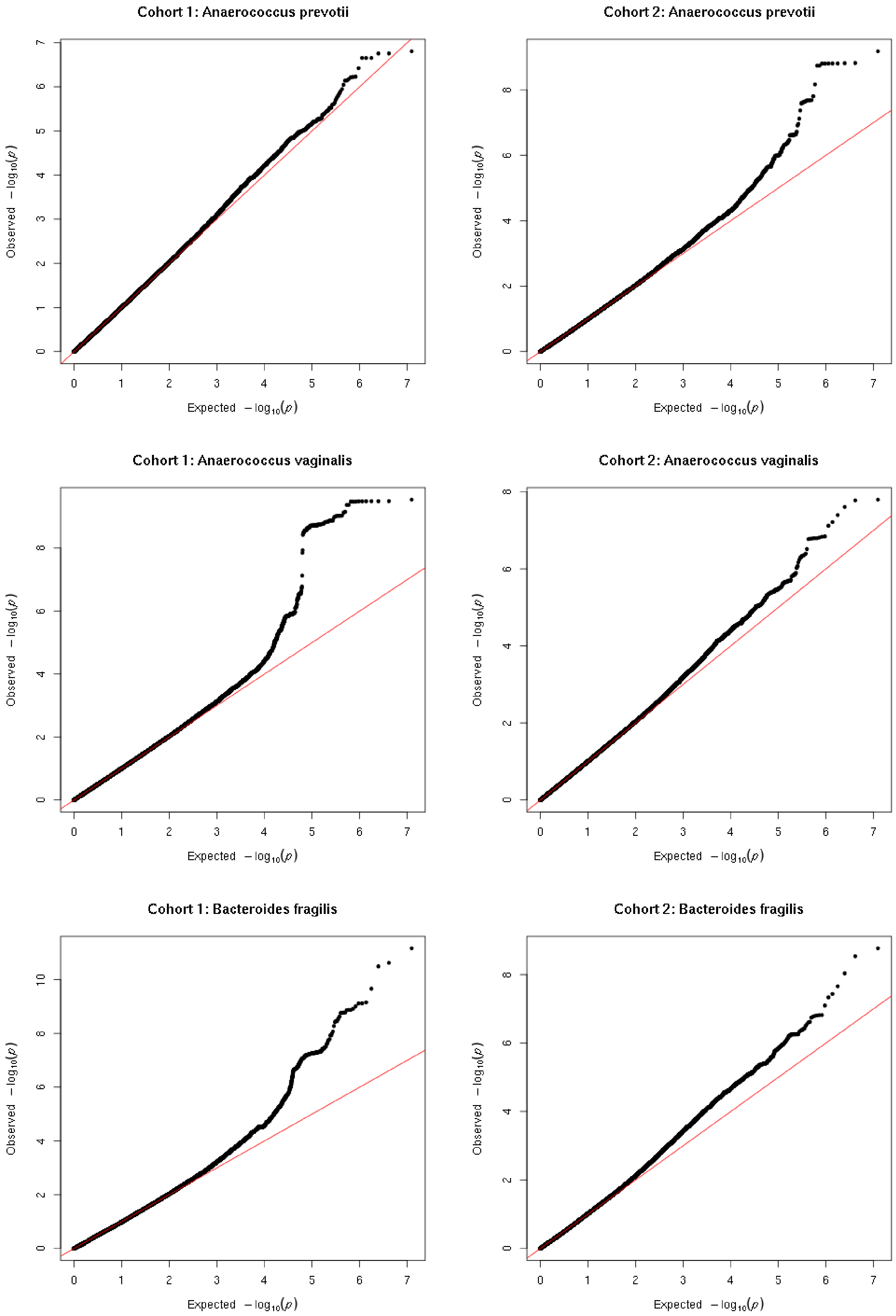
Cohort 1: Anaerococcus prevotii
Cohort 2: Anaerococcus prevotii
Cohort 1: Anaerococcus vaginalis
Cohort 2: Anaerococcus vaginalis
Cohort 1: Bacteroides fragilis
Cohort 2: Bacteroides fragilis
Observed $-\log_{10}(p)$
Expected $-\log_{10}(p)$

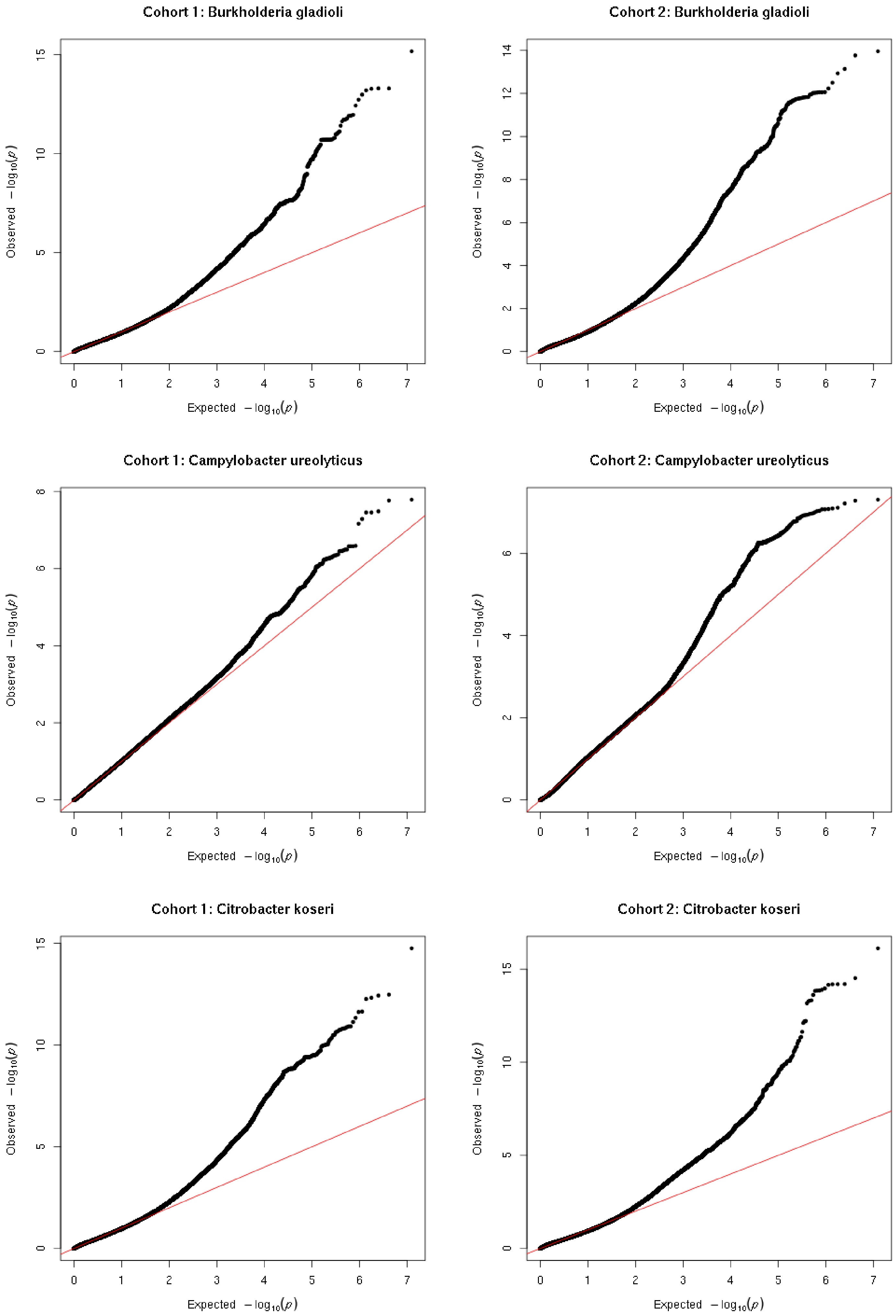
Cohort 1: Burkholderia gladioli
Cohort 2: Burkholderia gladioli
Cohort 1: Campylobacter ureolyticus
Cohort 2: Campylobacter ureolyticus
Cohort 1: Citrobacter koseri
Cohort 2: Citrobacter koseri
Observed $-\log_{10}(p)$
Expected $-\log_{10}(p)$

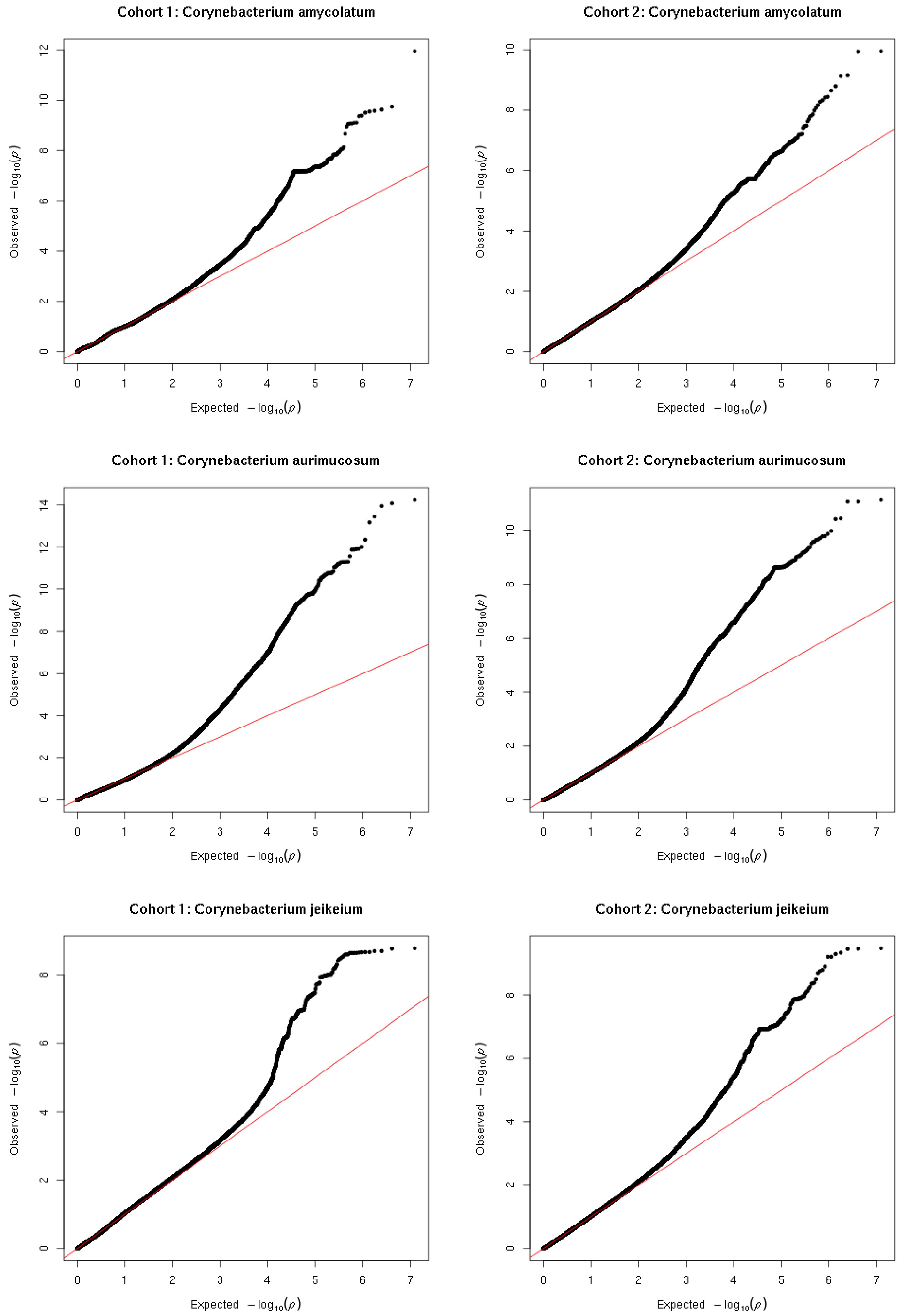
Cohort 1: Corynebacterium amycolatum
Cohort 2: Corynebacterium amycolatum
Cohort 1: Corynebacterium aurimucosum
Cohort 2: Corynebacterium aurimucosum
Cohort 1: Corynebacterium jeikeium
Cohort 2: Corynebacterium jeikeium
Observed $-\log_{10}(p)$
Expected $-\log_{10}(p)$

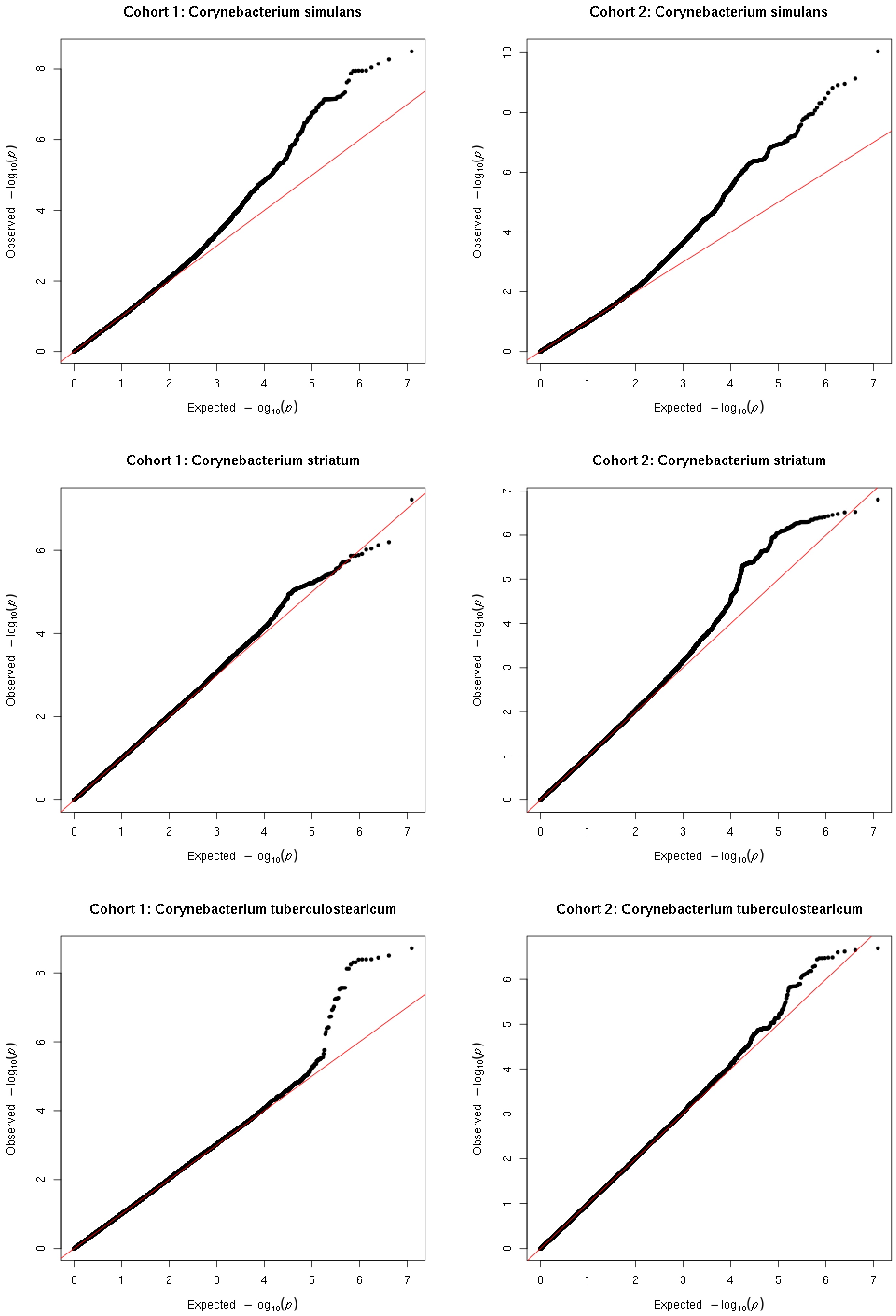
Cohort 1: Corynebacterium simulans
Cohort 2: Corynebacterium simulans
Cohort 1: Corynebacterium striatum
Cohort 2: Corynebacterium striatum
Cohort 1: Corynebacterium tuberculostearicum
Cohort 2: Corynebacterium tuberculostearicum
Observed $-\log_{10}(p)$
Expected $-\log_{10}(p)$

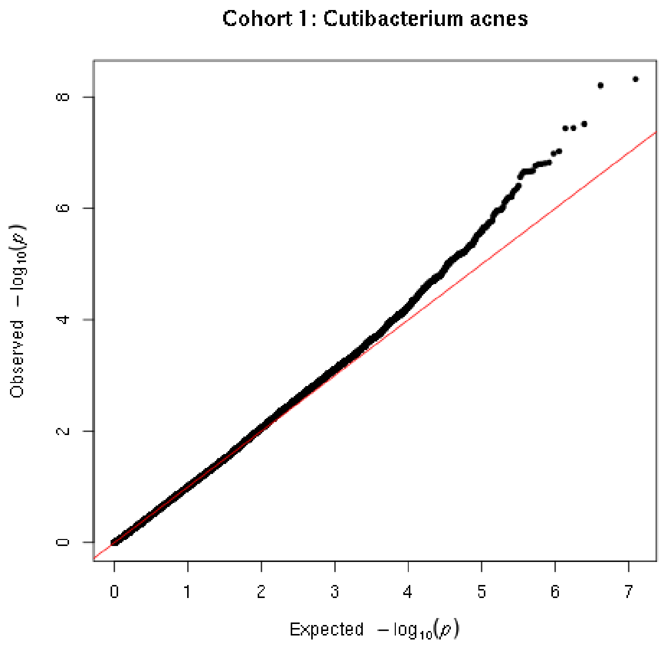
Cohort 1: Cutibacterium acnes
Observed $-\log_{10}(p)$
Expected $-\log_{10}(p)$

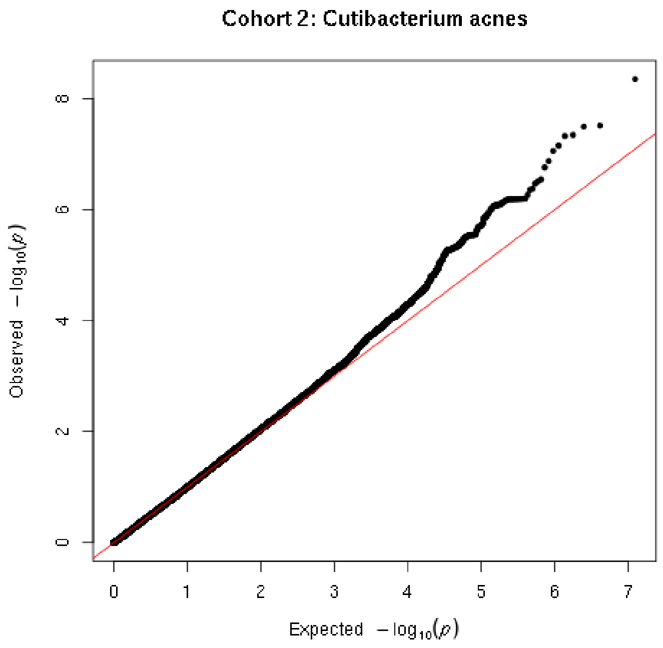
Cohort 2: Cutibacterium acnes
Observed $-\log_{10}(p)$
Expected $-\log_{10}(p)$

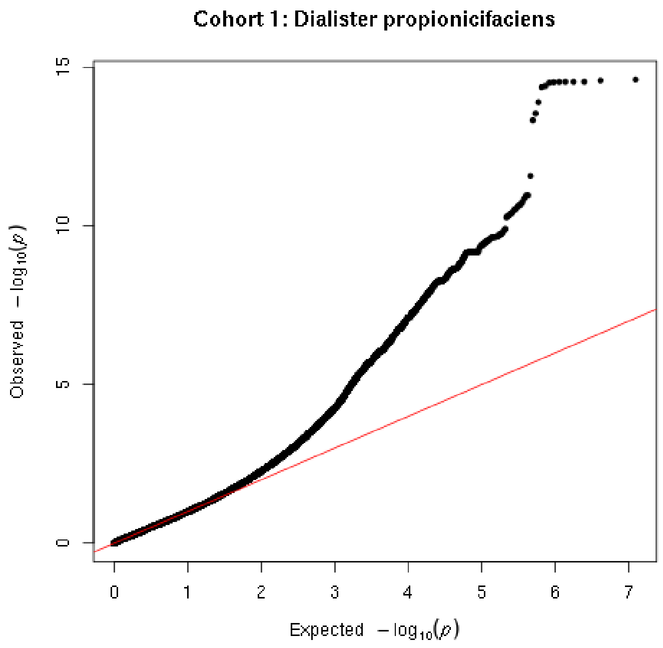
Cohort 1: Dialister propionicifaciens
Observed $-\log_{10}(p)$
Expected $-\log_{10}(p)$

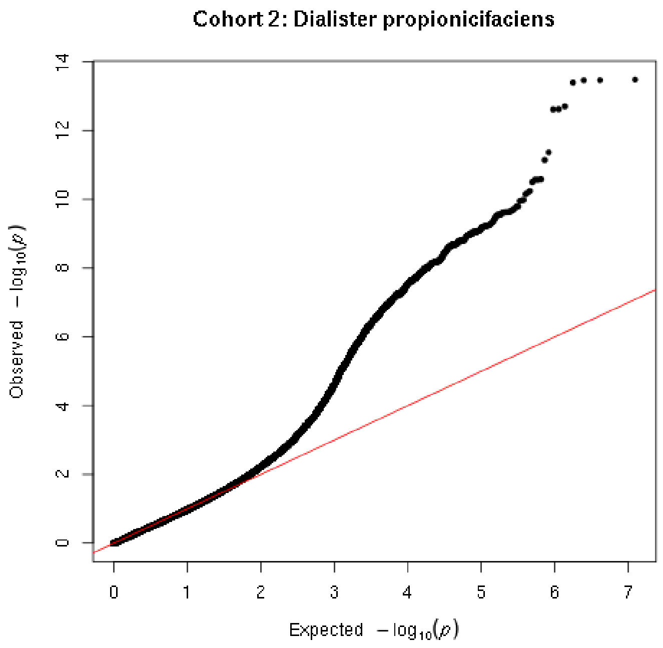
Cohort 2: Dialister propionicifaciens
Observed $-\log_{10}(p)$
Expected $-\log_{10}(p)$

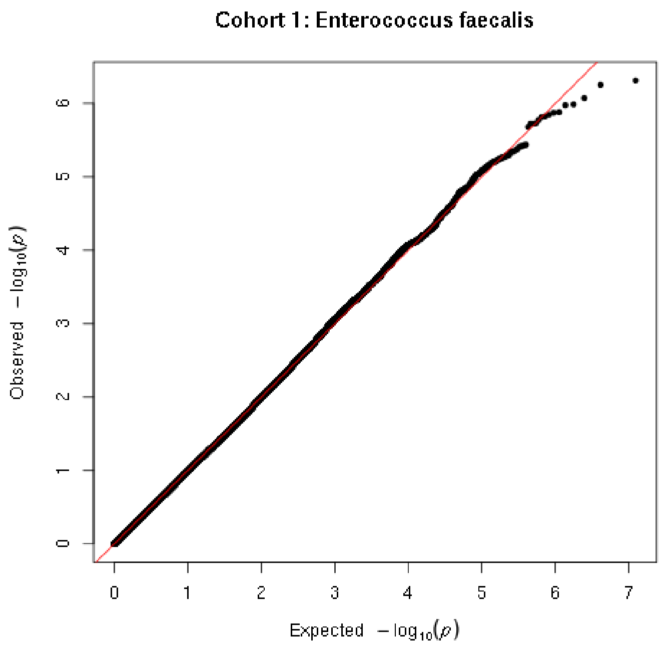
Cohort 1: Enterococcus faecalis
Observed $-\log_{10}(p)$
Expected $-\log_{10}(p)$

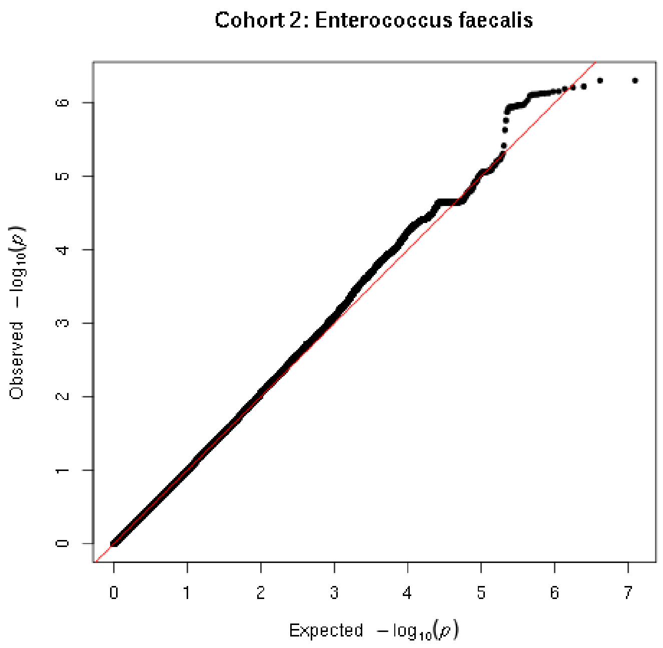
Cohort 2: Enterococcus faecalis
Observed $-\log_{10}(p)$
Expected $-\log_{10}(p)$

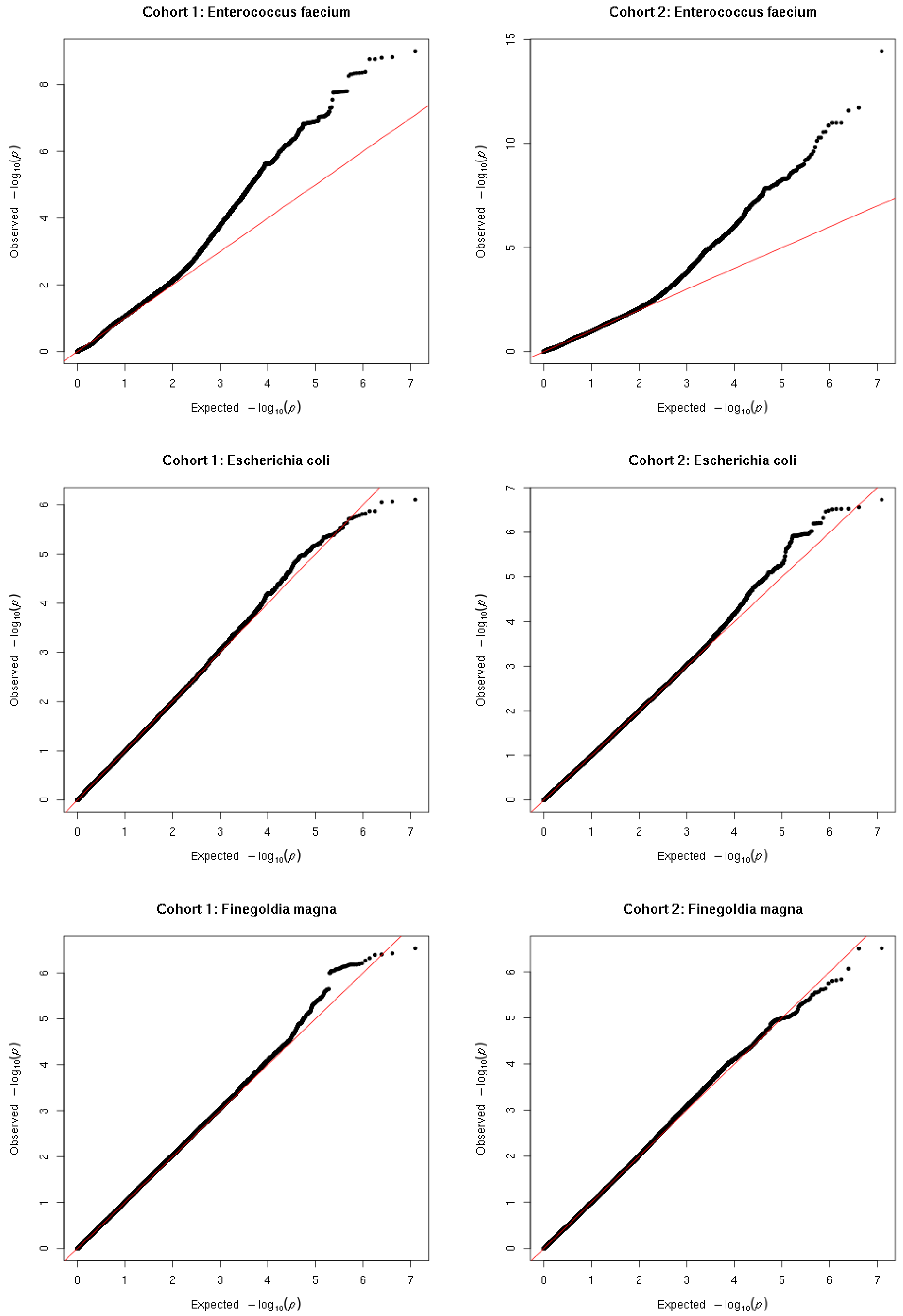
Cohort 1: Enterococcus faecium
Cohort 2: Enterococcus faecium
Cohort 1: Escherichia coli
Cohort 2: Escherichia coli
Cohort 1: Finegoldia magna
Cohort 2: Finegoldia magna
Observed $-\log_{10}(p)$
Expected $-\log_{10}(p)$

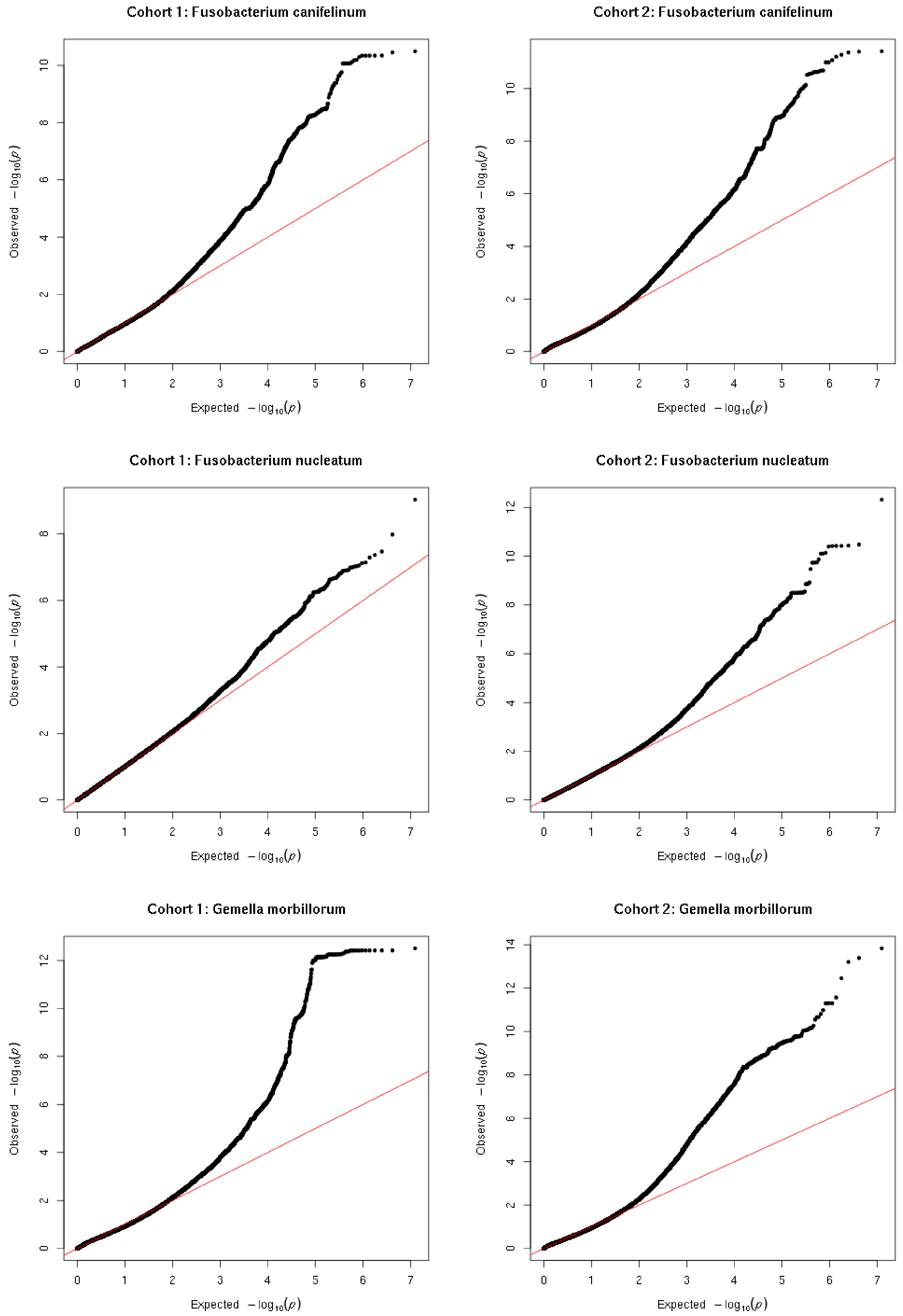
Cohort 1: Fusobacterium canifelinum
Cohort 2: Fusobacterium canifelinum
Cohort 1: Fusobacterium nucleatum
Cohort 2: Fusobacterium nucleatum
Cohort 1: Gemella morbillorum
Cohort 2: Gemella morbillorum
Observed $-\log_{10}(p)$
Expected $-\log_{10}(p)$

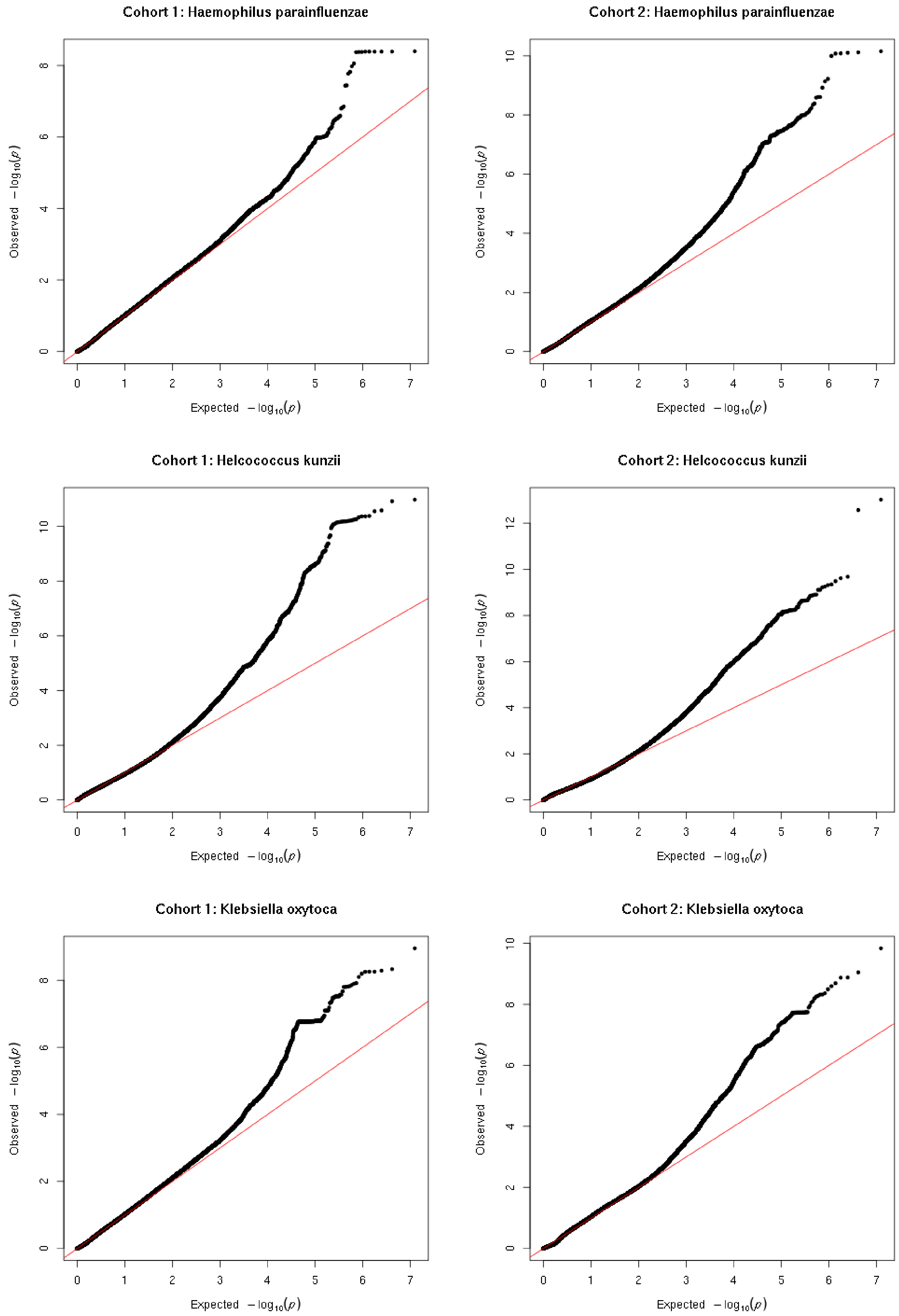
Cohort 1: Haemophilus parainfluenzae
Cohort 2: Haemophilus parainfluenzae
Cohort 1: Helcococcus kunzii
Cohort 2: Helcococcus kunzii
Cohort 1: Klebsiella oxytoca
Cohort 2: Klebsiella oxytoca
Observed $-\log_{10}(p)$
Expected $-\log_{10}(p)$

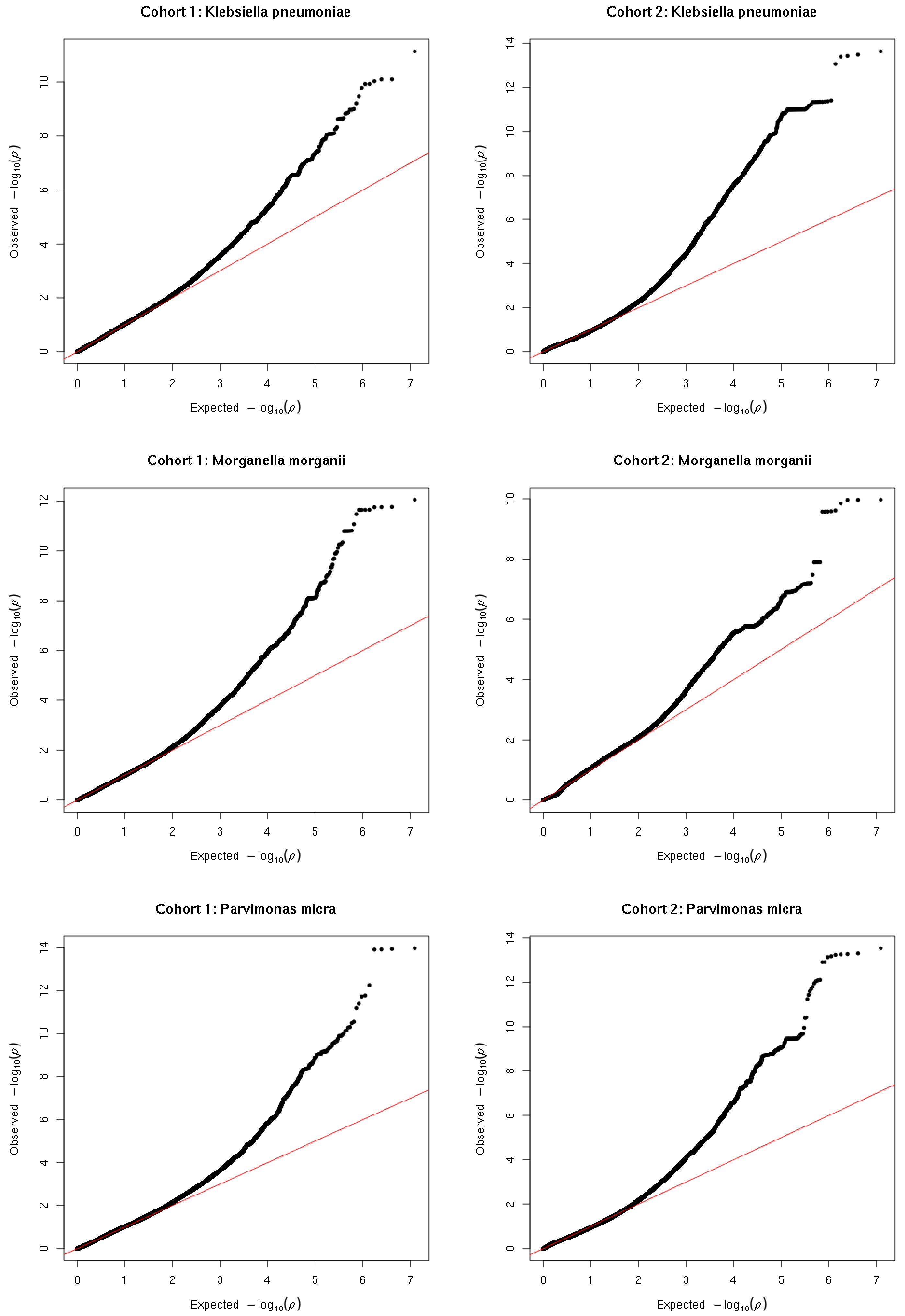
Cohort 1: Klebsiella pneumoniae
Cohort 2: Klebsiella pneumoniae
Cohort 1: Morganella morganii
Cohort 2: Morganella morganii
Cohort 1: Parvimonas micra
Cohort 2: Parvimonas micra
Observed $-\log_{10}(p)$
Expected $-\log_{10}(p)$

Cohort 1: Pasteurella canis

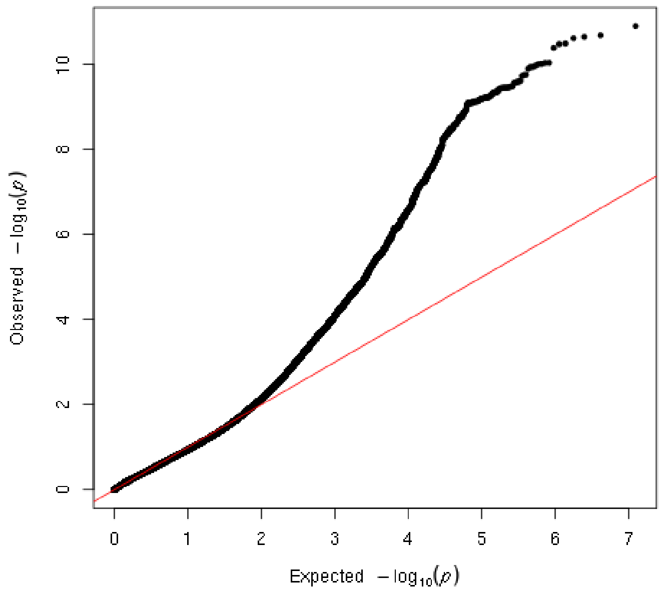


Cohort 2: Pasteurella canis

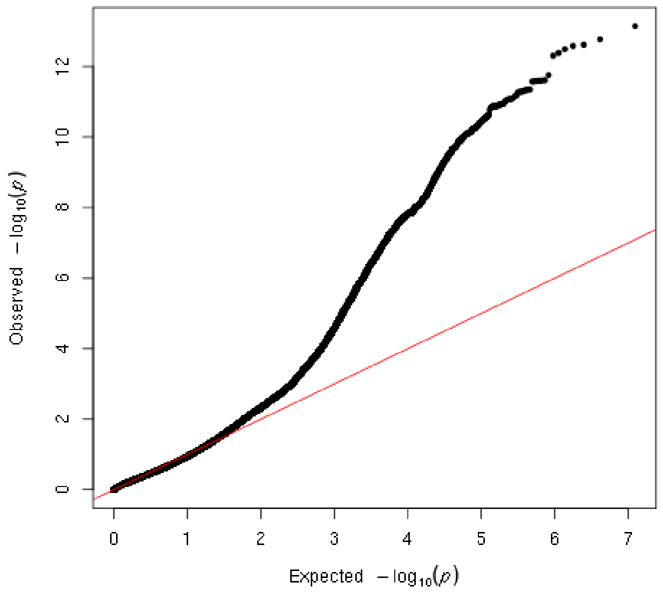


Cohort 1: Pelomonas saccharophila

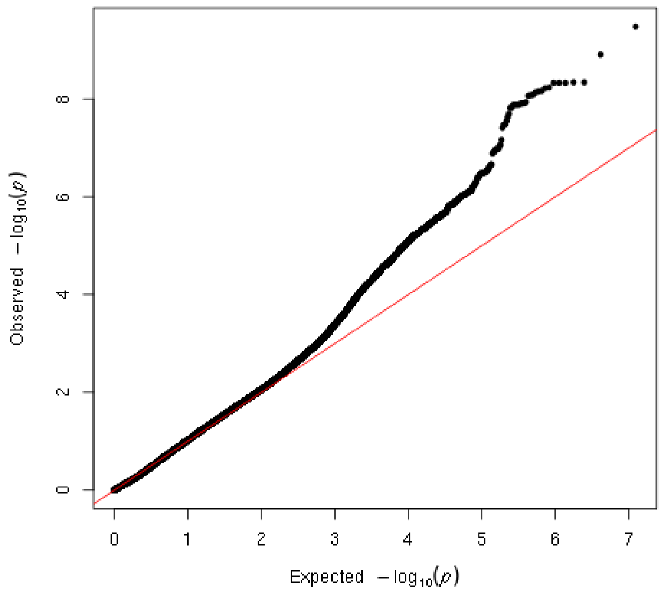


Cohort 2: Pelomonas saccharophila

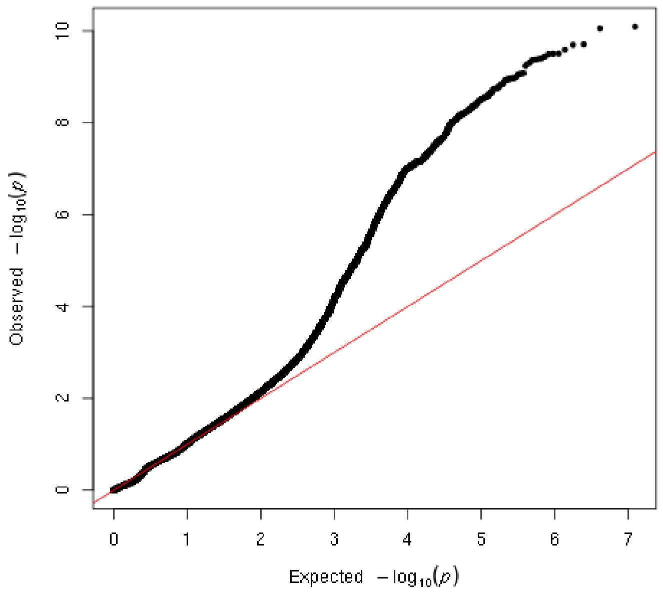


Cohort 1: Peptoniphilus asaccharolyticus

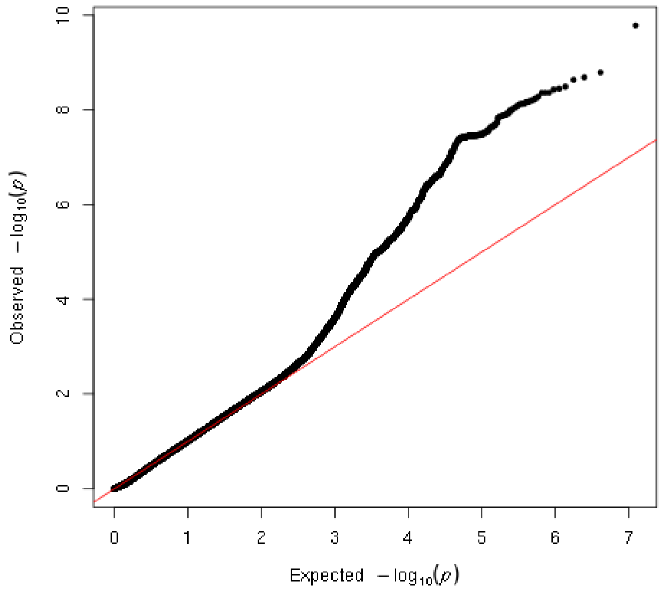


Cohort 2: Peptoniphilus asaccharolyticus

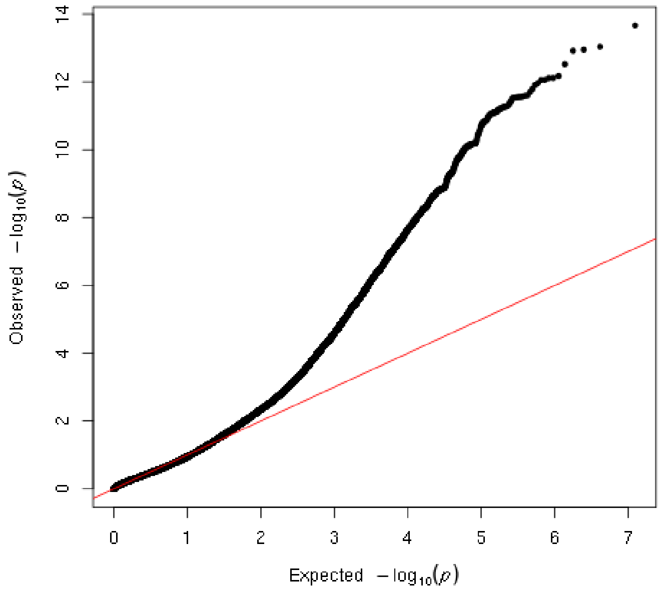

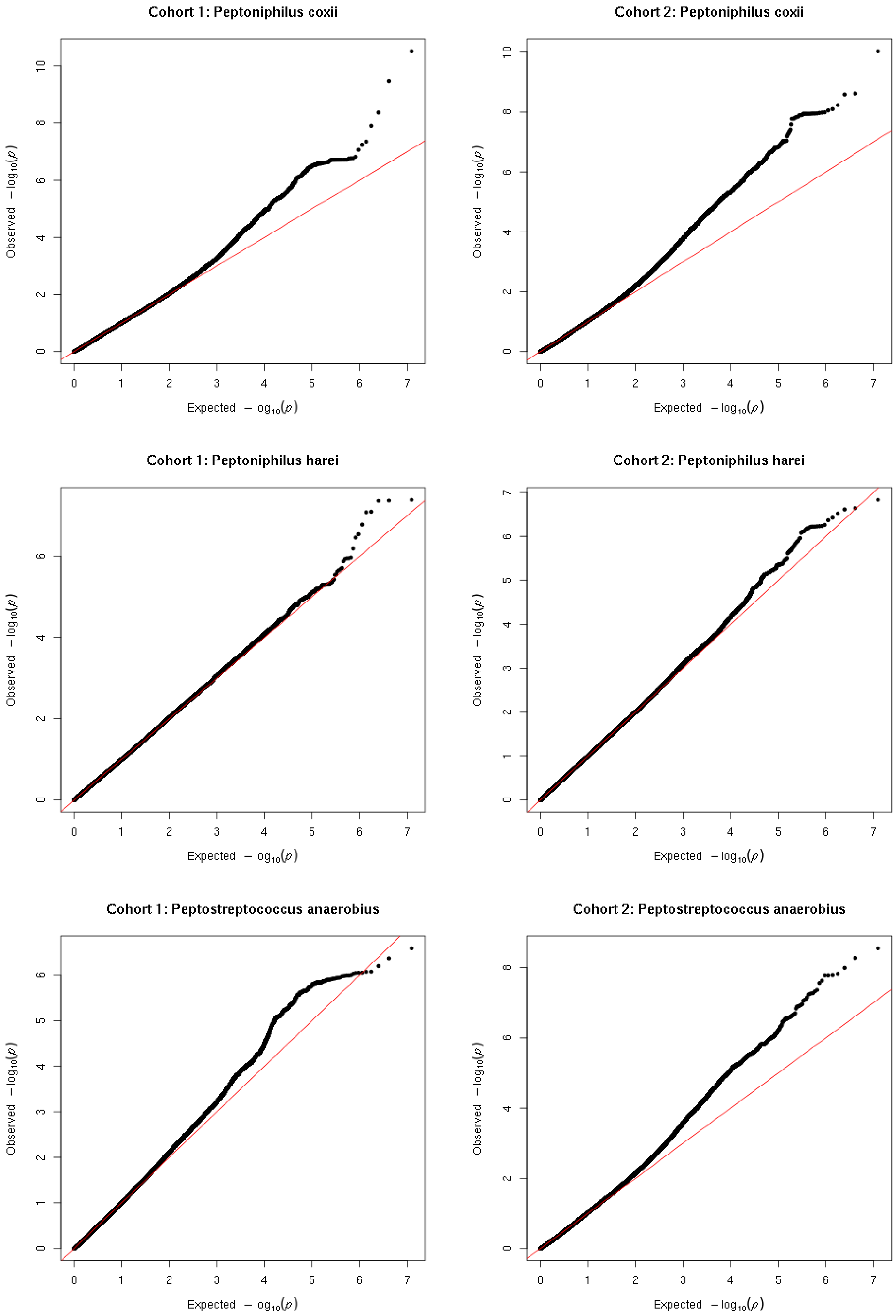
Cohort 1: Peptoniphilus coxii
Cohort 2: Peptoniphilus coxii
Cohort 1: Peptoniphilus harei
Cohort 2: Peptoniphilus harei
Cohort 1: Peptostreptococcus anaerobius
Cohort 2: Peptostreptococcus anaerobius
Observed $-\log_{10}(p)$
Expected $-\log_{10}(p)$

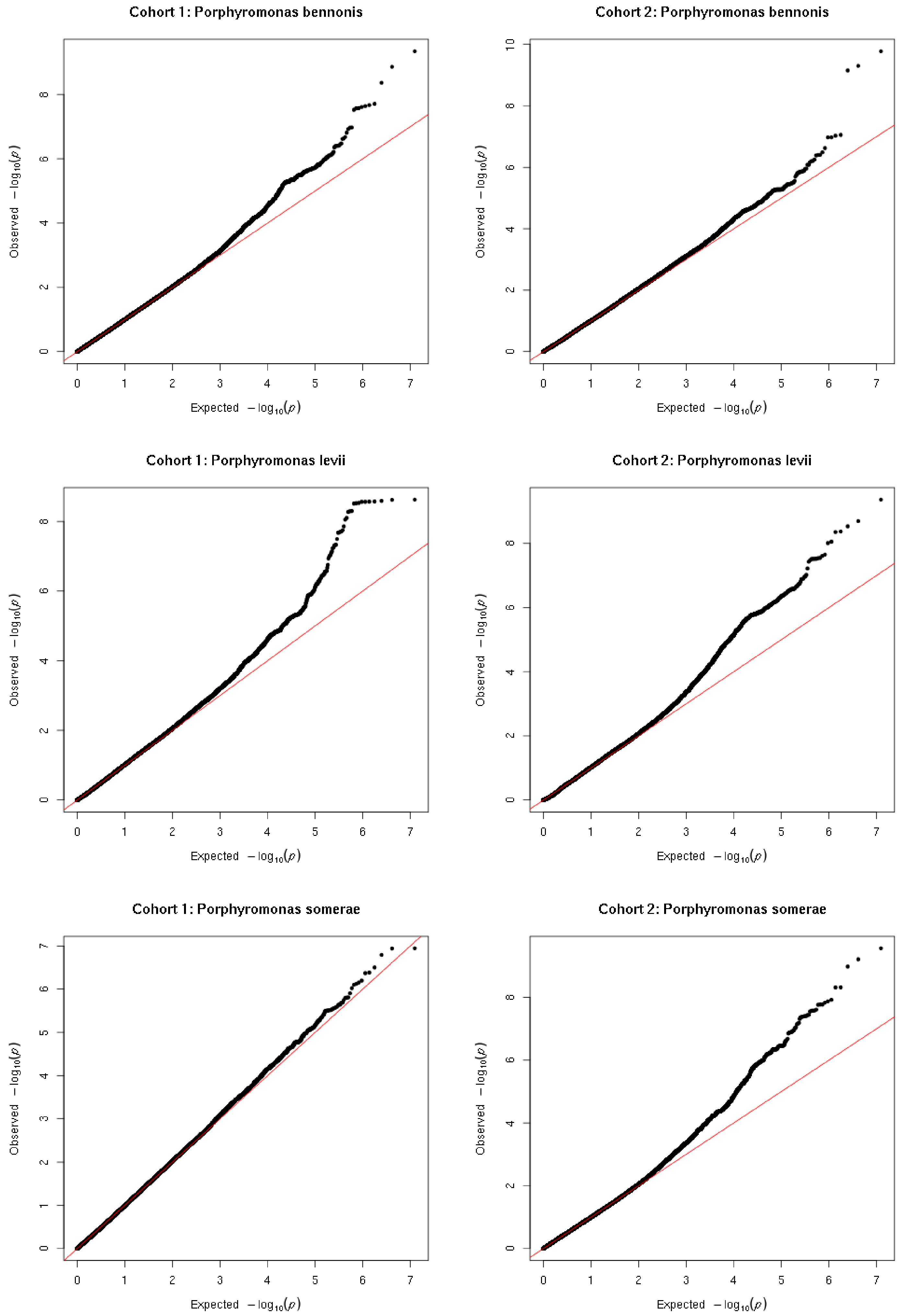
Cohort 1: Porphyromonas bennonis
Cohort 2: Porphyromonas bennonis
Cohort 1: Porphyromonas levii
Cohort 2: Porphyromonas levii
Cohort 1: Porphyromonas somerae
Cohort 2: Porphyromonas somerae
Observed $-\log_{10}(p)$
Expected $-\log_{10}(p)$

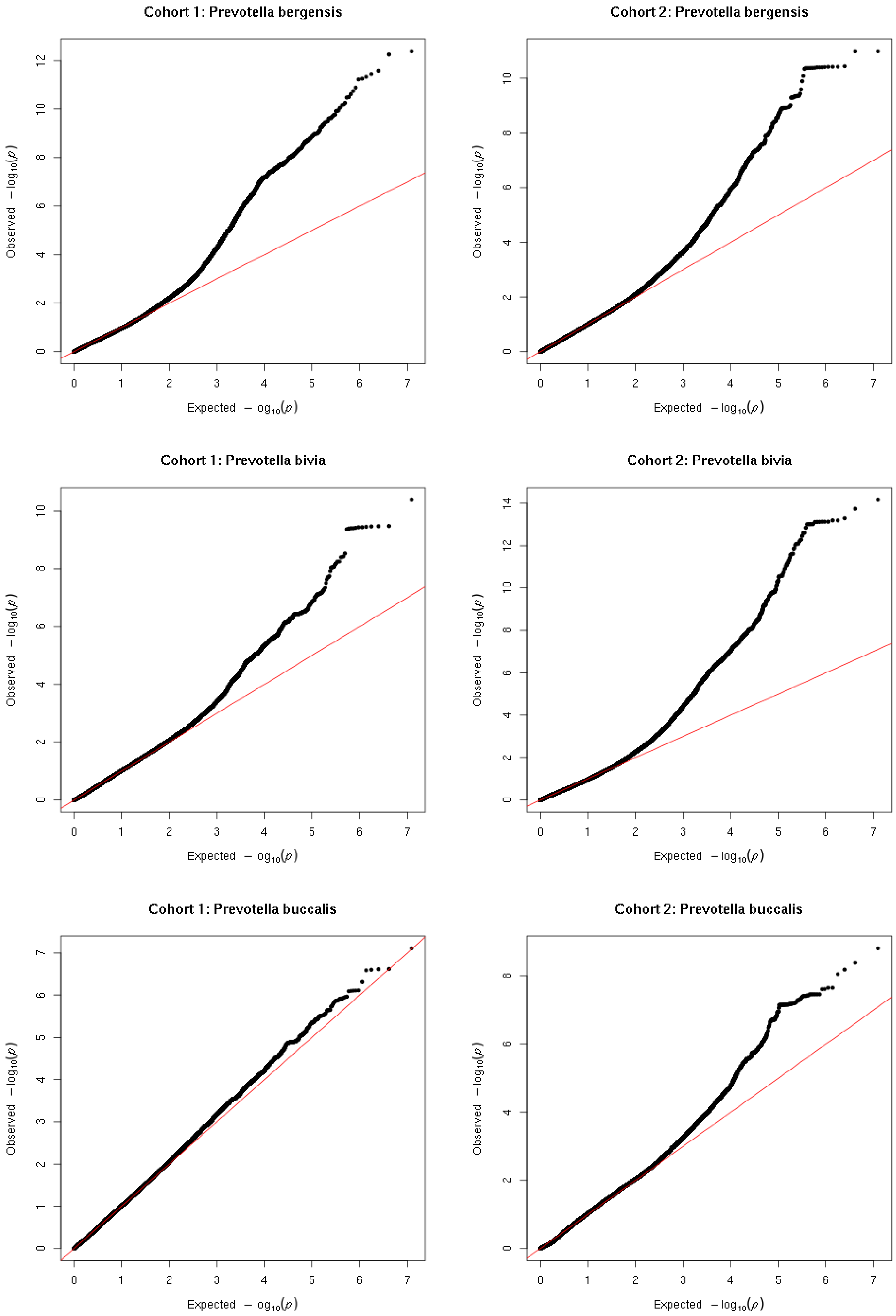
Cohort 1: Prevotella bergensis
Cohort 2: Prevotella bergensis
Cohort 1: Prevotella bivia
Cohort 2: Prevotella bivia
Cohort 1: Prevotella buccalis
Cohort 2: Prevotella buccalis
Observed $-\log_{10}(p)$
Expected $-\log_{10}(p)$

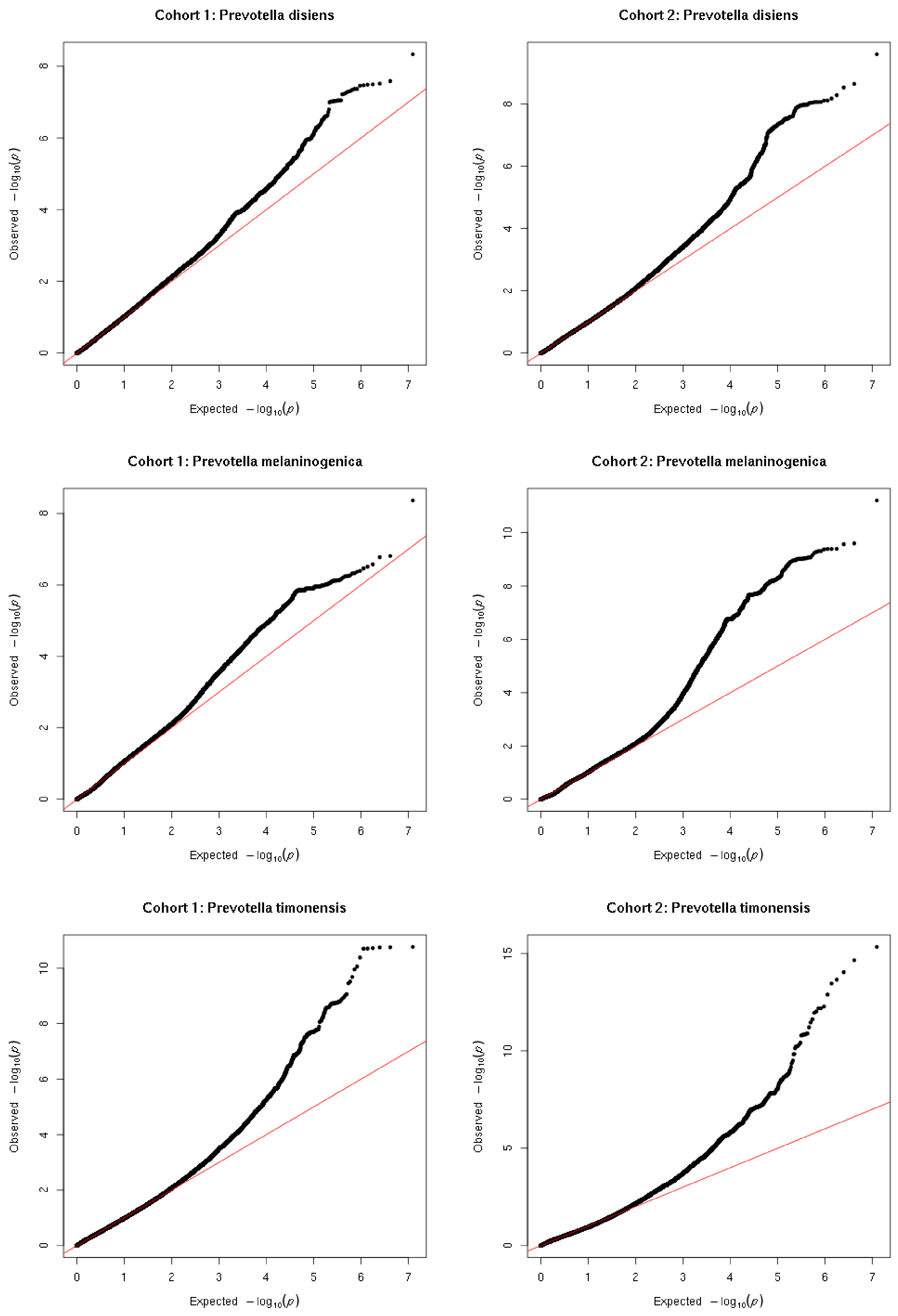
Cohort 1: Prevotella disiens
Cohort 2: Prevotella disiens
Cohort 1: Prevotella melaninogenica
Cohort 2: Prevotella melaninogenica
Cohort 1: Prevotella timonensis
Cohort 2: Prevotella timonensis
Observed $-\log_{10}(p)$
Expected $-\log_{10}(p)$

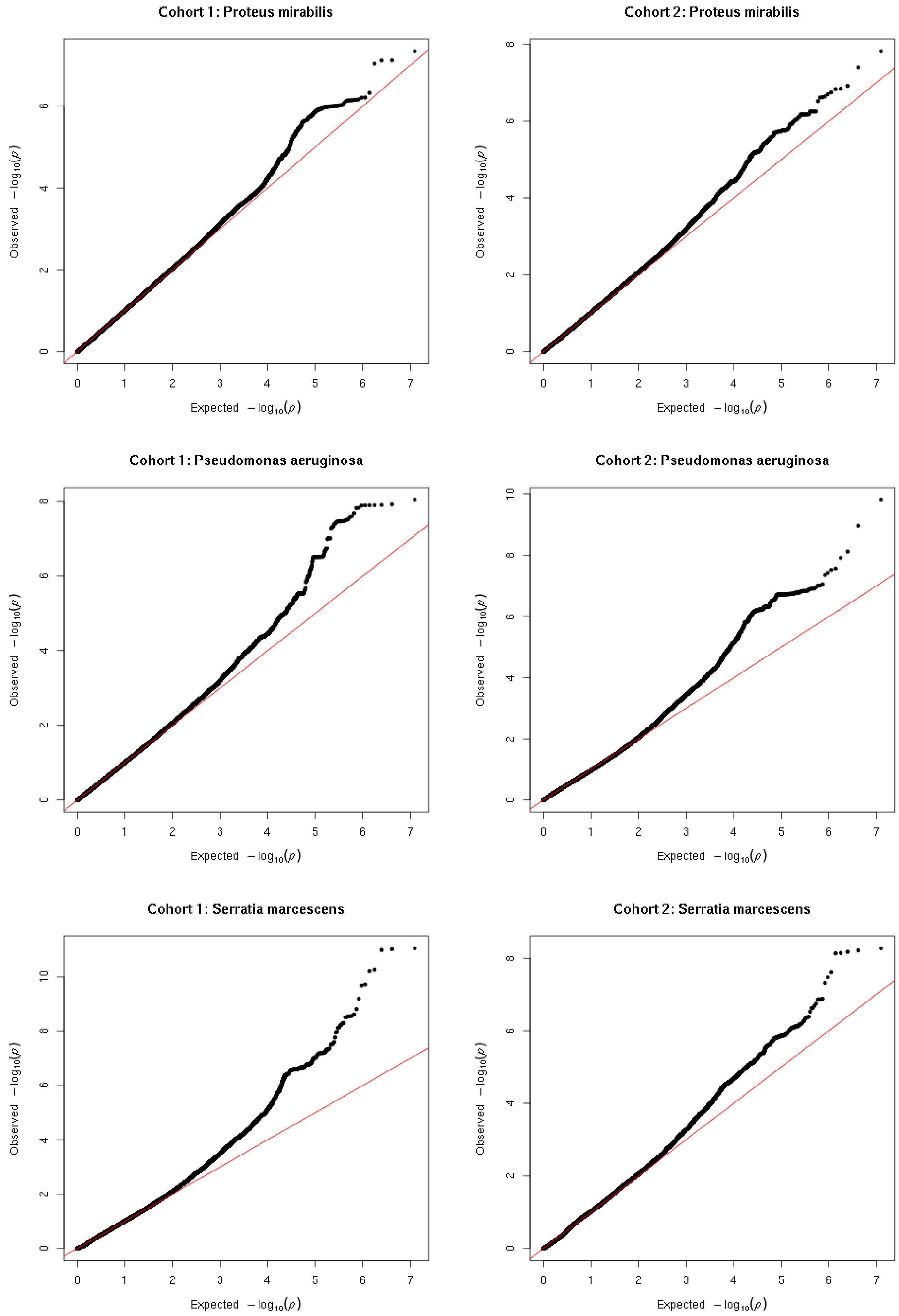
Cohort 1: Proteus mirabilis
Cohort 2: Proteus mirabilis
Cohort 1: Pseudomonas aeruginosa
Cohort 2: Pseudomonas aeruginosa
Cohort 1: Serratia marcescens
Cohort 2: Serratia marcescens
Observed $-\log_{10}(p)$
Expected $-\log_{10}(p)$

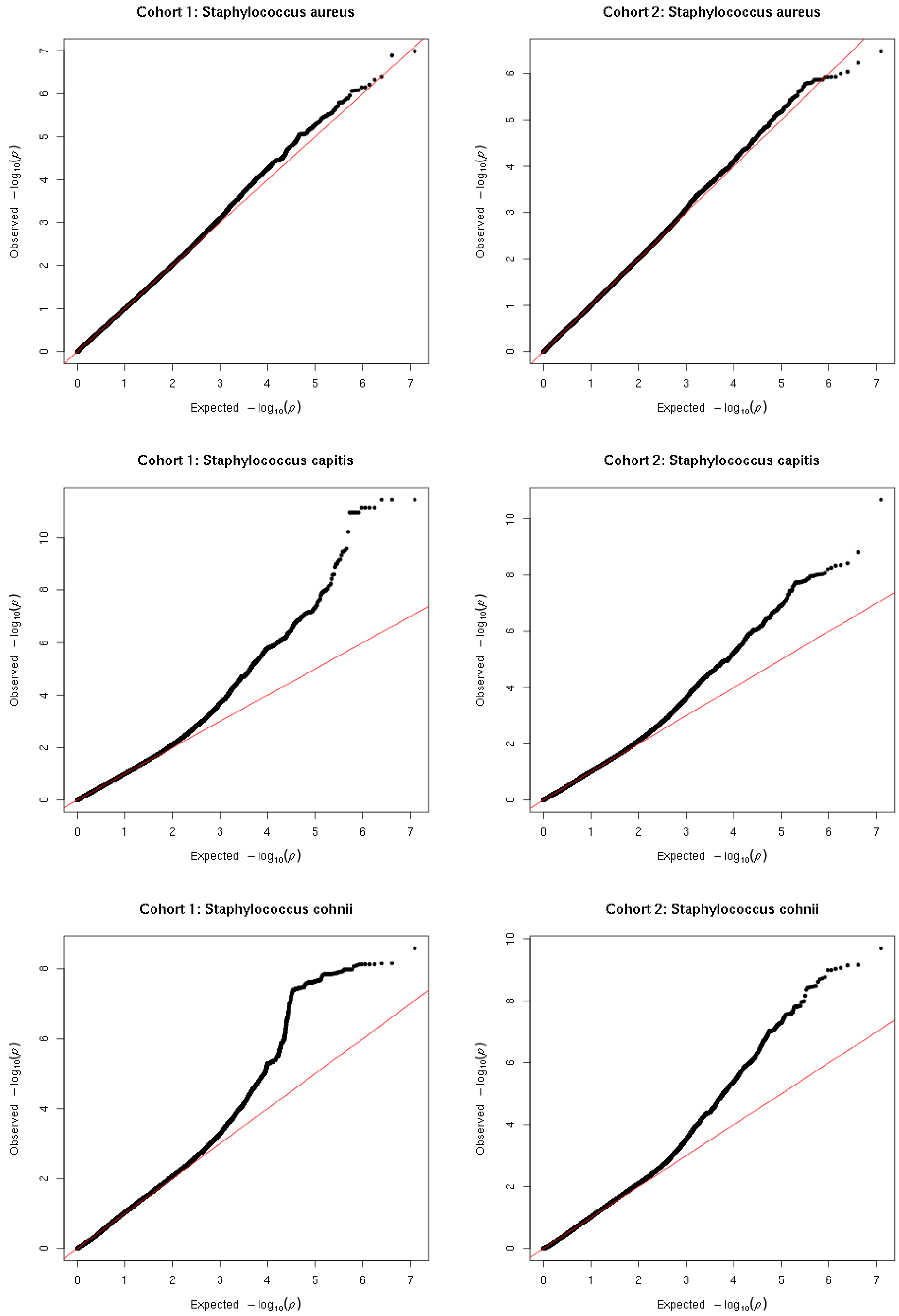
Cohort 1: Staphylococcus aureus
Cohort 2: Staphylococcus aureus
Cohort 1: Staphylococcus capitis
Cohort 2: Staphylococcus capitis
Cohort 1: Staphylococcus cohnii
Cohort 2: Staphylococcus cohnii
Observed $-\log_{10}(p)$
Expected $-\log_{10}(p)$

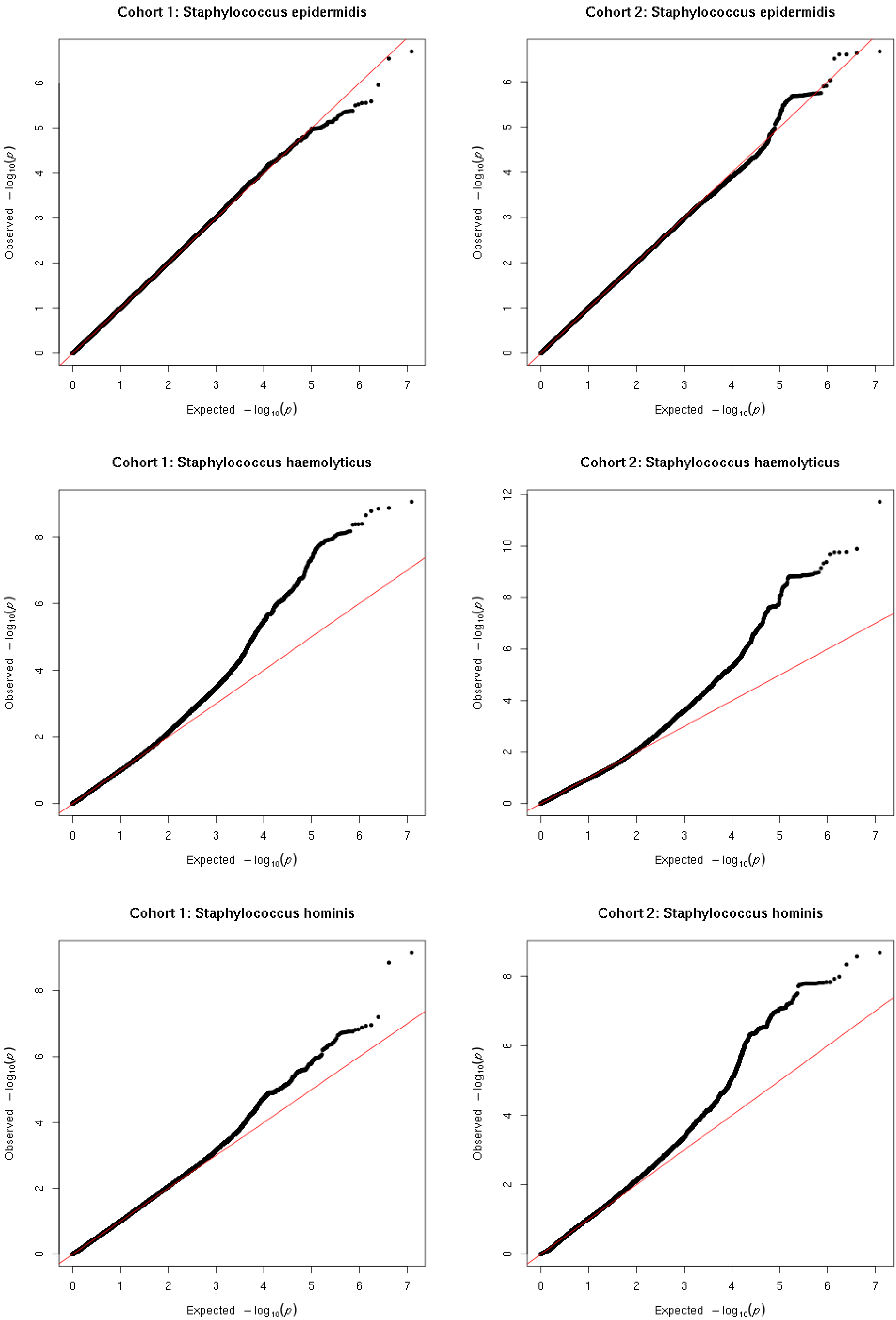

Cohort 1: Staphylococcus epidermidis
Cohort 2: Staphylococcus epidermidis
Cohort 1: Staphylococcus haemolyticus
Cohort 2: Staphylococcus haemolyticus
Cohort 1: Staphylococcus hominis
Cohort 2: Staphylococcus hominis
Observed $-\log_{10}(p)$
Expected $-\log_{10}(p)$

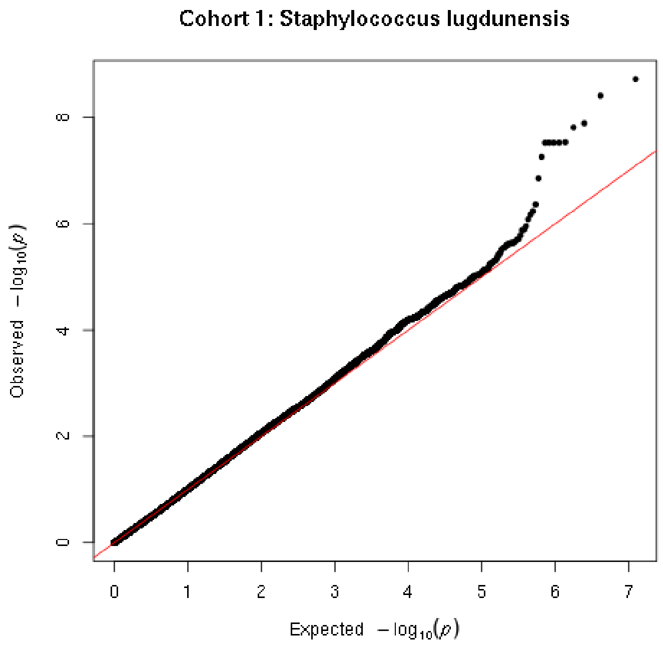
Cohort 1: Staphylococcus lugdunensis
Observed −log10(p)
Expected −log10(p)

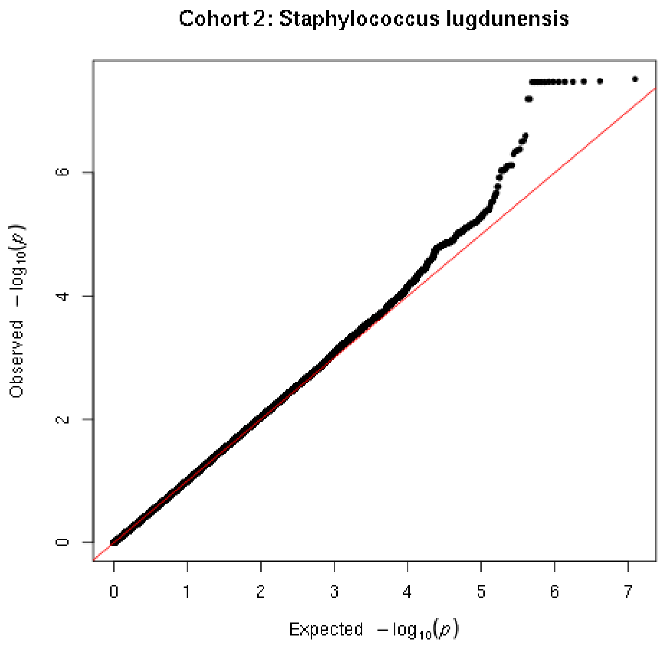
Cohort 2: Staphylococcus lugdunensis
Observed −log10(p)
Expected −log10(p)

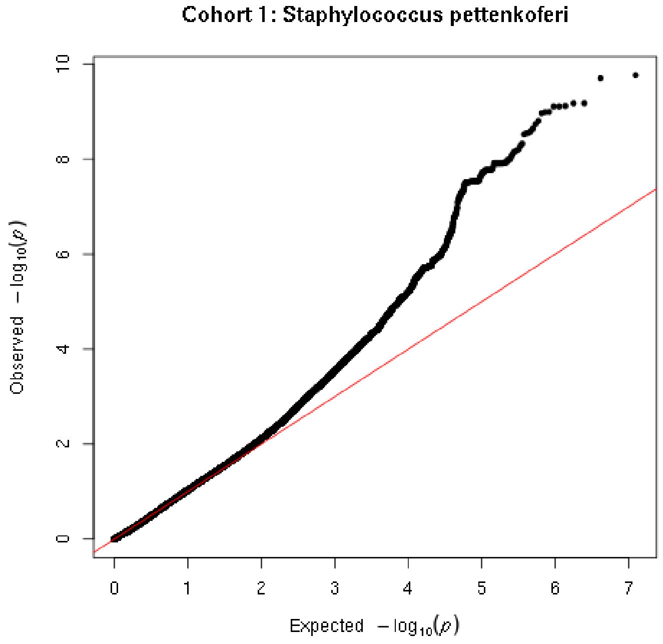
Cohort 1: Staphylococcus pettenkoferi
Observed −log10(p)
Expected −log10(p)

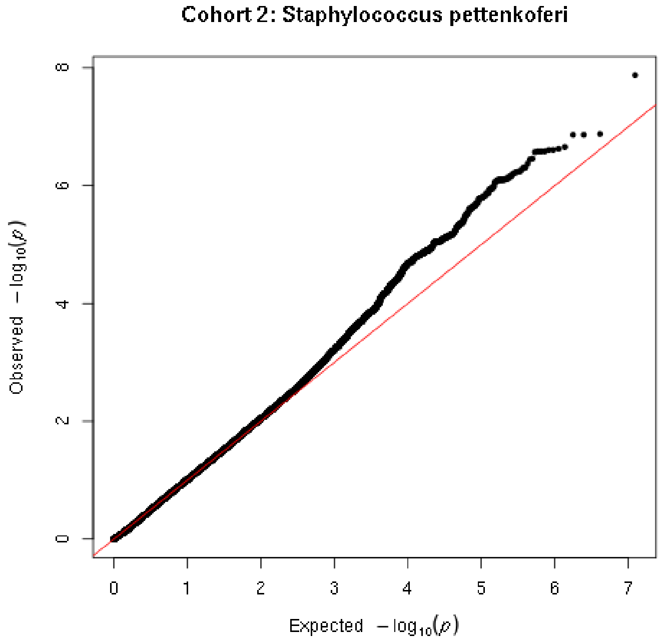
Cohort 2: Staphylococcus pettenkoferi
Observed −log10(p)
Expected −log10(p)

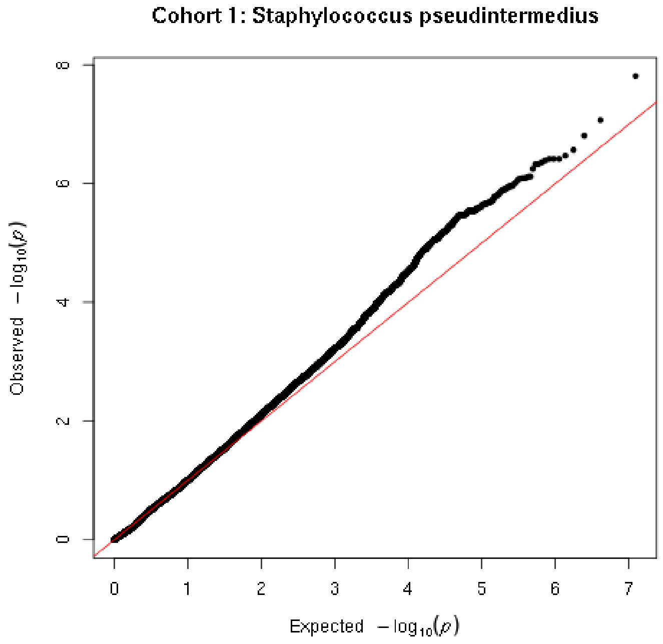
Cohort 1: Staphylococcus pseudintermedius
Observed −log10(p)
Expected −log10(p)

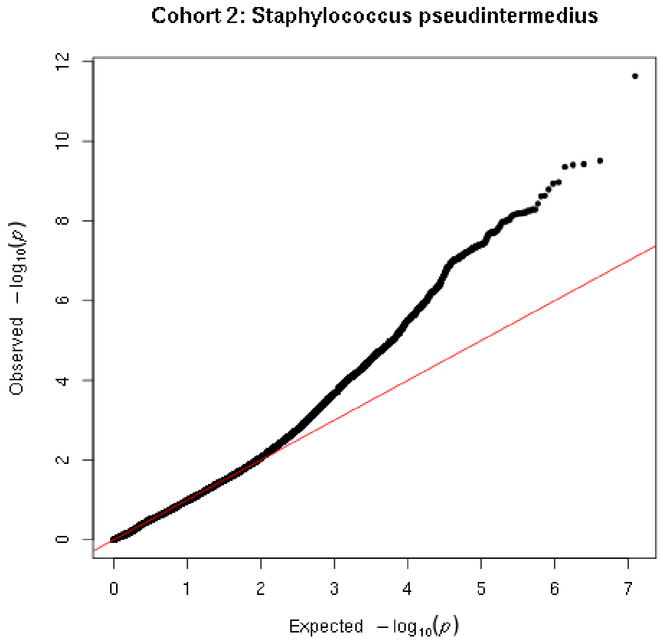
Cohort 2: Staphylococcus pseudintermedius
Observed −log10(p)
Expected −log10(p)

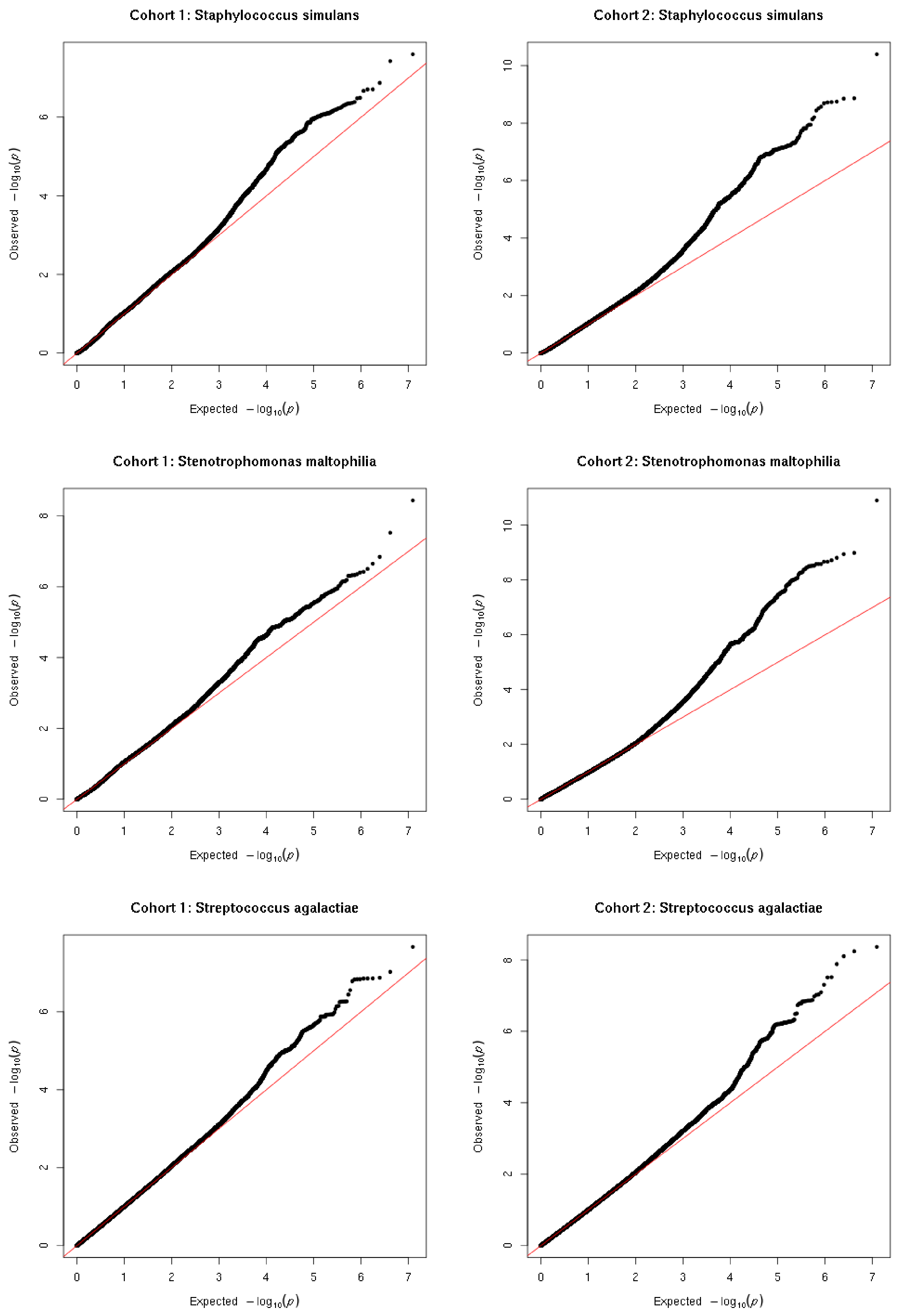
Cohort 1: Staphylococcus simulans
Cohort 2: Staphylococcus simulans
Cohort 1: Stenotrophomonas maltophilia
Cohort 2: Stenotrophomonas maltophilia
Cohort 1: Streptococcus agalactiae
Cohort 2: Streptococcus agalactiae
Observed $-\log_{10}(p)$
Expected $-\log_{10}(p)$

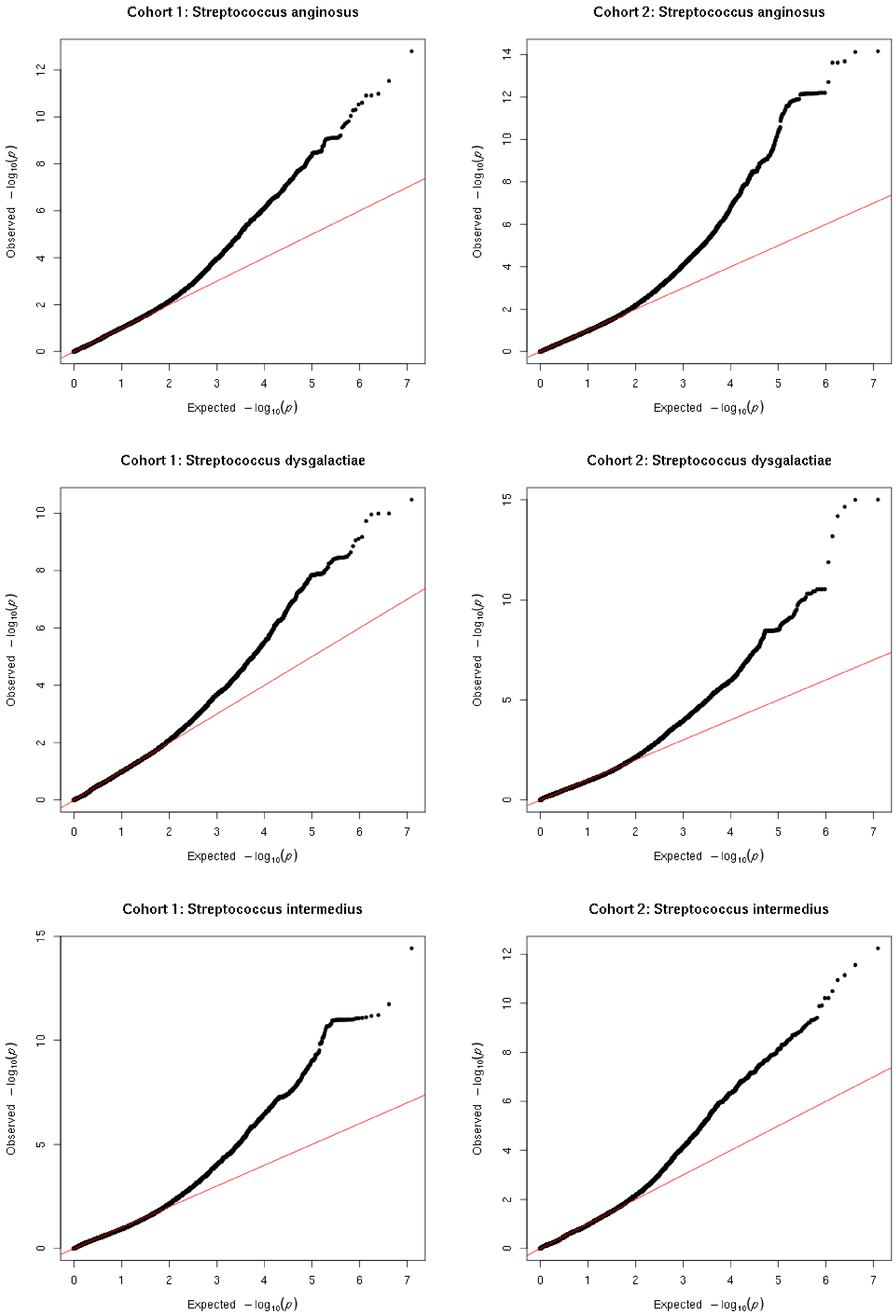
Cohort 1: Streptococcus anginosus
Cohort 2: Streptococcus anginosus
Cohort 1: Streptococcus dysgalactiae
Cohort 2: Streptococcus dysgalactiae
Cohort 1: Streptococcus intermedius
Cohort 2: Streptococcus intermedius
Observed $-\log_{10}(p)$
Expected $-\log_{10}(p)$

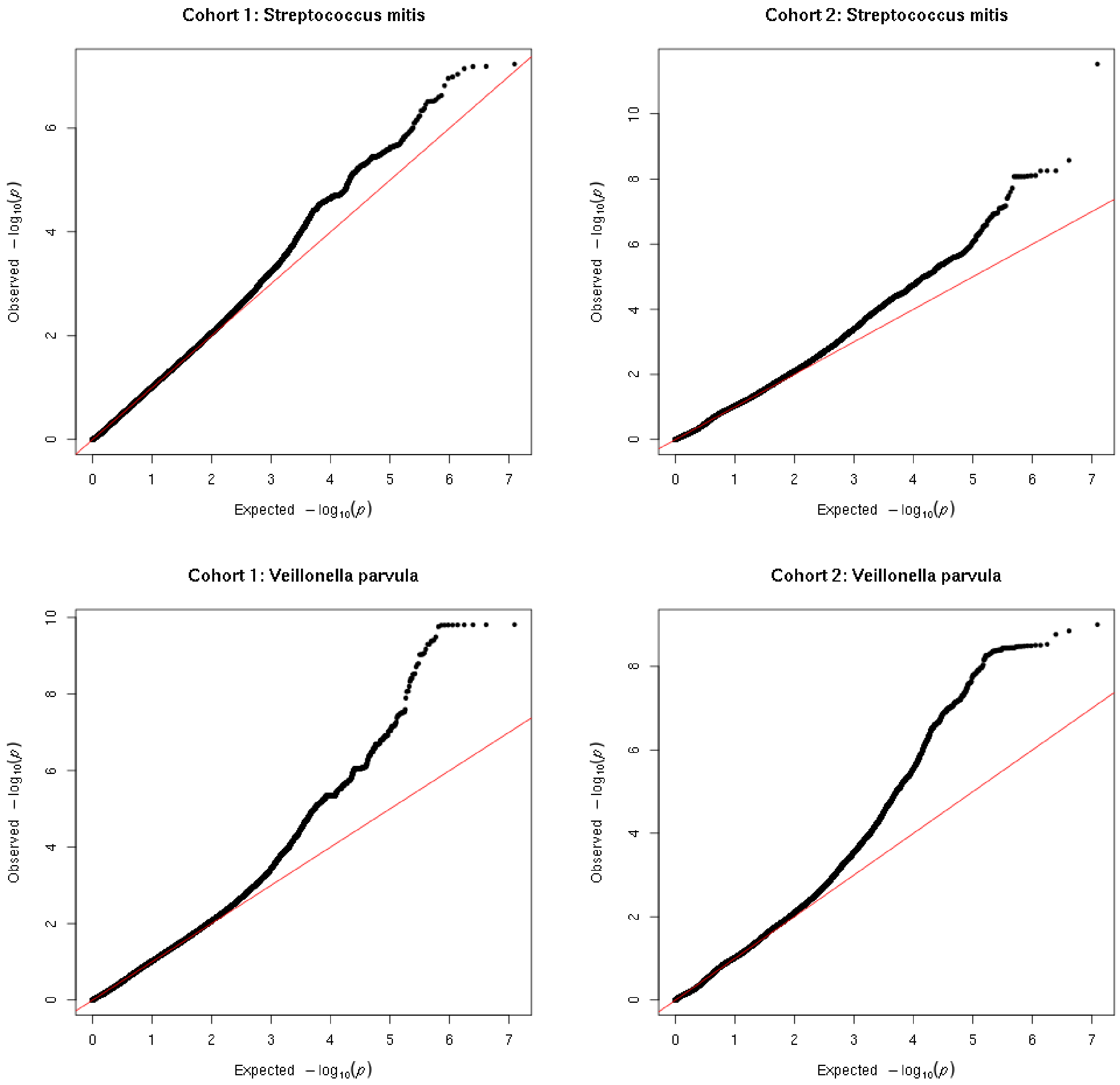

Cohort 1: Streptococcus mitis
Cohort 2: Streptococcus mitis
Cohort 1: Veillonella parvula
Cohort 2: Veillonella parvula
Observed $-\log_{10}(p)$
Expected $-\log_{10}(p)$

| Taxa | lambda.cohort1 | lambda.cohort2 |
|---|---|---|
| *Actinomyces neuii* | 0.89 | 1.04 |
| *Alcaligenes faecalis* | 0.87 | 1.03 |
| *Anaerococcus hydrogenalis* | 0.92 | 0.89 |
| *Anaerococcus lactolyticus* | 0.98 | 0.98 |
| *Anaerococcus obesiensis* | 0.81 | 0.64 |
| *Anaerococcus octavius* | 0.76 | 0.78 |
| *Anaerococcus prevotii* | 0.98 | 0.94 |
| *Anaerococcus vaginalis* | 0.99 | 0.99 |
| *Bacteroides fragilis* | 0.93 | 0.95 |
| *Burkholderia gladioli* | 1.06 | 1.05 |
| *Campylobacter ureolyticus* | 0.98 | 0.77 |
| *Citrobacter koseri* | 1.09 | 1.14 |
| *Corynebacterium amycolatum* | 0.82 | 0.93 |
| *Corynebacterium aurimucosum* | 1.01 | 0.88 |
| *Corynebacterium jeikeium* | 0.88 | 0.92 |
| *Corynebacterium simulans* | 0.92 | 0.96 |
| *Corynebacterium striatum* | 0.99 | 1.00 |
| *Corynebacterium tuberculostearicum* | 1.00 | 0.96 |
| *Cutibacterium acnes* | 0.93 | 0.97 |
| *Dialister propionicifaciens* | 0.99 | 0.92 |
| *Enterococcus faecalis* | 0.96 | 0.98 |
| *Enterococcus faecium* | 0.67 | 0.75 |
| *Escherichia coli* | 0.99 | 0.99 |
| *Finegoldia magna* | 1.00 | 0.99 |
| *Fusobacterium canifelinum* | 0.93 | 1.06 |
| *Fusobacterium nucleatum* | 1.00 | 0.95 |
| *Gemella morbillorum* | 1.14 | 1.03 |
| *Haemophilus parainfluenzae* | 0.85 | 0.83 |
| *Helcococcus kunzii* | 1.09 | 1.12 |
| *Klebsiella oxytoca* | 0.90 | 0.66 |
| *Klebsiella pneumoniae* | 0.93 | 0.98 |
| *Morganella morganii* | 0.93 | 0.57 |
| *Parvimonas micra* | 0.91 | 1.08 |
| *Pasteurella canis* | 1.03 | 0.98 |
| *Pelomonas saccharophila* | 0.81 | 0.63 |
| *Peptoniphilus asaccharolyticus* | 0.81 | 1.06 |
| *Peptoniphilus coxii* | 0.93 | 0.84 |
| *Peptoniphilus harei* | 0.97 | 1.01 |
| *Peptostreptococcus anaerobius* | 0.94 | 0.90 |
| *Porphyromonas bennonis* | 0.98 | 0.94 |

| | | |
|---|---|---|
| *Porphyromonas levii* | 0.92 | 0.85 |
| *Porphyromonas somerae* | 0.93 | 1.01 |
| *Prevotella bergensis* | 0.94 | 0.91 |
| *Prevotella bivia* | 0.91 | 0.96 |
| *Prevotella buccalis* | 0.91 | 0.71 |
| *Prevotella disiens* | 0.92 | 0.97 |
| *Prevotella melaninogenica* | 0.76 | 0.69 |
| *Prevotella timonensis* | 1.05 | 1.12 |
| *Proteus mirabilis* | 0.95 | 0.96 |
| *Pseudomonas aeruginosa* | 0.96 | 1.02 |
| *Serratia marcescens* | 0.96 | 0.81 |
| *Staphylococcus aureus* | 1.01 | 0.99 |
| *Staphylococcus capitis* | 0.99 | 0.90 |
| *Staphylococcus cohnii* | 0.85 | 0.84 |
| *Staphylococcus epidermidis* | 1.00 | 1.01 |
| *Staphylococcus haemolyticus* | 0.99 | 0.92 |
| *Staphylococcus hominis* | 0.96 | 0.84 |
| *Staphylococcus lugdunensis* | 0.97 | 0.96 |
| *Staphylococcus pettenkoferi* | 0.87 | 0.89 |
| *Staphylococcus pseudintermedius* | 0.83 | 0.88 |
| *Staphylococcus simulans* | 0.81 | 0.87 |
| *Stenotrophomonas maltophilia* | 0.79 | 0.95 |
| *Streptococcus agalactiae* | 0.98 | 0.97 |
| *Streptococcus anginosus* | 0.90 | 0.96 |
| *Streptococcus dysgalactiae* | 0.87 | 1.05 |
| *Streptococcus intermedius* | 1.09 | 0.81 |
| *Streptococcus mitis* | 0.92 | 0.75 |
| *Veillonella parvula* | 0.88 | 0.71 |
